# Combining stacked polygenic scores with clinical risk factors improves cardiovascular risk prediction in people with type 2 diabetes

**DOI:** 10.1101/2022.09.01.22279477

**Authors:** K Dziopa, N Chaturvedi, M. Vugt, J Gratton, R Maclean, A Hingorani, F W Asselbergs, C Finan, A F Schmidt

## Abstract

**Background:** Recommended CVD prediction models do not perform well in people with diabetes. We aimed to determine whether models combining polygenic scores (PGS) with clinical risk factors could more accurately predict 10-year risk of six facets of CVD, including: coronary heart disease (CHD), heart failure (HF), and atrial fibrillation (AF).

**Methods:** Three groups were selected from the UK Biobank: 143,459 control participants without diabetes or a history of CVD, 5,229 with diabetes but without CVD, and 1,621 with diabetes and a history of CVD. Data from 29 phenotype-specific polygenic scores (PGS) were stacked and combined with clinical risk-factors. Performance was evaluated using a 20% independent hold-out sample, with results stratified on duration of diabetes.

**Results:** In people without diabetes combining the stacked PGS with clinical risk factor modestly outperformed models that exclusively used clinical risk factors, with the largest improvement observed for AF (c-statistic difference: 0.03). In people with diabetes, models that combined the stacked PGS with clinical risk factors showed marked improved performance compared to the risk factor only models. This difference was largest in people with newly diagnosed diabetes (without a history of CVD), with a PGS + clinical risk factor model c-statistic: 0.83 (95%CI 0.83; 0.84) for CHD and 0.84 (95%CI 0.82; 0.85) for HF, compared to a clinical risk factor model c-statistic: 0.68 (95%CI 0.68; 0.69) and 0.60 (95%CI 0.58; 0.62) for CHD and HF respectively.

**Conclusions:** Combining PGS with clinical risk factors improves CVD risk prediction in people with diabetes.

**Research in context:** *What is already known about this subject?:*

- Cardiovascular disease (CVD) remains a prominent cause of morbidity and mortality for people with type 2 diabetes. The currently available CVD prediction models do not provide sufficiently accurate prediction in people with diabetes, prohibiting much-needed personalization of management strategies.
- In the general population, phenotype-specific polygenic scores (PGS) have shown to modestly improve CVD risk prediction. However, models for CVD prediction in the general population are often already highly accurate, limiting the scope for PGS to further improve performance.
- Given the multifactorial etiology of CVD, combining information (stacking) from multiple trait-specific PGS (e.g., on CHD, LDL-C and blood pressure) is expected to improve performance.

*What is the key question?:*

- What is the added benefit of incorporating PGS with conventional clinical risk factors in CVD prediction for people with type 2 diabetes?

*What are the new findings?:*

- In people with diabetes, models that combined the stacked PGS with clinical risk factors showed marked improved performance compared to the risk factor-only models.
- While age was the predominant risk factor in people without diabetes, in people with diabetes the contribution of age was outranked by our stacked PGS.
- Model performance depended on the duration of diabetes, with models performing better in people with a recent diagnosis, for example in this group the c-statistic for CHD was 0.83 (95%CI 0.83; 0.84), and for HF 0.84 (95%CI 0.82; 0.85).

*How might this impact on clinical practice in the foreseeable future?:*

- Combining PGS with clinical risk factors improves CVD risk prediction in people with diabetes. Incorporating PGS in risk prediction models may offer unique possibilities to reliably identify people with a meaningful risk of developing CVD.

**ACRONYMS:** 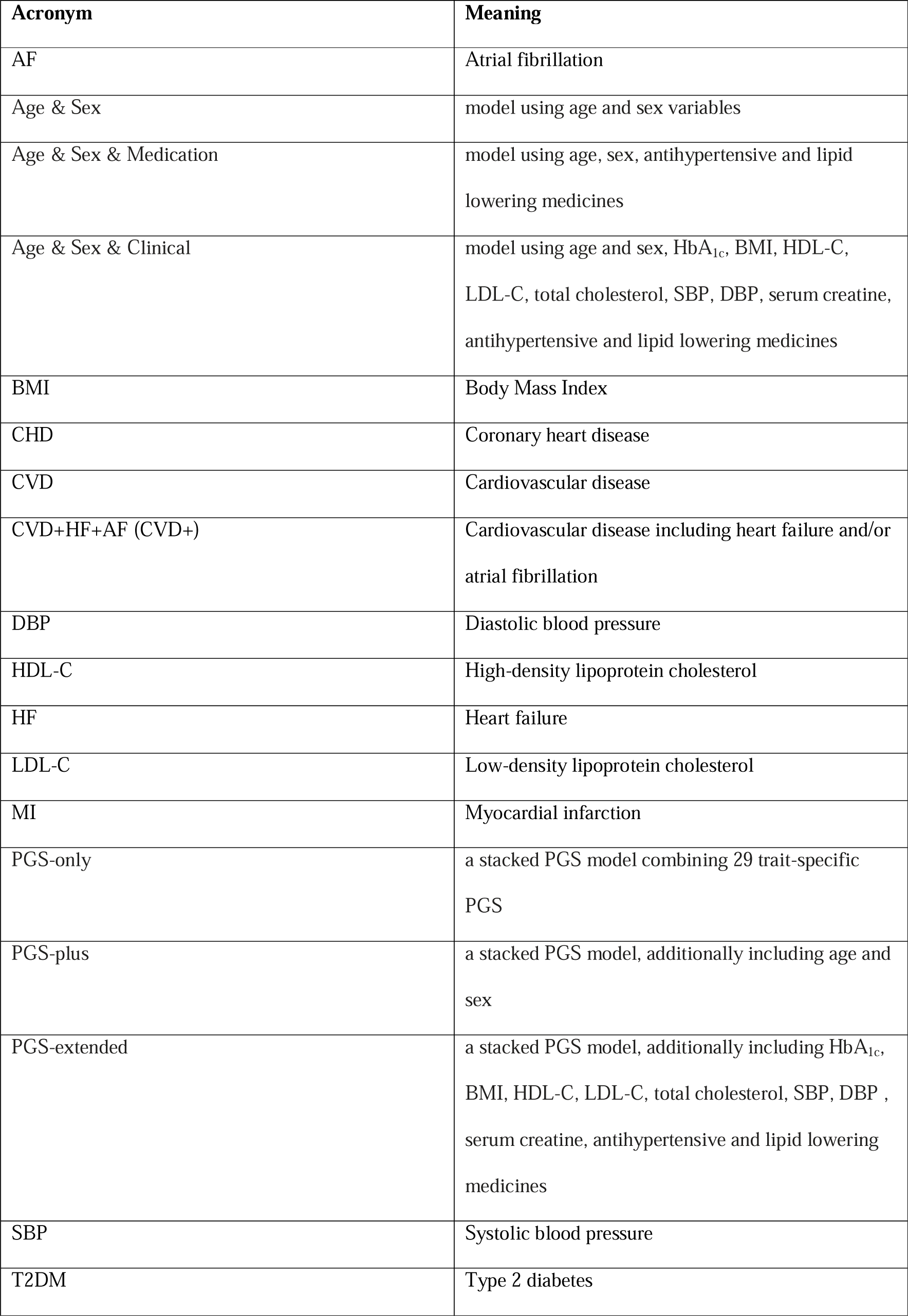

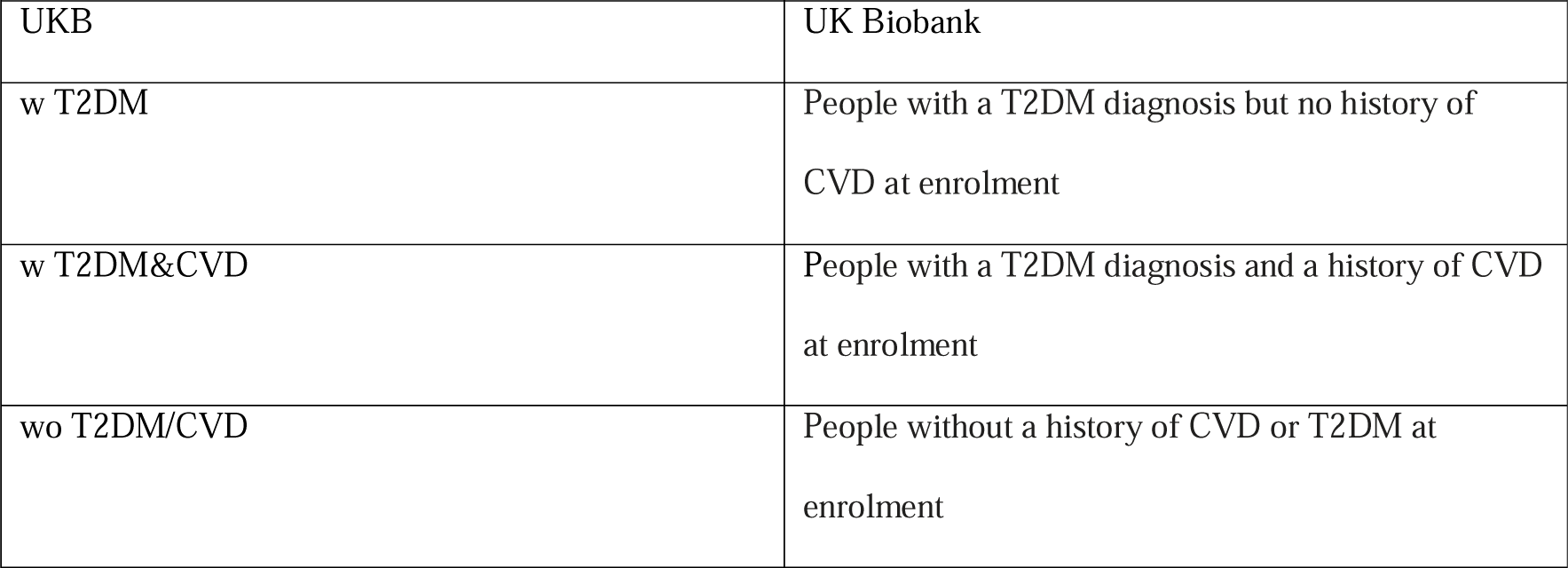

## INTRODUCTION

In clinical practice, mitigation of cardiovascular disease (CVD) is guided by risk prediction algorithms. The UK National Institute for Health and Care Excellence (NICE^1^) guidelines recommend the use of the QRISK2 for CVD risk prediction for people with and without diabetes. Similarly, the American College of Cardiology/American Heart Association (ACC/AHA) recommends estimating the 10-year risk of CVD using the Atherosclerotic Cardiovascular Disease (ASCVD) risk score^2^ which includes diabetes as a predictor.

Despite major advances in treatment, CVD remains the main cause of morbidity and mortality in people with type 2 diabetes (T2DM)^3^. Recently, we performed a head-to-head comparison of 22 cardiovascular risk scores, including those designed for people with T2DM alone as well as population scores, showing that all scores discriminated poorly (e.g., c-statistic less than 0.70), and performed markedly worse than originally reported when tested in the general population^4^. This suggests that the current type of prognostic CVD models and the included risk-factors are less applicable to people with T2DM.

In general population studies, polygenic scores (PGS) have been shown to only modestly improve risk prediction of CVD^5^. The shared *genetic* burden between type 2 diabetes and coronary heart disease (CHD) ^6,7^ suggests that people with diabetes are at a higher CVD risk. Polygenic scores attempt to capture an individual’s genetic susceptibility to a trait or disease by summarizing information on multiple (sometimes thousands) of genetic variants, often identified through genome-wide association studies (GWAS) ^8^. The UK Department of Health and Social Care, and the US Centres for Disease Control and Prevention have set-up programs to incorporate genomic advances into routine healthcare, aiming to improve the diagnosis, risk-stratification and treatment of diseases ^9^ ^10^ . Many PGS focus exclusively on discrimination (the ability of a model to separate individuals with and without an outcome)^11^, and do not provide individual risk predictions, making these scores difficult to incorporate into pre-existing risk-based management strategies. As such healthcare professionals are now increasingly confronted with genetic data but lack guidance on their use.

In the current study we aimed to explore the added benefit of incorporating PGS with conventional clinical risk factors and biomarkers to improve CVD prediction for people with type 2 diabetes, specifically deriving models that provide estimates of 10-year risk which can be incorporated within current guidelines on risk-stratification. To optimize the PGS predictive potential we created 29 univariable PGS, and combined these to derive a multivariable “stacked” PGS model trained to predict individually CHD, ischemic stroke, heart failure (HF) and atrial fibrillation (AF), a collective CVD definition (combining CHD, stroke, and peripheral arterial disease (PAD)), and a broader definition of major CVD+ additionally including HF and AF - outcomes which are more common in people with diabetes^12, 13^. Models were derived for three distinct UK Biobank (UKB) groups: people without pre-existing CVD or type 2 diabetes diagnosis at enrolment, and people with T2DM stratified by CVD history at the time of enrolment. Performance was evaluated in an independent 20% testing set, for diabetes patients this was additionally stratified on duration (prevalent or incident T2DM).

## METHODS

### Data source

Data was sourced from the UKB, a cohort of ∼500,000 men and women aged 40-69 years between 2006 and 2010 enrolled from primary care registers across the UK ^14^. At enrolment, questionnaire, nurse interview and clinic investigations collected data on pre-existing conditions, medication, and CVD risk factors. Additionally, a blood sample was drawn for biomarkers and DNA extraction. Genetic data is available for the majority of participants through genotyping arrays and exome sequencing. Due to the European focus of most GWAS, we excluded participants of non-European descent, and additionally removed related individuals (a kinship coefficient greater or equal to 0.0442). Applying additional quality control steps (Appendix Figure 1) and linking to hospital episode statistics (HES) and general practitioners (GP) data (GP data is only available on around half of samples) resulted in 341,516 participants; see Appendix Figure 2.

The study sample was stratified into three groups based on T2DM and CVD histories at the time of UKB enrolment: 1) (wo T2DM/CVD) without a history of CVD or evidence of T2DM at enrolment (N = 143,459), 2) (w T2DM) individuals with type 2 diabetes but no history of CVD (N = 5,229), before the enrolment date or 3 months later, and 3) (w T2DM & CVD), individuals with diabetes and a history of CVD (N = 1,621). To predict 10 year CVD risk, individuals were followed-up from the time of UKB enrolment, recording of the first occurrence of a CVD event, death, or end of study, or up to 10 years after enrolment.

Non-genetic predictors were extracted from the cross-sectional UKB assessment centre data, and the longitudinal General Practice (GP) records, taking the measurement closest to the enrolment date and no more than 1 year before or 3 months after enrolment. Specifically, data were extracted on sex, age (years), glycated haemoglobin (HbA1c, mmol/mol), body mass index (BMI), high-density lipoprotein cholesterol (HDL-C, mmol/L), low-density lipoprotein cholesterol (LDL-C, mmol/L), total cholesterol (mmol/L), systolic blood pressure (SBP, mm Hg), diastolic blood pressure (DBP, mm Hg), smoking status (never, previous, current); see Appendix Tables 1-3. Prescription of antihypertensive and lipid-lowering medicines was extracted from the year before enrolment; Appendix Figure 3 and Appendix 2 for the drug keywords.

CVD was defined as the occurrence of fatal or non-fatal myocardial infarction (MI), sudden cardiac death, ischemic heart disease, fatal or non-fatal stroke or PAD after the start of follow-up. We additionally considered a broader definition of CVD, also including heart failure (HF) and / or atrial fibrillation (AF): ‘CVD+’, as well as the individual CVD components: CHD, stroke, AF, and HF; see Appendix Methods.

People with T2DM were identified using a CALIBER phenotyping algorithm (combines data from GP records and HES) enhanced by HbA_1c_ measurements and diabetes related medication (Appendix Figure 4, Appendix Table 4), labelling participants as prevalent (known) diabetes (based on HES and GP records), incident (previously undiagnosed) diabetes (HbA_1c_ at time of enrolment 2: 48 mmol/mol), pre-diabetes (from 42 to 48 mmol/mol), normoglycemic (from 35 to less than 42 mmol/mol), and low HbA1C (< 35 mmol/mol) ^15^. People with prevalent and incident (newly diagnosed) diabetes were included in the current manuscript.

### Derivation of genetic scores and model stacking

Given the multifactorial origin of both CVD and T2DM, where for example increased BMI, hypertension and hypercholesterolemia are known risk factors for CVD and T2DM, we a priori identified 29 GWAS (Appendix Table 5) with publicly available data on the genetic associations (e.g., point estimates) that might be relevant for CVD prediction. Next, we applied a 10-fold cross-validation grid-searching algorithm to identify the optimal (highest c-statistic) parametrization of each univariable PGS (e.g., a PGS for LDL-C) to predict each type of CVD. The following PGS parameters were considered 1) the variant specific p-value inclusion threshold, 2) the number of correlated variants contributing to the score (LD threshold), 3) the minor allele frequency (MAF) threshold of the genetic variants in the score, and 4) whether the scores should by their variant-specific point estimate (weighted or unweighted). This resulted in 29 GWAS-specific univariable PGS which were optimized to predict the six CVD outcomes in each of the three groups.

The 29 optimized univariable PGS were combined into a single *stacked* PGS model (referred as “PGS-only”) using four distinct machine learners: generalized linear model (GLM)^16^, Lasso Regression (LR; Logistic Regression model with L1 regularization)^17^, Random Forest (RF)^18^, and a joint model of LR and RF, with 10-fold cross-validation to identify the model with the best discriminative ability (c-statistic). To prevent potential overfitting, the more flexible RF models were only applied if the number of training data cases was larger than 250; see Appendix Table 6 for the RF hyperparameters that were considered.

### Joint modelling of risk factors and stacked PGS

PGS performance was compared against CVD risk prediction using clinical risk factors, both alone and in combination. Three models were defined based on their candidate set of predictors: 1) age and sex (Age & Sex), 2) additionally considered clinical characteristics, HbA_1c_, BMI, HDL-C, LDL-C, total cholesterol, SBP, DBP, serum creatine, c-reactive protein, and antihypertensive and lipid lowering medicines (Age & Sex & Clinical), and 3) using age, sex, and medicines exclusively (Age & Sex & Medication). The Age & Sex & Clinical model was affected by missing biomarker data (Table 1), which were imputed using single imputation procedure leveraging the MICE package^19^, with results were compared to a complete case analysis. The Age & Sex & Medication model was not affected by missing data and was used to indirectly assess the impact of missing data. Finally, the value of combining PGS with clinical risk factors was evaluated by combining 1) the stacked PGS with sex and age (PGS-plus), and 2) a PGS-extended model including the Age & Sex & Clinical candidate features.

**Table 1.** Clinical characteristics of UK biobank participants stratified by group: participants without a history of CVD and T2DM (wo T2DM/CVD), participants with type 2 diabetes (w T2DM), participants with a history of CVD prior to a T2DM diagnosis (w T2DM&CVD).

| Clinical characteristic | wo T2DM/CVD |  | w T2DM |  | w T2DM&CVD |  |
| --- | --- | --- | --- | --- | --- | --- |
|  | Mean (SD) or N(%) | Missing data (%) | Mean (SD) or N(%) | Missing data (%) | Mean (SD) or N(%) | Missing data (%) |
| Total no. of individuals | 143,459 |  | 5,229 |  | 1,621 |  |
| Women (%) | 80,733 (56.3) | 0 | 2,091 (40.0) | 0 | 401 (24.7) | 0.0 |
| Age (years) | 56.4 (8.0) | 0 | 59.8 (7.0) | 0 | 62.8 (5.4) | 0.0 |
| Smoking Status: |  |  |  |  |  | 0.3 |
| Never | 80,991 (56.5) |  | 2,384 (45.6) |  | 485 (29.9) |  |
| Previous | 48,110 (33.5) |  | 2,299 (44.0) |  | 905 (55.8) |  |
| Current | 13,924 (9.7) |  | 511 (9.8) |  | 220 (13.6) |  |
| BMI (kg/m<sup>2</sup>) | 27.2 (4.6) | 0.2 | 31.6 (5.8) | 0.4 | 32.1 (5.5) | 0.6 |
| HDL cholesterol (mmol/L) | 1.5 (0.4) | 9.6 | 1.2 (0.3) | 4.5 | 1.1 (0.3) | 4.8 |
| Total cholesterol (mmol/L) | 5.8 (1.1) | 4.3 | 4.5 (1.0) | 4.3 | 4.3 (1.0) | 4.9 |
| LDL cholesterol (mmol/L) | 3.7 (0.8) | 3.8 | 2.7 (0.8) | 3 | 2.5 (0.7) | 3.0 |
| HbA1C (mmol/mol) | 35.1 (4.5) | 4.5 | 53.2 (13.5) | 0.8 | 54.4 (13.9) | 0.4 |
| SBP (mm Hg) | 140.2 (19.7) | 0.1 | 145.0 (18.2) | 0.2 | 143.3 (20.5) | 0.1 |
| DBP (mm Hg) | 82.5 (10.7) | 6.7 | 82.3 (10.1) | 5.1 | 78.5 (10.7) | 4.2 |
| Antihypertensive medication | 20,437 (14.2) |  | 2,599 (49.7) |  | 1,112 (68.6) | 0.0 |
| Lipid lowering medication | 12,738 (8.9) |  | 3,159 (60.4) |  | 1,135 (70.0) | 0.0 |

n.b. Clinical information was obtained from the UKB assessment centre data, and the longitudinal GP records, selecting the measurements closest to baseline from within a time window of 1 year before and 3 months after baseline. Medication information reflect any prescription within the year prior the baseline date (i.e., UKB enrolment or diagnosis of T2DM). SD refers to standard deviation.

### Estimating model performance

Performance in terms of discrimination (c-statistic) and calibration (calibration-in-the-large, calibration slope) was determined through an independent (20% split) test data. Feature importance of each PGS and non-genetic variable was evaluated using a permutation feature importance algorithm, assessing the c-statistic change in the test data; see Appendix Methods. Additionally, Net Reclassification Index (NRI) tables were used to compare the PGS-extended model against the Age & Sex & Clinical (which differed solely on their PGS inclusion) using the following risk cut-offs: <0.10: low risk, between 0.10 and 0.20: intermediate risk, >0.20: high risk. A number of sensitivity analyses are described in the Appendix methods and Appendix Table 7.

## RESULTS

### Patient characteristics

Data were available on 143,459 participants without T2DM and CVD at baseline “wo T2DM/CVD”, 5,229 participants with T2DM but without a history of CVD at the time of diagnosis “w T2DM”, and 1,621 individuals who had a history of CVD at the time of T2DM diagnosis “w T2DM&CVD”; Table 1. Participants with disease were on average older (w T2DM&CVD: 62.8 SD 5.4, w T2DM: 59.8 SD 7.0, wo T2DM/CVD: 56.4 SD 8.0), less likely to be female (w T2DM&CVD: 24.7%, w T2DM: 40.0%, wo T2DM/CVD: 56.3%), had a higher BMI (w T2DM&CVD: 32.1 kg/m^2^ SD 5.5, w T2DM: 31.6 SD 5.8, wo T2DM/CVD: 27.2 SD 4.6) and a higher HbA_1c_ (w T2DM&CVD: 54.4 mmol/mol SD 13.9, w T2DM: 53.2 SD 13.5, wo T2DM/CVD: 35.1 SD 4.5). During a median follow-up time of 10 years, CVD, AF or HF events occurred in 13,133 (9.2%) “wo T2DM/CVD” individuals, 1,057 (20.2%) “w T2DM” individuals, and 1,295 (79.9%) of the “w T2DM&CVD” individuals; Appendix Table 23.

### Stacking multiple PGS into multivariable prognostic models

We identified the optimal parametrisation of 29 univariable PGS by training each to predict six CVD outcomes. Through grid-searching we found that most selected PGS utilized a p-value threshold lower than the conventional threshold of GWAS significance^20^ (less than 20% of the PGS used a p-value of 5x 10^-8^), and showed a clear preference for approximately independent variants (selecting an r-squared of 0.20 or 0.01 at least 50% of the time); Figure 1. Subsequently the univariate GWAS-specific PGS were combined into multivariate (stacked) models to predict each of the six CVD-related outcomes. See Appendix Results, Appendix Table 5, 8 and Appendix Figures 5-16.

**Figure 1.**
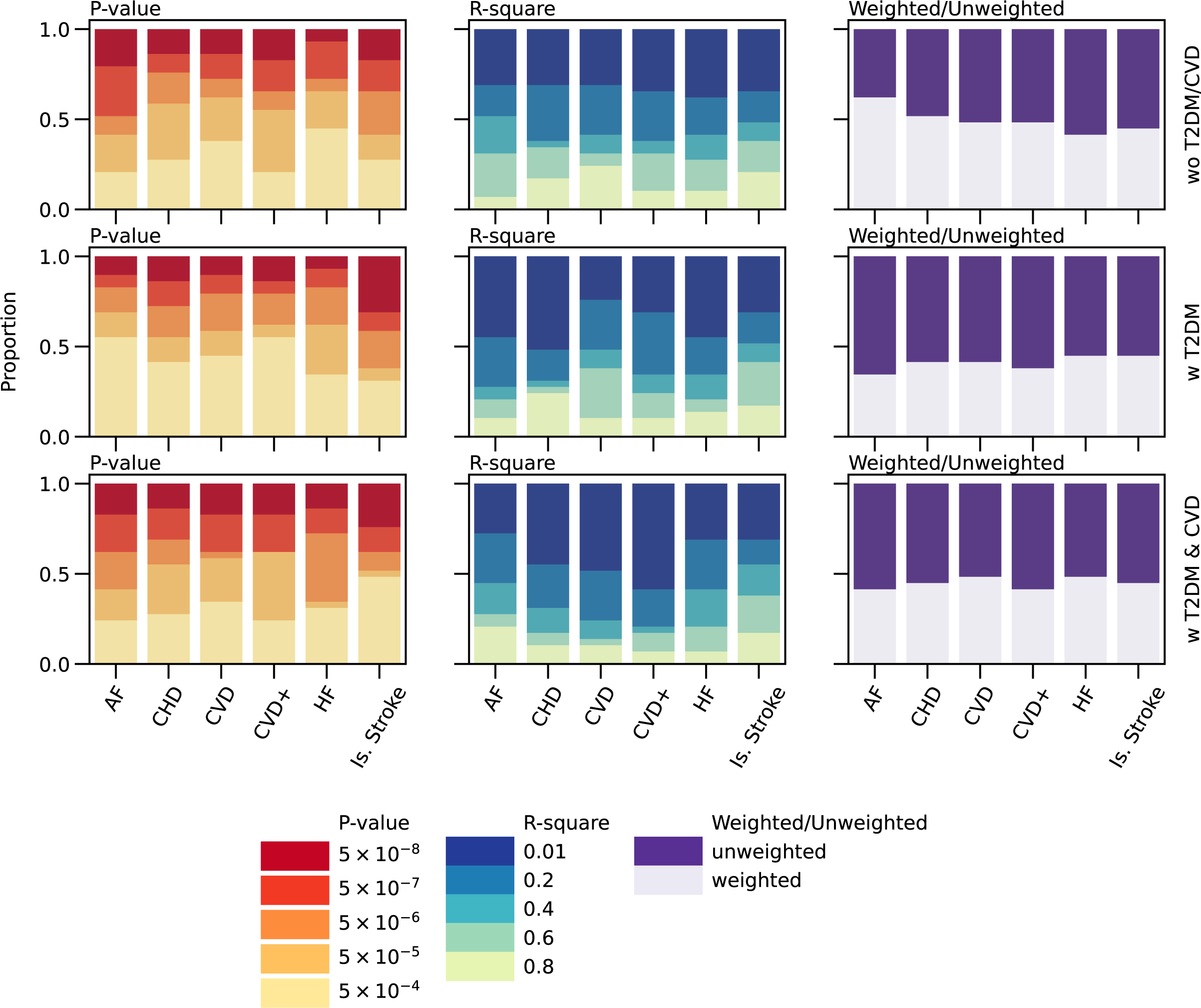
The polygenic (PGS) parameters selected through a cross-validated grid search. N.B. Results were stratified by outcome (x-axis) and group (rows). The parameterization of 29 PGS in terms of GWAS p-value, linkage disequilibrium r-squared, and whether each variant should be weighted by its regression coefficient (weighted/unweighted). The proportion therefore reflects division by 29. The optimal combination of parameters was determined through a grid-search algorithm selecting combination with the largest training data c-statistic for each outcome listed on the x-axis). Subjects were stratified as followed “wo T2DM/CVD: participants without T2DM or CVD at baseline, “w T2DM”: participants with diabetes at baseline, “w T2DM&CVD”: participants with T2DM at baseline and a history of CVD.

### Combining clinical characteristics with stacked PGS

We evaluated the stacked PGS performance to models combining the PGS with clinical risk factors, and prediction models that exclusively used clinical risk factors. The PGS only model had a low c-statistic in people without diabetes (below 0.60) but attained higher c-statistic values (above 0.60) for the “w T2DM” group (Figure 2). Simply adding age and sex improved discrimination considerable improved performance for people without diabetes, for example the CVD c-statistic was 0.58 (0.58; 0.58) for the PGS only model, compared to 0.70 (95%CI 0.70; 0.70) after adding age and sex (Figure 2). Including age and sex did not result in similar marked improvements people with diabetes, indicating that these variables do not explain CVD as well in people with diabetes (Figure 2, Appendix Tables 8 - 9). Feature importance (Appendix Figure 23) confirmed the importance of age for CVD prediction in people without diabetes, and conversely showed that the stacked PGS was the most important feature for people with diabetes; Appendix Figures 17-18.

**Figure 2.**
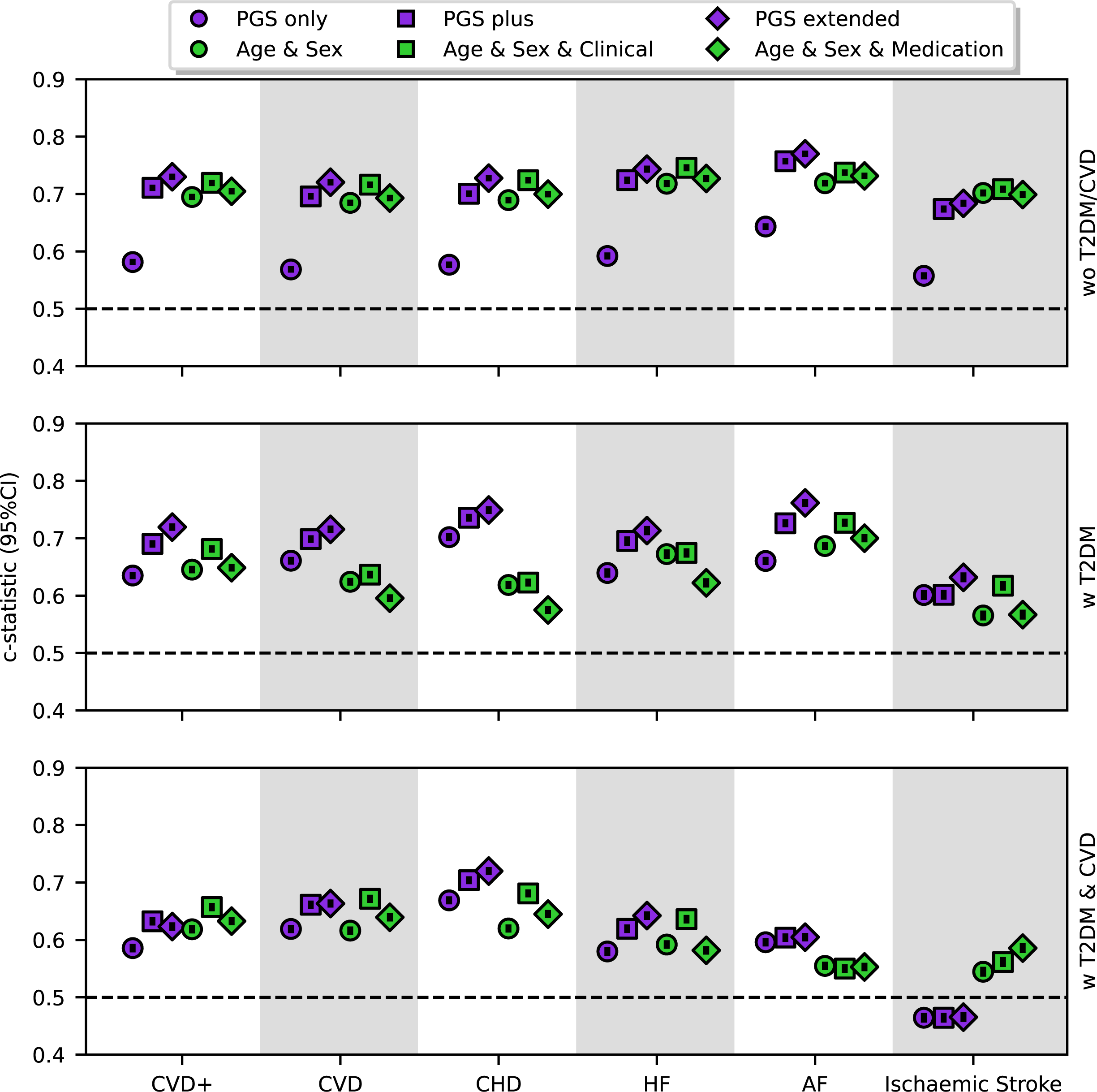
Discriminative performance of stacked polygenic scores (PGS) and conventional risk prediction models attempting to predict the 10-year risk of CVD in patient with or without diabetes. N.B. The c-statistics were estimated using an independent test dataset, not used in model training. The stacked PGS combined genetic scores for at most 29 GWAS specific PGS, and were step-wise elaborated to consider age and sex at baseline (PGS-plus) and additionally smoking status, blood lipids, blood pressure, BMI, HbA1c, and medicines (PGS-extended). The conventional, non-genetic models used information only on age and sex (Age & Sex), the second model additionally considered clinical characteristics along with antihypertensive and lipid lowering medicines (Age & Sex & Clinical), and the last model used age, sex and medication biomarkers (Age & Sex & Medication). Individuals were stratified as followed “wo T2DM/CVD: participants without T2DM or CVD at baseline, “w T2DM”: participants with diabetes at baseline, “w T2DM&CVD”: participants with T2DM at baseline and a history of CVD.

Adding further clinical characteristics including smoking status, BMI, HbA_1c_, blood lipid measurement, blood pressure, and antihypertensive and lipid-lowering medication (PGS-extended) improved the discriminative ability of most of the models (Figure 2, Appendix Table 10), when compared to PGS-plus results. For example the CHD c-statistic was: 0.73 (95%CI 0.73; 0.73), 0.75 (95%CI 0.75; 0.75), and 0.72 (95%CI 0.726; 0.72) for the “wo T2DM/CVD”, “w T2DM”, and “w T2DM&CVD” participants groups. Feature importance showed a similar pattern as before, with the stacked PGS the most important predictor for the two T2DM groups, and age the most important predictor in people without diabetes (Appendix Tables 11-13, Appendix Figure 19). The calibration plots (Appendix Figure 24) of the PGS-extended models indicated that these models under-estimated the risk of CVD+, CVD and CHD in the “w T2DM&CVD” group, but otherwise showed reasonable calibration; See Appendix Figures 15-20 and Appendix 3 for the remaining calibration plots and distributions of predicted risk.

Comparing the models combining models, to prediction models exclusively sourcing clinical risk factors revealed near equivalent performance in people without diabetes (Figure 2); Appendix Tables 10, 14 – 16. In people with diabetes however, the clinical risk factor only models (Figure 2) often performed worse than models combining the PGS and the clinical risk factors. For example, the c-statistic for 10-year risk of CHD was 0.62 (95%CI 0.62; 0.63) in “w T2DM”, and 0.68 (95%CI 0.68; 0.68) in “w T2DM&CVD” for a non-genetic model including age, sex and clinical measurements “Age & Sex & Clinical”, whereas the PGS-extended c-statistic was 0.75 (95%CI 0.75; 0.75) in “w T2DM” and 0.72 (95%CI 0.72; 0.72) in “w T2DM&CVD”; Appendix Tables 10, 16.

Finally, NRI tables comparing “Age & Sex & Clinical” and PGS-extended models in their ability to correctly classify subjects in low, mid, high-risk groups were calculated for CHD (Appendix Table 24, Appendix Tables 17–18), which indicated that improved performance of the PGS-extended model in “w T2DM” was due to assigning a higher risk to subjects who would develop CHD event and assigning lower risk to individual who would not develop CHD in the considered 10-years follow-up. Sensitivity analyses are described in the Appendix Results including Appendix Tables 19 – 20.

### Performance in people with prevalent or incident diabetes

Given that diabetes duration influences CVD risk factor associations, we compared model performance stratified by prevalent (before UKB enrolment date) and incident type 2 diabetes (at the UKB enrolment date). Similar as before the PGS-extended model outperformed the “Age & Sex & Clinical” model in most cases (Figure 3). Performance of the PGS-extended model (combining the PGS with clinical risk factors) meaningfully improved in people with newly diagnosed “incident” diabetes without a history of CVD. For example the c-statistic for CHD was 0.83 (95%CI 0.83; 0.84), and for HF 0.84 (95%CI 0.82; 0.85). Similar, but attenuated, performance was observed in the “w T2DM&CVD” group stratified for timing of diabetes diagnosis; Figure 3, Appendix Tables 21 - 22. Despite the difference in model accuracy between people with prevalent and incident diabetes, we did not observe a difference in their cumulative incidence of disease risk; See Appendix Figure 21-22.

**Figure 3.**
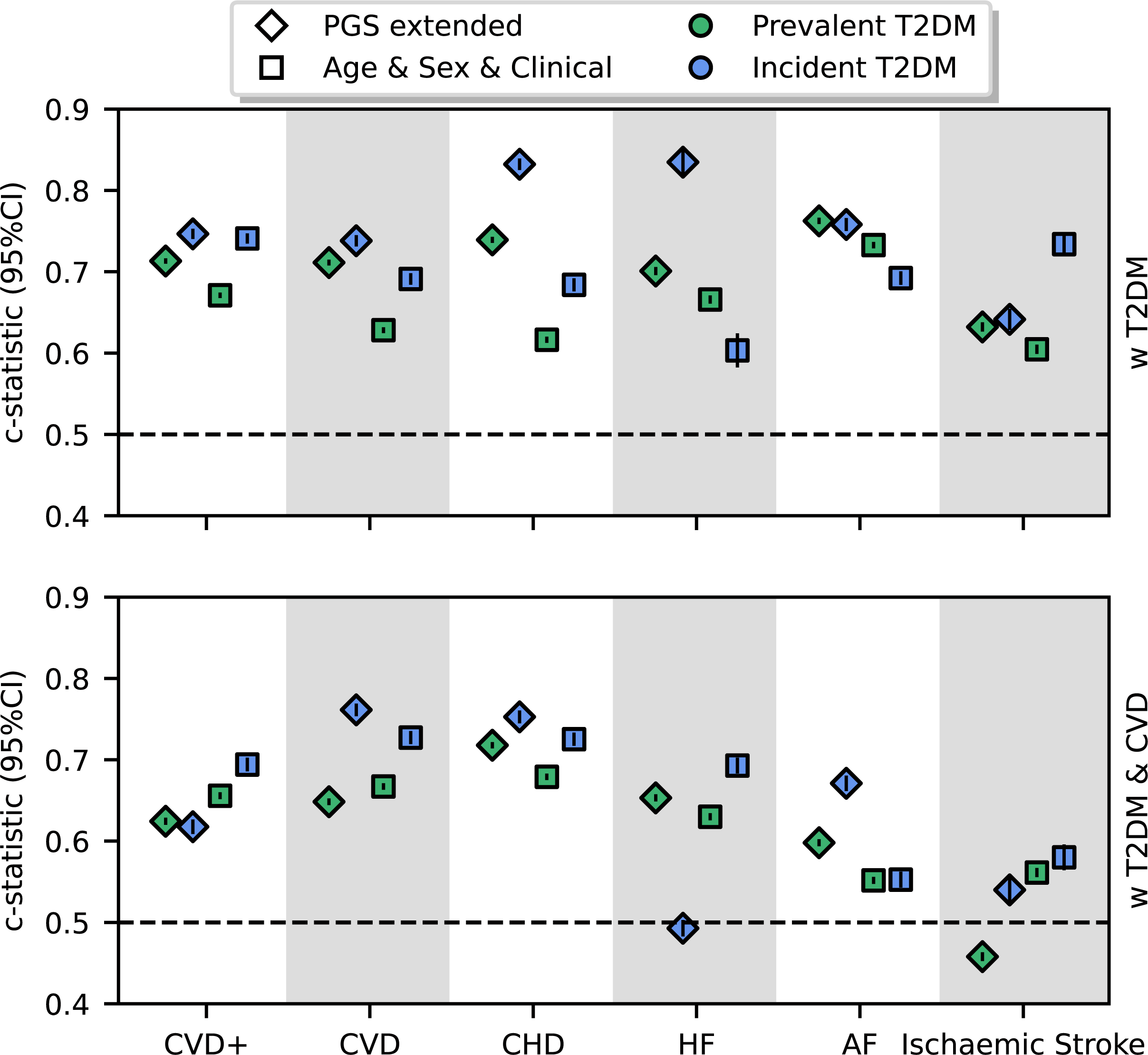
The discriminative performance of models predicting CVD in people with prevalent and incident type 2 diabetes N.B. The c-statistics were estimated using an independent test dataset, not used in model training. The stacked PGS combined genetic scores for at most 29 GWAS specific PGS, and was combined with non-genetic risk factors: smoking status, blood lipids, blood pressure, BMI, HbA1c, and medicines (PGS-extended). The Age & Sex & Clinical model only include the clinical risk factors. Individuals were stratified on history of CVD and timing of diabetes diagnosis (prevalent: a diagnosis before enrolment, incident: newly diagnosed during enrolment).

## DISCUSSION

In the current study we showed that while PGS are important predictors for the 10-years risk of CVD, univariable their discriminative potential was modest and by themselves the PGS could not accurately predict who developed CVD. When considering multiple potential predictor variables the PGS was however an important predictors, and in people with diabetes the PGS was even the most important predictors outranking age; the most important predictor in people without diabetes. Clinical risk factor model with and without addition of a PGS did not meaningfully differ in their ability to predict 10-year risk of six types of CVD in people without diabetes. In people with diabetes however, the PGS + clinical risk factor models often outperformed models exclusively using clinical risk factors. For example, the c-statistic for 10-year risk of CHD using the “Age & Sex & Clinical” model was 0.62 (95%CI 0.62; 0.63) in the “w T2DM” group, compared to 0.75 (95%CI 0.75; 0.75) when adding the PGS. The PGS + clinical risk factor models showed the best calibration compared with other analysed models. We additionally showed that model performance changed with the duration of diabetes diagnoses, finding censurably improved performance in people with a recent diagnosis, for example in this group the c-statistic for CHD was 0.83 (95%CI 0.83; 0.84), and for HF 0.84 (95%CI 0.82; 0.85).

Previous applications of PGS have often attempted to predict a combination of incident and prevalent disease, or even exclusively focused on the latter. Stratified analyses have shown that models for prevalent disease often show a better discriminative ability than similar models for incident disease^21^. Given, however, that clinical prediction is predominantly concerned with identifying individuals with a high-risk of future disease, one may rightly question the utility of prediction models for historical disease. Additionally, some previous PGS applications have not specified the time of the predictions, limiting the scores application in clinical practice, where clinical guidelines include management options linked to risk-thresholds across a specific follow-up time (often 10 years). Furthermore, many previous PGS studies have exclusively considered predicting CVD in disease-free “general population” samples similar to our “wo T2DM/CVD”. For example, Khera ^11^*et al* showed a c-statistic for coronary artery (CAD) and AF of 0.81 (95%CI 0.81; 0.81) and 0.77 (95%CI 0.76– 0.77) respectively. Contrary to our current manuscript, the models from Khera *et al* are problematic to apply out-of-sample because they were trained (and tested) on a combination of incident and prevalent disease, and additionally included study-specific predictors not available (or generalizable) to external data such, as genetic principal components and genotyping array. Inouye^22^ *et al* focussed on predicting incident CAD using a combination of seven known disease risk-factors and a CAD PGS to reach a c-statistic of 0.70 (95%CI 0.69; 0.70), which is slightly lower than the CHD c-statistic for our PGS-extended model CHD: 0.73 (95%CI 0.73; 0.73) in people without diabetes. Recent applications of PGS have followed this approach and developed model combining healthcare information with genetics, typically finding that PGS only moderately improves predictive performance in the general population ^23, 24, 25^. Here we show that PGS might provide much needed and unique information to improve predictive modelling for people with established disease such as diabetes.

By considering participants with diabetes stratified by their history of CVD at the enrolment we find that while the PGS was the most important features, the overall predictive performance was attenuated compared to participants without diabetes. The attenuated performance in diabetes participants was related to a decreased contribution of age to the overall c-statistic (Appendix Figure 19). For example, when predicting AF, permuting age resulted in a change in c-statistic of 0.12 for the “wo T2DM/CVD” group, compared to a more modest change of 0.06 for “w T2DM” and 0.01 for “w T2DM&CVD”; in contrast, the PGS was important in all three groups. Accounting for the duration of diabetes diagnoses, suggested that part of the attenuated performance compared to people without diabetes, might be due to inclusion of people with a prevalent diabetes diagnosis. Focussing instead on incident diabetes patients meaningfully improved performance, resulting in highly accurate models for CHD and HF (c-statistic above 0.80) for the “w T2DM” group.

We wish to acknowledge the follow study limitations. Firstly, some of the PGS partially included UKB data due to the cumulative nature of GWAS. Exploring the difference in c-statistic between weighted and unweighted scores, where the unweighted PGS are less susceptible to overfitting, suggesting this had limited influence (difference in c-statistic often below 0.01, Appendix Table 19). Secondly, comparing the UKB enrolled subjects to the general UK populations has revealed important differences, where UKB participants are generally healthier: they are less likely to be obese, to smoke or drink alcohol^26^. To further confirm a clinical utility of the derived models, it is therefore necessary to perform external validation studies. We note, however, that any model, irrespective of its derivation source, requires external validation. When considering deploying a model to *any* local setting, it is good practice to perform such validation studies using data representative of the intended population, which can also be used to recalibrate a model to local settings ^27^. Relatedly, our current paper exclusively focussed on performance in participants of European ancestry, reflecting the sampling design used by the available source GWAS. Thirdly, in this study we used a limited subset of clinical characteristics that might be relevant for CVD prediction. It is highly likely that including additional features may further optimize performance.

In summary, we have evaluated the added benefit of PGS scores to predict six types of CVD in participants without diabetes and CVD at baseline, compared to performance in participants with T2DM, and participants with T2DM and a history of CVD. In isolation, the PGS could only moderately predict incident disease.

Combining these scores with known clinical risk factors improved performance, showing relatively good discriminative ability especially for CHD, AF, and HF in people with T2DM but no history of CVD at enrolment. When considering only individuals with newly diagnosed diabetes, the discriminative ability of the models was substantially improved.

## Data Availability

All data produced in the present study are available upon reasonable request to the authors

## Author contributions

FWA, AFS, NC contributed to the idea and design of the study. CF and AFS implemented modules for genetic score calculation and biomarkers extraction from GP data. JG conducted genetic data quality control. CF supported the process of prescription extraction, and RM performed a manual review of lipid-lowering and antihypertensive prescriptions. MV reviewed and validated the computer application. KD prepared the dataset for analysis and implemented stacked genetic models and conventional risk prediction models. KD and AFS conducted the data analysis and created the figures. KD wrote the manuscript with support from FWA, AFS, CF, and NC. FWA, CF, NC, AFS, AH and JG provided critical feedback on the analysis and its interpretation, and commented on the drafted manuscript. KD is responsible for the integrity of the work as a whole.

## Code availability

Analyses were carried out in Python v3.6 using *scikit-learn*^28^, *statsmodels*^29^, *pandas*^30^, and *numpy*^31^, plots were generated using *matplotlib*^32^, and *seaborn*^33^. Imputation was performed in R v4.1, using *mice*^19^ .

To facilitate model deployment, we have prepared a straightforward computer application with detailed guidelines (https://gitlab.com/cvd_in_t2dm/prs_cvd_prediction).

## Acknowledgments

This research has been conducted using the UK Biobank Resource under application numbers 12113, 24711 and 44972. We are grateful to the UK Biobank participants. UK Biobank was established by the Wellcome Trust medical charity, Medical Research Council, Department of Health, Scottish Government, and the Northwest Regional Development Agency. It has also had funding from the Welsh Assembly Government and the British Heart Foundation.

## Prior postings and presentations

This study and its results have not been published previously. A preprint version has been deposited on medrxiv.

