## Appendix for "Combining stacked polygenic scores with clinical risk factors improves cardiovascular risk prediction in people with type 2 diabetes"

Dziopa et al

### Acronyms

**AF** Atrial fibrillation

**BMI** Body Mass Index

**CAD** Coronary artery disease

**CHD** Coronary heart disease

**CIL** Calibration-in-the-large

**cIMT** Carotid artery intima media thickness

**CKD** Chronic Kidney Disease

**CS** Calibration slope

**CVD+** Cardiovascular disease including heart failure and/or atrial fibrillation

**DBP** Diastolic blood pressure

**eGFR** Estimated glomerular filtration rate

**GLM** Generalized linear model

**HbA1c** Glycated haemoglobin

**HDL** High-density lipoprotein

**HF** Heart failure

**HOMA-B** Homeostasis Model Assessment of Beta-cell Function

**HOMA-IR** Homeostatic Model Assessment of Insulin Resistance

**IBD** Inflammatory bowel disease

**Is. Stroke** Ischaemic Stroke

**LDL** Low-density lipoprotein

**LR** Logistic Regression

**RF** Random Forest

**SBP** Systolic blood pressure

**T2DM** Type 2 diabetes

**w T2DM** Individuals with T2DM diagnosis but not history of CVD

**w T2DM&CVD** Individuals with T2DM and a history of CVD

**wo T2DM/CVD** General population without a history of CVD or T2DM

### Contents

**Methods** 8

**Results** 9

**Tables** 11

**Figures** 35

13 An exemplar classification tree from the 100 many trees that were used by the "PGS-only" model for 10 years CHD risk in participants with T2DM and a history of CVD. 47

14 The contribution of each of the 29 considered univariable polygenic scores to an overall stacked polygenic score (PGS-only). . . . . . . . . . . . . . . . . . . . . . . . . 48
15 Calibration plots for the PGS-only models stratified by outcome and participant group. 49
16 Probability distribution plots for the PGS-only models stratified by outcome, participant group, and case/control group. . . . . . . . . . . . . . . . . . . . . . . . . . 50
17 Calibration plots for the PGS-plus models stratified by outcome and participant group. 51
18 Probability distribution plots for the PGS-plus models stratified by outcome, participant group, and case/control group. . . . . . . . . . . . . . . . . . . . . . . . . . 52
19 The contribution of each clinical feature to the prediction of cardiovascular disease. . 53
20 Probability distribution plots for the PGS-extended models stratified by outcome, participant group, and case/control status. . . . . . . . . . . . . . . . . . . . . . . 54
21 Kaplan-Meier estimates of the 5-year cumulative incident of CVD after an enrolment date for the ”w T2DM” group, stratified by prevalent and incident type 2 diabetes. . . 55
22 Kaplan-Meier estimates of the 5-year cumulative incident of CVD after an enrolment date for the ”w T2DM&CVD” group, stratified by prevalent and incident type 2 diabetes. 56
23 The contribution of each feature (PRS, age, and sex) to the prediction of cardiovascular disease. . . . . . . . . . . . . . . . . . . . . . . . . . . . . . . . . . . . . 56
24 Calibration plots for the PGS-extended models stratified by outcome and participant group. . . . . . . . . . . . . . . . . . . . . . . . . . . . . . . . . . . . . . 57

### Methods

#### Data source

A CVD history and incident CVD were ascertained using the International Classification of Diseases ICD9-10, Read Codes v2 and Clinical Terms Version 3 (CTV3), and Office of Population Censuses and Surveys (OPCS) codes from Hospital Episode Statistics (HES) following the published CALIBER phenotyping algorithms [1].

#### Estimating model performance using independent testing data

In the feature importance procedure, iteratively the values of each variable was randomly assigned to an individual (permuted – thus breaking its association with the endpoint) after which the c-statistic was re-estimated with these permuted data and the difference in performance used as an estimate of the variables' contribution to the unpermuted c-statistic.

#### Sensitivity analyses

Due to the cumulative nature of most GWAS, some of the univariable PGS will have partially included UKB data (specifically here the PGS of AF, BMI, CAD in diabetes, CKD, HF, SBP, T2DM, and eGFR), which may have led to a degree of over optimization not alleviated by our strict separation of training and testing data. In most cases the UKB contributed less than 45% of the overall sample, limiting the degree of overfit (Appendix Table 7). Nevertheless, to explore this further each univariable PGS was derived weighted by the genetic association with the GWAS trait (e.g., by the log odd ratio for HF) as well as unweighted (i.e., setting the weights to 1). Given that the unweighted univariable scores use less information than their weighted counterparts, the difference in c-statistic was used as an indication of potential model overfitting.

### Results

#### Training 29 genetic scores to predict 10-years CVD risk

The PGS from a CAD GWAS conducted in T2DM showed the largest c-statistic (0.63) contribution for 10-year CVD+ risk in both “w T2DM” and “w T2DM&CVD”. In contrast, the polygenic score (PGS) from the AF GWAS showed the largest c-statistic for predicting CVD+ in “wo T2DM/CVD” participants (a c-statistic of 0.58), see Appendix Table 5 and Appendix Figure 5. Similar performance was observed for the 5 most of the remaining outcomes (Appendix Figure 5).

Examination of the parameter values selected by the trained models (Figure 1), found that most selected PGS utilized a p-value threshold lower than the conventional threshold of GWAS significance [2] (less than 20% of the PGS used a p-value of $5 \times 10^{-8}$), and showed a clear preference for approximately independent variants (selecting an r-squared of 0.20 or 0.01 at least 50% of the time). The choice of weighted or unweighted PGS (where the genetic variant was weighted by its GWAS effect size) was often close to 50%, with the PGS derived for the two disease groups showing a slight tendency for unweighted scores. The number of unique variants used by each PGS varied substantially; for example, in the “w T2DM&CVD” sample the HOMA-B PGS trained to predict AF consisted of only 8 variants, while the SBP PGS leveraged 23,034 variants for the same group and outcome (see Appendix Figure 6).

#### Stacking multiple PGS into multivariable prognostic models

First the pairwise correlation between each PGS was considered, and screened for potential multicollinearity (correlation coefficient of 0.90 or larger, to prevent model instability), which did not identify any highly correlated PGS in any of the three participant groups (Appendix Figures 7 - 9).

Through 10-fold cross-validation the PGS scores were stacked using four distinct learners (see methods), a random forest (RF) learner was often the top performing model, especially for the ”wo

T2DM/CVD" group (Appendix Figure 10, see example trees in Appendix Figures 11 – 13). Compared to the "wo T2DM/CVD" group, the stacked PGS-only models showed relatively good discrimination in the "w T2DM" and "w T2DM&CVD" test data for the CVD+, CVD, and CHD outcomes. For example, the c-statistic for CHD was 0.58 (95%CI 0.58; 0.58) for the "wo T2DM/CVD" compared to 0.70 (95%CI 0.70; 0.70) for "w T2DM" participants and 0.67 (95%CI 0.67; 0.67) for the "w T2DM&CVD" participants (Figure 2, Appendix Figures 14 - 16 and Appendix Table 8).

#### Sensitivity analyses

Some univariable PGS were derived from GWAS which included UKB data. While often the UKB constituted less than 45% of the overall data, this could potentially lead to an unknown degree of overfitting especially in PGS that were weighted by variant specific effect estimates (e.g., log odds ratios). To explore this, we compared the grid-search selected univariable PGS with its unweighted counterpart (which by not weighting by the variant effect estimate is less susceptible to overfitting). The difference in c-statistic between the selected univariable PGS and its unweighted counterpart was typically $< 0.01$, and of unlikely concern (Appendix Table 19).

To explore the influence of missing data on models including clinical characteristics such as blood pressure and BMI, we compared the discriminative ability under imputation and complete-case strategies, showing minimal differences (Appendix Tables 16, 20). Additionally, we explored performance of non-genetic model exclusively using age, sex and medicine prescriptions, which were available in all participants. Aside from a larger difference when predicting HF in "w T2DM&CVD" and "w T2DM" participants, the "Age & Sex & Clinical" models only showed a slightly better discriminative ability than the "Age & Sex & Medication" models for most of the endpoints (Figure 2).

### Tables

**Table 1:** Clinical characteristics and related UK field identifiers.

| **Clinical characteristics** | **Data Field Id** |
|---|---|
| Sex | 22001 |
| Smoking status | 20116 |
| BMI | 21001 |
| SBP | 4080, 93 |
| DBP | 4079 |
| LDL cholesterol | 23405 |
| HDL cholesterol | 30760 |
| Total cholesterol | 30690 |
| HbA1C | 30750 |

n.b. Considered clinical characteristics include: sex, smoking status, Body Mass Index (BMI), diastolic blood pressure (DBP), systolic blood pressure (SBP), low-density lipoprotein (LDL) cholesterol, high-density lipoprotein (HDL), total cholesterol, and glycated haemoglobin (HbA1C).

**Table 2:** Clinical codes used for patient characteristic extraction.

| Clinical characteristics | Clinical code | Reference |
|---|---|---|
| HbA1C | 42W4., 42W2., 42W1., 42W3., 42W5., 42W50, 42W51, 42W.., 44TB., 42WZ., 44TB0, 44TL., 44TB1, XaERp, 42W2., 42W1., 42W3., XaPbt, Xaeze, X772q, 42WZ., X80U3, XaJNg, XabrE, X80U4, XabrF | Free text searches through code mappings |
| BMI | 22K.., 22K1., 22K2., 22K3., 22K4., 22K5., 22K6., 22K7., 22K8., 22Z.., Y7DRh, Y7DRj, YMKUW, YamHz, YamI0 | Clinical Codes |
| HDL cholesterol | 44d2., 44dA., 44P5., 44PC. | Caliber HDL cholesterol |
| LDL cholesterol | 44d4., 44dB., 44P6., 44PE., 44PI. | Caliber LDL cholesterol |
| Total cholesterol | 44OE., 44P1., 44P2., 44P3., 44P4., 44P.., 44P9. | Caliber Total Cholesterol Plasma<br>Caliber Total Cholesterol Serum |
| SBP, DBP | X773t, X779R, XM02Y, X779T, X779U, X779W, 315B., XM02X, X779Q, X779S, X779V, X779S, 246N., 246P., 246Q., 246R., 246D., 246C., 246.., 246Z., 315B. | Free text searches through code mappings |
| Creatinine | 44J3., 44J30, 44J31, 44J32, 44J33, 44J3z, 44JC. | Caliber Creatinine |
| C-reactive protein | 44CC., 44CC0, 44CC1, 44CS. | Caliber C-reactive protein |

**Table 3:** Summary of applied cut-offs for the continuous variables.

| Clinical characteristics | Valid data ranges (applied cut-offs) |
|---|---|
| Age (years) | [18.0, 100.0] |
| HbA1c (mmol/mol)<br>HbA1c (%) | [20.0, 120.0]<br>[4.0, 13.1] |
| HDL cholesterol (mmol/L) | [0.0, 20.0] |
| Total cholesterol (mmol/L) | [0.0, 20.0] |
| SBP (mm Hg) | [70, 250] |
| BMI | [15, 55] |
| C-reactive protein (mg/L) | [8, 1000] |
| Serum creatinine (micromol/L) | [0.0 - 900] |

**Table 4:** UK Biobank data field identifiers and free-text fields used to identify diabetes medications relevant for the phenotyping algorithm detailed in Figure 1.

| UK Biobank Data field | Diabetes medication |
|---|---|
| 20003 | Insulin, insulin product, Metformin, metformin<br>glucophage 500mg tablet, orabet 500mg tablet, Repaglinide<br>novonorm, repaglinide, Sulfonylureas, glibenclamide,<br>glibenese 5mg tablet, daonil 5mg tablet, semi-daonil 2.5mg tablet<br>diabetamide 2.5mg tablet, euglucon 2.5mg tablet, malix 2.5mg tablet<br>calabren 2.5mg tablet, gliclazide, diamicron 80mg tablet<br>chlorpropamide, diabinese 100mg tablet, glymese 250mg tablet<br>tolbutamide, glyconon 500mg tablet, rastinon 500mg tablet,<br>glymidine, gondafon 500mg tablet, pramidex 500mg tablet<br>tolazamide, tolanase 100mg tablet, glipizide product<br>glipizide, glibenese 5mg tablet, minodiab 2.5mg tablet<br>gliquidone, glurenorm 30mg tablet, glimepiride<br>amaryl 1mg tablet, Alpha glucosidase inhibitors<br>acarbose, glucobay 50mg tablet, Thiazolidinediones<br>troglitazone, glutril 25mg tablet, romozin 200mg tablet,<br>rosiglitazone 1mg / metformin 500mg tablet, rosiglitazone<br>avandia 4mg tablet, avandamet 1mg / 500mg tablet, pioglitazone<br>actos 15mg tablet, Nateglinide, nateglinide<br>starlix 60mg tablet |
| 6177 | Insulin |
| 6153 | Insulin |

**Table 5:** Genome wide association studies (GWAS) used to construct polygenic scores for the prediction of cardiovascular disease risk.

| **Phenotype** | **GWAS**<br>Website reference | **Used UKB data** |
|---|---|---|
| Atrial fibrillation (AF) | `https://www.nature.com/articles/s41588-018-0171-3` | yes |
| Arthritis | `https://www.ncbi.nlm.nih.gov/pubmed/?term=24390342` | no |
| Body Mass Index (BMI) | `https://pubmed.ncbi.nlm.nih.gov/30124842/` | yes |
| Coronary artery disease (CAD) in diabetes | `https://link.springer.com/article/10.1007%2Fs00125-018-4686-z` | yes |
| Coronary heart disease (CHD) | `https://www.nature.com/articles/ng.3396` | no |
| Chronic Kidney Disease (CKD) | `https://www.ncbi.nlm.nih.gov/pmc/articles/PMC6698888/` | yes |
| Cardioembolic stroke | `http://www.megastroke.org/index.html` | no |
| Crohn's disease | `https://www.nature.com/articles/ng.717` | no |
| Female Fasting Insulin, Male Fasting Insulin | `https://www.magicinvestigators.org/downloads/` | no |
| Female fasting glucose, Male fasting glucose | `https://www.magicinvestigators.org/downloads/` | no |
| HDL cholesterol | `https://www.nature.com/articles/ng.2797` | no |
| Heart Failure (HF) | `https://www.nature.com/articles/s41467-019-13690-5` | yes |
| Homeostasis Model Assessment of Beta-cell Function (HOMA-B) | `https://www.magicinvestigators.org/downloads/` | no |
| Homeostatic Model Assessment of Insulin Resistance (HOMA-IR) | https://www.magicinvestigators.org/downloads/ | no |
| Glycated haemoglobin (HbA1c) | `https://pubmed.ncbi.nlm.nih.gov/28898252/` | no |
| Inflammatory bowel disease (IBD) | `https://www.ncbi.nlm.nih.gov/pubmed/23128233` | no |
| Ischaemic Stroke (Is. Stroke) | `http://www.megastroke.org/index.html` | no |
| LDL cholesterol | `https://www.nature.com/articles/ng.2797` | no |
| Large artery stroke | `http://www.megastroke.org/index.html` | no |
| Systolic blood pressure (SBP) | `https://www.nature.com/articles/s41588-018-0205-x` | yes |
| Small vessel stroke | `http://europepmc.org/abstract/MED/20858683` | no |
| Stroke | `http://www.megastroke.org/index.html` | no |
| Type 2 diabetes (T2DM) | `https://www.nature.com/articles/s41588-018-0241-6` | yes |
| Triglycerides | `https://www.nature.com/articles/ng.2797` | no |
| Ulcerative colitis | `https://www.nature.com/articles/ng.764` | no |
| Carotid artery intima media thickness (cIMT) | `https://www.nature.com/articles/s41467-018-07340-5` | no |
| Estimated glomerular filtration rate (eGFR) | `https://www.ncbi.nlm.nih.gov/pmc/articles/PMC6698888/` | yes |

**Table 6:** Random forest (RF) grid search hyperparameters.

| Hyperparameter | Hyperparameter role | Explored values |
|---|---|---|
| `max_depth` | the longest path between the root node and the leaf node | 3, 4, 5 |
| `min_samples_leaf` | the minimum number of samples that should be present in the leaf node after splitting a node | 50, 100, 150, 200 |
| `min_samples_split` | the minimum required number of observations in any given node in order to split it | 50, 70, 90, 100 |
| `n_estimators` | the number of trees to be considered | 50, 100, 150, 200 |
| `max_samples` | determines what fraction of the original dataset is given to any individual tree | 0.2, 0.5, 0.7, 0.99 |
| `max_features` | the number of maximum features provided to each tree in a random forest. | 5, 10, 15, 20, 25 |

**Table 7:** The proportion of UK Biobank cases that were used for constructing polygenic scores for the prediction of cardiovascular disease risk.

| Phenotype | Number of UK Biobank cases | Number of total cases | (%) |
|---|---|---|---|
| Atrial fibrillation (AF) | 395,739 | 1,030,836 | 38.4 |
| BMI | 450,000 | 700,000 | 64.3 |
| Coronary artery disease (CAD) in diabetes | 3,968 | 3,968 | 100.0 |
| Chronic Kidney Disease (CKD) | 452,264 | 1,046,070 | 43.2 |
| Heart Failure (HF) | 394,156 | 977,323 | 40.3 |
| Systolic blood pressure (SBP) | 458,577 | 757,601 | 60.5 |
| Type 2 diabetes (T2DM) | 19,119 | 74,124 | 25.8 |
| Estimated glomerular filtration rate (eGFR) | 452,264 | 1,046,070 | 43.2 |

**Table 8:** Discriminative performance of stacked polygenic score (PGS) models combining up to 29 GWAS specific PGS to predict cardiovascular disease.

| Outcome | Group | C-statistic (95%CI) | CS (95%CI) | CIL (95%CI) |
|---|---|---|---|---|
| CVD+ | wo T2DM/CVD | 0.581 (0.581; 0.582) | 1.088 (0.947; 1.229) | 0.125 (0.086; 0.165) |
| CVD+ | w T2DM | 0.635 (0.634; 0.636) | 3.812 (2.563; 5.061) | 0.308 (0.160; 0.456) |
| CVD+ | w T2DM & CVD | 0.586 (0.583; 0.588) | 8.548 (1.267; 15.828) | 1.589 (1.326; 1.852) |
| CVD | wo T2DM/CVD | 0.569 (0.568; 0.569) | 1.284 (1.032; 1.535) | 0.119 (0.071; 0.167) |
| CVD | w T2DM | 0.661 (0.659; 0.662) | 1.021 (0.732; 1.311) | 0.029 (-0.144; 0.202) |
| CVD | w T2DM & CVD | 0.619 (0.617; 0.621) | 0.909 (0.370; 1.449) | -0.073 (-0.322; 0.175) |
| CHD | wo T2DM/CVD | 0.577 (0.576; 0.577) | 1.293 (1.034; 1.552) | 0.106 (0.052; 0.160) |
| CHD | w T2DM | 0.702 (0.700; 0.704) | 1.185 (0.903; 1.467) | 0.182 (-0.012; 0.377) |
| CHD | w T2DM & CVD | 0.669 (0.667; 0.671) | 2.058 (1.160; 2.957) | 1.213 (0.981; 1.444) |
| HF | wo T2DM/CVD | 0.592 (0.591; 0.593) | 0.760 (0.504; 1.015) | -0.043 (-0.154; 0.068) |
| HF | w T2DM | 0.640 (0.636; 0.643) | 0.511 (0.171; 0.851) | 0.053 (-0.273; 0.379) |
| HF | w T2DM & CVD | 0.580 (0.577; 0.582) | 0.801 (0.086; 1.516) | 0.021 (-0.235; 0.276) |
| AF | wo T2DM/CVD | 0.643 (0.643; 0.644) | 0.949 (0.851; 1.046) | 0.056 (-0.000; 0.113) |
| AF | w T2DM | 0.660 (0.658; 0.662) | 1.366 (0.869; 1.862) | 0.136 (-0.075; 0.348) |
| AF | w T2DM & CVD | 0.596 (0.594; 0.598) | 0.961 (0.265; 1.657) | 0.289 (0.045; 0.533) |
| Ischaemic Stroke | wo T2DM/CVD | 0.557 (0.556; 0.558) | 1.034 (0.543; 1.524) | 0.107 (-0.002; 0.216) |
| Ischaemic Stroke | w T2DM | 0.601 (0.598; 0.605) | 0.413 (0.011; 0.815) | 0.184 (-0.191; 0.559) |
| Ischaemic Stroke | w T2DM & CVD | 0.464 (0.461; 0.468) | -0.110 (-0.584; 0.364) | -0.011 (-0.393; 0.372) |

Individuals are stratified as followed ”wo T2DM/CVD”: participants without T2DM or CVD at baseline, “w T2DM”: participants with diabetes at baseline, “w T2DM&CVD”: participants with T2DM at baseline and a history of CVD. The analysed outcomes include cardiovascular disease including heart failure (HF) and/or atrial fibrillation (AF) (CVD+), cardiovascular disease (CVD), coronary heart disease (CHD), HF, AF, and Ischaemic Stroke. Discrimination (c-statistic) and calibration (calibration-in-the-large (CIL) and calibration slope (CS)) are based on a 20% of test set of the total data used for this study. Point estimates are presented alongside 95% CI.

**Table 9:** Performance of stacked polygenic score (PGS) models combining up to 29 GWAS specific PGS with age and sex to predict cardiovascular disease.

| Outcome | Group | C-statistic (95%CI) | CS (95%CI) | CIL (95%CI) |
|---|---|---|---|---|
| CVD+ | wo T2DM/CVD | 0.711 (0.711; 0.711) | 1.029 (0.970; 1.087) | 0.176 (0.136; 0.217) |
| CVD+ | w T2DM | 0.690 (0.689; 0.691) | 1.149 (0.878; 1.420) | 0.367 (0.214; 0.519) |
| CVD+ | w T2DM & CVD | 0.632 (0.630; 0.635) | 1.621 (0.670; 2.573) | 1.639 (1.375; 1.904) |
| CVD | wo T2DM/CVD | 0.696 (0.695; 0.696) | 0.931 (0.864; 0.998) | 0.156 (0.107; 0.205) |
| CVD | w T2DM | 0.698 (0.697; 0.700) | 1.153 (0.870; 1.436) | 0.228 (0.055; 0.402) |
| CVD | w T2DM & CVD | 0.661 (0.659; 0.663) | 2.428 (1.386; 3.469) | 1.392 (1.148; 1.637) |
| CHD | wo T2DM/CVD | 0.701 (0.700; 0.701) | 0.901 (0.830; 0.972) | 0.140 (0.086; 0.195) |
| CHD | w T2DM | 0.736 (0.734; 0.737) | 1.143 (0.889; 1.397) | 0.170 (-0.026; 0.366) |
| CHD | w T2DM & CVD | 0.704 (0.702; 0.706) | 2.156 (1.300; 3.012) | 1.214 (0.981; 1.447) |
| HF | wo T2DM/CVD | 0.724 (0.723; 0.725) | 0.708 (0.608; 0.809) | -0.036 (-0.148; 0.077) |
| HF | w T2DM | 0.695 (0.692; 0.698) | 0.703 (0.368; 1.039) | 0.072 (-0.255; 0.398) |
| HF | w T2DM & CVD | 0.620 (0.617; 0.622) | 0.695 (0.245; 1.145) | 0.313 (0.053; 0.572) |
| AF | wo T2DM/CVD | 0.757 (0.757; 0.758) | 0.940 (0.880; 1.000) | 0.103 (0.045; 0.161) |
| AF | w T2DM | 0.726 (0.724; 0.728) | 0.967 (0.711; 1.222) | 0.222 (0.004; 0.440) |
| AF | w T2DM & CVD | 0.604 (0.602; 0.606) | 0.478 (0.153; 0.802) | 0.356 (0.104; 0.609) |
| Ischaemic Stroke | wo T2DM/CVD | 0.674 (0.673; 0.675) | 0.513 (0.423; 0.602) | 0.147 (0.037; 0.257) |
| Ischaemic Stroke | w T2DM | 0.602 (0.598; 0.605) | 0.411 (0.042; 0.781) | 0.197 (-0.179; 0.573) |
| Ischaemic Stroke | w T2DM & CVD | 0.464 (0.461; 0.468) | -0.177 (-0.903; 0.550) | -0.189 (-0.567; 0.189) |

Individuals are stratified as followed ”wo T2DM/CVD”: participants without T2DM or CVD at baseline, “w T2DM”: participants with diabetes at baseline, “w T2DM&CVD”: participants with T2DM at baseline and a history of CVD. The analysed outcomes include cardiovascular disease including heart failure (HF) and/or atrial fibrillation (AF) (CVD+), cardiovascular disease (CVD), coronary heart disease (CHD), HF, AF, and Ischaemic Stroke. Discrimination (c-statistic) and calibration (calibration-in-the-large (CIL) and calibration slope (CS)) are based on a 20% of test set of the total data used for this study. Point estimates are presented alongside 95% CI.

**Table 10:** Performance of stacked polygenic score (PGS) models combining up to 29 GWAS specific PGS with clinical characteristics to predict cardiovascular disease.

| Outcome | Group | C-statistic (95%CI) | CS (95%CI) | CIL (95%CI) |
|---|---|---|---|---|
| CVD+ | wo T2DM/CVD | 0.730 (0.730; 0.730) | 1.065 (1.010; 1.121) | 0.187 (0.146; 0.228) |
| CVD+ | w T2DM | 0.719 (0.718; 0.721) | 1.157 (0.915; 1.400) | 0.417 (0.263; 0.571) |
| CVD+ | w T2DM & CVD | 0.623 (0.621; 0.625) | 1.849 (0.718; 2.981) | 1.647 (1.383; 1.911) |
| CVD | wo T2DM/CVD | 0.721 (0.720; 0.721) | 0.982 (0.918; 1.045) | 0.166 (0.117; 0.215) |
| CVD | w T2DM | 0.716 (0.714; 0.717) | 1.060 (0.816; 1.304) | 0.292 (0.117; 0.467) |
| CVD | w T2DM & CVD | 0.663 (0.661; 0.665) | 2.832 (1.627; 4.038) | 1.406 (1.161; 1.650) |
| CHD | wo T2DM/CVD | 0.728 (0.727; 0.728) | 0.960 (0.893; 1.028) | 0.148 (0.093; 0.203) |
| CHD | w T2DM | 0.749 (0.748; 0.751) | 1.158 (0.907; 1.410) | 0.180 (-0.017; 0.376) |
| CHD | w T2DM & CVD | 0.720 (0.718; 0.722) | 2.337 (1.515; 3.158) | 1.223 (0.990; 1.457) |
| HF | wo T2DM/CVD | 0.743 (0.742; 0.744) | 0.751 (0.654; 0.849) | -0.031 (-0.144; 0.081) |
| HF | w T2DM | 0.713 (0.710; 0.716) | 0.671 (0.379; 0.962) | 0.103 (-0.227; 0.432) |
| HF | w T2DM & CVD | 0.642 (0.640; 0.645) | 0.759 (0.366; 1.153) | 0.328 (0.065; 0.590) |
| AF | wo T2DM/CVD | 0.770 (0.770; 0.771) | 0.966 (0.908; 1.025) | 0.111 (0.053; 0.169) |
| AF | w T2DM | 0.762 (0.760; 0.763) | 1.066 (0.812; 1.320) | 0.221 (0.002; 0.440) |
| AF | w T2DM & CVD | 0.605 (0.602; 0.607) | 0.599 (0.195; 1.004) | 0.261 (0.012; 0.510) |
| Ischaemic Stroke | wo T2DM/CVD | 0.684 (0.683; 0.685) | 0.536 (0.448; 0.623) | 0.155 (0.044; 0.265) |
| Ischaemic Stroke | w T2DM | 0.632 (0.628; 0.635) | 0.454 (0.115; 0.794) | 0.248 (-0.130; 0.625) |
| Ischaemic Stroke | w T2DM & CVD | 0.465 (0.462; 0.469) | -0.124 (-0.836; 0.589) | -0.186 (-0.565; 0.192) |

Individuals are stratified as followed ”wo T2DM/CVD”: participants without T2DM or CVD at baseline, “w T2DM”: participants with diabetes at baseline, “w T2DM&CVD”: participants with T2DM at baseline and a history of CVD. The analysed outcomes include cardiovascular disease including heart failure (HF) and/or atrial fibrillation (AF) (CVD+), cardiovascular disease (CVD), coronary heart disease (CHD), HF, AF, and Ischaemic Stroke. Discrimination (c-statistic) and calibration (calibration-in-the-large (CIL) and calibration slope (CS)) are based on a 20% of test set of the total data used for this study. Point estimates are presented alongside 95% confidence interval (CI).

**Table 11:** The univariable odds ratio for the predictor association with the cardiovascular outcome in the ”wo T2DM/CVD” study cohort.

| Predictor | CVD+ (95%CI) | CVD (95%CI) | CHD (95%CI) | HF (95%CI) | AF (95%CI) | Ischaemic Stroke (95%CI) |
|---|---|---|---|---|---|---|
| PGS (SD) | 1.45 (1.43; 1.48) | 1.51 (1.47; 1.54) | 1.59 (1.55; 1.63) | 2.03 (1.95; 2.12) | 1.93 (1.88; 1.98) | 2.55 (2.42; 2.68) |
| Sex (Male vs Female) | 1.98 (1.90; 2.06) | 1.99 (1.89; 2.09) | 2.15 (2.03; 2.28) | 2.01 (1.80; 2.25) | 2.22 (2.10; 2.35) | 1.66 (1.48; 1.86) |
| Age (years) | 1.09 (1.09; 1.10) | 1.08 (1.08; 1.09) | 1.08 (1.07; 1.08) | 1.11 (1.10; 1.12) | 1.11 (1.11; 1.12) | 1.10 (1.09; 1.11) |
| Smoking status (Previous vs Never) | 1.46 (1.40; 1.52) | 1.46 (1.38; 1.54) | 1.42 (1.34; 1.51) | 1.49 (1.32; 1.67) | 1.46 (1.38; 1.55) | 1.36 (1.20; 1.54) |
| Smoking status (Current vs Never) | 1.68 (1.57; 1.79) | 2.07 (1.92; 2.22) | 1.79 (1.64; 1.94) | 1.91 (1.63; 2.25) | 1.11 (1.00; 1.23) | 1.90 (1.61; 2.25) |
| BMI (kg/m2) | 1.05 (1.04; 1.05) | 1.04 (1.04; 1.05) | 1.05 (1.04; 1.06) | 1.07 (1.06; 1.08) | 1.05 (1.04; 1.06) | 1.03 (1.01; 1.04) |
| SBP (mm Hg) | 1.02 (1.02; 1.02) | 1.02 (1.02; 1.02) | 1.02 (1.02; 1.02) | 1.02 (1.02; 1.02) | 1.01 (1.01; 1.02) | 1.02 (1.02; 1.02) |
| DBP (mm Hg) | 1.01 (1.01; 1.02) | 1.02 (1.02; 1.02) | 1.02 (1.02; 1.02) | 1.02 (1.01; 1.02) | 1.01 (1.01; 1.01) | 1.02 (1.02; 1.03) |
| HDL cholesterol (mmol/L) | 0.67 (0.64; 0.71) | 0.63 (0.59; 0.67) | 0.58 (0.55; 0.62) | 0.66 (0.58; 0.75) | 0.70 (0.66; 0.75) | 0.80 (0.69; 0.91) |
| Total cholesterol (mmol/L) | 0.93 (0.91; 0.95) | 0.99 (0.97; 1.01) | 1.00 (0.97; 1.02) | 0.89 (0.84; 0.93) | 0.85 (0.83; 0.87) | 0.96 (0.91; 1.01) |
| HbA1C (mmol/mol) | 1.04 (1.04; 1.05) | 1.05 (1.04; 1.05) | 1.04 (1.04; 1.05) | 1.05 (1.04; 1.05) | 1.03 (1.03; 1.04) | 1.04 (1.03; 1.05) |
| Antihypertensive medication | 2.36 (2.25; 2.48) | 2.13 (2.01; 2.25) | 2.16 (2.03; 2.31) | 2.28 (2.02; 2.57) | 2.64 (2.48; 2.81) | 1.99 (1.74; 2.28) |
| Lipid lowering medication | 2.38 (2.25; 2.52) | 2.36 (2.21; 2.52) | 2.46 (2.28; 2.64) | 2.10 (1.82; 2.43) | 2.31 (2.14; 2.49) | 2.11 (1.81; 2.46) |

The analysed outcomes include cardiovascular disease including heart failure (HF) and/or atrial fibrillation (AF) (CVD+), cardiovascular disease (CVD), coronary heart disease (CHD), HF, AF, and Ischaemic Stroke. Odds ratio estimates are presented alongside 95% confidence interval (CI).

**Table 12:** The univariable odds ratio for the predictor association with the cardiovascular outcome in the ”w T2DM” study cohort.

| Predictor | CVD+ (95%CI) | CVD (95%CI) | CHD (95%CI) | HF (95%CI) | AF (95%CI) | Ischaemic Stroke (95%CI) |
|---|---|---|---|---|---|---|
| PGS (SD) | 2.11 (1.94; 2.30) | 2.23 (2.03; 2.44) | 2.53 (2.31; 2.78) | 2.89 (2.41; 3.47) | 2.39 (2.13; 2.69) | 2.65 (2.14; 3.27) |
| Sex (Male vs Female) | 1.47 (1.25; 1.72) | 1.38 (1.15; 1.64) | 1.42 (1.16; 1.74) | 1.49 (1.05; 2.09) | 1.92 (1.51; 2.43) | 1.64 (1.06; 2.51) |
| Age (years) | 1.05 (1.04; 1.07) | 1.04 (1.03; 1.05) | 1.03 (1.02; 1.05) | 1.07 (1.04; 1.10) | 1.07 (1.05; 1.09) | 1.04 (1.01; 1.08) |
| Smoking status (Previous vs Never) | 1.40 (1.19; 1.65) | 1.41 (1.18; 1.70) | 1.43 (1.17; 1.75) | 1.33 (0.95; 1.86) | 1.38 (1.10; 1.73) | 1.21 (0.80; 1.84) |
| Smoking status (Current vs Never) | 1.45 (1.11; 1.88) | 1.51 (1.13; 2.02) | 1.30 (0.93; 1.81) | 0.99 (0.54; 1.82) | 1.40 (0.97; 2.01) | 1.53 (0.81; 2.88) |
| BMI (kg/m2) | 1.03 (1.02; 1.04) | 1.02 (1.01; 1.04) | 1.04 (1.02; 1.05) | 1.07 (1.05; 1.10) | 1.04 (1.03; 1.06) | 0.98 (0.95; 1.02) |
| SBP (mm Hg) | 1.01 (1.01; 1.01) | 1.01 (1.00; 1.01) | 1.01 (1.00; 1.01) | 1.02 (1.01; 1.03) | 1.01 (1.00; 1.02) | 1.01 (1.00; 1.02) |
| DBP (mm Hg) | 1.00 (0.99; 1.01) | 1.00 (0.99; 1.01) | 1.00 (0.99; 1.01) | 1.01 (0.99; 1.03) | 1.00 (0.99; 1.01) | 1.01 (0.99; 1.03) |
| LDL cholesterol (mmol/L) | 1.07 (0.98; 1.17) | 1.03 (0.93; 1.13) | 1.06 (0.94; 1.18) | 1.11 (0.92; 1.34) | 1.01 (0.89; 1.14) | 1.06 (0.84; 1.34) |
| HDL cholesterol (mmol/L) | 0.86 (0.74; 1.00) | 0.86 (0.72; 1.02) | 0.78 (0.64; 0.94) | 0.91 (0.66; 1.25) | 0.75 (0.61; 0.93) | 0.82 (0.56; 1.22) |
| Total cholesterol (mmol/L) | 1.01 (0.94; 1.08) | 0.98 (0.91; 1.06) | 0.99 (0.91; 1.08) | 1.02 (0.88; 1.18) | 0.96 (0.87; 1.06) | 0.96 (0.80; 1.16) |
| HbA1C (mmol/mol) | 1.01 (1.00; 1.01) | 1.01 (1.00; 1.02) | 1.00 (1.00; 1.01) | 1.01 (1.00; 1.03) | 1.00 (0.99; 1.01) | 1.02 (1.01; 1.03) |
| Antihypertensive medication | 1.61 (1.38; 1.88) | 1.65 (1.39; 1.96) | 1.93 (1.58; 2.34) | 1.80 (1.30; 2.51) | 1.59 (1.28; 1.97) | 1.20 (0.81; 1.78) |
| Lipid lowering medication | 1.08 (0.92; 1.26) | 1.27 (1.06; 1.51) | 1.38 (1.13; 1.68) | 0.91 (0.66; 1.26) | 0.95 (0.77; 1.18) | 1.16 (0.77; 1.74) |

The analysed outcomes include cardiovascular disease including heart failure (HF) and/or atrial fibrillation (AF) (CVD+), cardiovascular disease (CVD), coronary heart disease (CHD), HF, AF, and Ischaemic Stroke. Odds ratio estimates are presented alongside 95% confidence interval (CI).

**Table 13:** The univariable odds ratio for the predictor association with the cardiovascular outcome in the ”w T2DM&CVD” study cohort.

| Predictor | CVD+ (95%CI) | CVD (95%CI) | CHD (95%CI) | HF (95%CI) | AF (95%CI) | Ischaemic Stroke (95%CI) |
|---|---|---|---|---|---|---|
| PGS (SD) | 2.63 (2.23; 3.09) | 2.35 (2.03; 2.73) | 2.35 (2.04; 2.70) | 2.12 (1.83; 2.46) | 3.02 (2.57; 3.55) | 2.44 (1.96; 3.02) |
| Sex (Male vs Female) | 1.50 (1.11; 2.03) | 1.71 (1.29; 2.26) | 1.66 (1.27; 2.17) | 1.44 (1.04; 1.99) | 1.52 (1.12; 2.07) | 1.03 (0.67; 1.57) |
| Age (years) | 1.02 (1.00; 1.05) | 1.01 (0.99; 1.04) | 1.01 (0.99; 1.03) | 1.05 (1.02; 1.08) | 1.05 (1.03; 1.08) | 1.04 (1.00; 1.08) |
| Smoking status (Previous vs Never) | 1.63 (1.21; 2.20) | 1.67 (1.26; 2.21) | 1.54 (1.18; 2.00) | 1.30 (0.96; 1.77) | 1.18 (0.89; 1.56) | 1.67 (1.05; 2.65) |
| Smoking status (Current vs Never) | 1.31 (0.85; 2.03) | 1.35 (0.90; 2.03) | 1.04 (0.71; 1.51) | 1.30 (0.85; 2.01) | 1.20 (0.80; 1.80) | 2.66 (1.50; 4.72) |
| BMI (kg/m2) | 1.02 (1.00; 1.05) | 1.02 (0.99; 1.04) | 1.02 (1.00; 1.05) | 1.05 (1.02; 1.07) | 1.03 (1.01; 1.05) | 1.02 (0.98; 1.05) |
| SBP (mm Hg) | 1.00 (0.99; 1.00) | 1.00 (0.99; 1.00) | 0.99 (0.99; 1.00) | 0.99 (0.99; 1.00) | 1.00 (1.00; 1.01) | 1.01 (1.00; 1.02) |
| DBP (mm Hg) | 0.98 (0.97; 0.99) | 0.98 (0.97; 0.99) | 0.98 (0.97; 0.99) | 1.00 (0.98; 1.01) | 1.01 (1.00; 1.02) | 1.01 (0.99; 1.03) |
| LDL cholesterol (mmol/L) | 0.94 (0.78; 1.12) | 0.91 (0.77; 1.07) | 0.89 (0.76; 1.04) | 0.84 (0.71; 1.00) | 1.06 (0.90; 1.24) | 0.99 (0.78; 1.25) |
| HDL cholesterol (mmol/L) | 0.83 (0.64; 1.08) | 0.86 (0.67; 1.09) | 0.90 (0.72; 1.13) | 0.74 (0.58; 0.95) | 0.89 (0.70; 1.12) | 1.28 (0.90; 1.82) |
| Total cholesterol (mmol/L) | 0.84 (0.74; 0.95) | 0.84 (0.74; 0.95) | 0.85 (0.76; 0.96) | 0.85 (0.75; 0.97) | 0.99 (0.87; 1.11) | 0.94 (0.78; 1.12) |
| HbA1C (mmol/mol) | 1.01 (1.00; 1.02) | 1.01 (1.00; 1.02) | 1.01 (1.00; 1.02) | 1.02 (1.01; 1.03) | 1.01 (1.00; 1.02) | 1.02 (1.01; 1.03) |
| Antihypertensive medication | 1.64 (1.23; 2.17) | 1.52 (1.16; 1.98) | 1.47 (1.14; 1.89) | 1.29 (0.97; 1.73) | 1.32 (1.00; 1.74) | 0.99 (0.67; 1.47) |
| Lipid lowering medication | 1.47 (1.11; 1.97) | 1.51 (1.15; 1.98) | 1.44 (1.11; 1.86) | 1.14 (0.85; 1.52) | 1.10 (0.83; 1.44) | 0.85 (0.58; 1.26) |

The analysed outcomes include cardiovascular disease including heart failure (HF) and/or atrial fibrillation (AF) (CVD+), cardiovascular disease (CVD), coronary heart disease (CHD), HF, AF, and Ischaemic Stroke. Odds ratio estimates are presented alongside 95% confidence interval (CI).

**Table 14:** Performance of Age & Sex models trained on the imputed dataset to predict cardiovascular disease.

| Outcome | Group | C-statistic (95%CI) | CS (95%CI) | CIL (95%CI) |
|---|---|---|---|---|
| CVD+ | wo T2DM/CVD | 0.695 (0.694; 0.695) | 1.074 (1.007; 1.140) | 0.160 (0.119; 0.200) |
| CVD+ | w T2DM | 0.645 (0.644; 0.647) | 1.573 (1.070; 2.077) | 0.340 (0.191; 0.489) |
| CVD+ | w T2DM & CVD | 0.618 (0.616; 0.621) | 10.530 (5.212; 15.849) | 1.582 (1.319; 1.844) |
| CVD | wo T2DM/CVD | 0.685 (0.684; 0.685) | 1.056 (0.974; 1.138) | 0.140 (0.091; 0.188) |
| CVD | w T2DM | 0.624 (0.623; 0.626) | 1.653 (0.948; 2.358) | 0.198 (0.029; 0.367) |
| CVD | w T2DM & CVD | 0.616 (0.614; 0.618) | 6.877 (3.313; 10.442) | 1.355 (1.112; 1.599) |
| CHD | wo T2DM/CVD | 0.689 (0.689; 0.690) | 1.064 (0.973; 1.155) | 0.120 (0.066; 0.175) |
| CHD | w T2DM | 0.619 (0.617; 0.620) | 1.556 (0.762; 2.349) | 0.138 (-0.051; 0.328) |
| CHD | w T2DM & CVD | 0.620 (0.618; 0.622) | 5.809 (2.808; 8.810) | 1.174 (0.944; 1.404) |
| HF | wo T2DM/CVD | 0.718 (0.717; 0.719) | 0.970 (0.818; 1.122) | -0.041 (-0.152; 0.071) |
| HF | w T2DM | 0.672 (0.669; 0.675) | 1.325 (0.514; 2.136) | 0.044 (-0.277; 0.365) |
| HF | w T2DM & CVD | 0.592 (0.589; 0.594) | 1.634 (0.414; 2.854) | 0.282 (0.029; 0.536) |
| AF | wo T2DM/CVD | 0.719 (0.718; 0.719) | 0.993 (0.915; 1.071) | 0.072 (0.015; 0.129) |
| AF | w T2DM | 0.686 (0.684; 0.688) | 1.414 (0.909; 1.919) | 0.140 (-0.072; 0.352) |
| AF | w T2DM & CVD | 0.555 (0.552; 0.557) | 0.875 (-0.107; 1.857) | 0.306 (0.064; 0.548) |
| Ischaemic Stroke | wo T2DM/CVD | 0.702 (0.701; 0.703) | 1.017 (0.849; 1.184) | 0.110 (0.001; 0.220) |
| Ischaemic Stroke | w T2DM | 0.565 (0.562; 0.569) | 0.567 (-0.448; 1.581) | 0.154 (-0.216; 0.524) |
| Ischaemic Stroke | w T2DM & CVD | 0.544 (0.541; 0.548) | -0.137 (-2.227; 1.953) | -0.032 (-0.407; 0.343) |

Individuals are stratified as followed ”wo T2DM/CVD”: participants without T2DM or CVD at baseline, “w T2DM”: participants with diabetes at baseline, “w T2DM&CVD”: participants with T2DM at baseline and a history of CVD. The analysed outcomes include cardiovascular disease including heart failure (HF) and/or atrial fibrillation (AF) (CVD+), cardiovascular disease (CVD), coronary heart disease (CHD), HF, AF, and Ischaemic Stroke. Discrimination (c-statistic) and calibration (calibration-in-the-large (CIL) and calibration slope (CS)) are based on a 20% of test set of the total data used for this study. Point estimates are presented alongside 95% CI.

**Table 15:** Performance of Age & Sex & Medication models to predict cardiovascular disease.

| Outcome | Group | C-statistic (95%CI) | CS (95%CI) | CIL (95%CI) |
|---|---|---|---|---|
| CVD+ | wo T2DM/CVD | 0.705 (0.704; 0.705) | 1.102 (1.038; 1.167) | 0.162 (0.121; 0.202) |
| CVD+ | w T2DM | 0.649 (0.647; 0.650) | 1.457 (1.015; 1.900) | 0.345 (0.196; 0.494) |
| CVD+ | w T2DM & CVD | 0.633 (0.631; 0.635) | 8.718 (4.536; 12.900) | 1.586 (1.323; 1.849) |
| CVD | wo T2DM/CVD | 0.693 (0.693; 0.693) | 1.079 (1.000; 1.158) | 0.139 (0.090; 0.187) |
| CVD | w T2DM | 0.596 (0.594; 0.597) | 1.381 (0.670; 2.092) | 0.165 (-0.004; 0.334) |
| CVD | w T2DM & CVD | 0.639 (0.637; 0.641) | 10.672 (5.473; 15.872) | 1.376 (1.133; 1.620) |
| CHD | wo T2DM/CVD | 0.700 (0.699; 0.700) | 1.084 (0.998; 1.169) | 0.123 (0.068; 0.177) |
| CHD | w T2DM | 0.575 (0.573; 0.577) | 0.927 (0.234; 1.621) | 0.098 (-0.092; 0.287) |
| CHD | w T2DM & CVD | 0.645 (0.643; 0.647) | 9.394 (5.109; 13.679) | 1.197 (0.967; 1.427) |
| HF | wo T2DM/CVD | 0.727 (0.726; 0.728) | 0.999 (0.852; 1.147) | -0.040 (-0.151; 0.071) |
| HF | w T2DM | 0.622 (0.619; 0.625) | 0.846 (0.197; 1.495) | 0.027 (-0.294; 0.348) |
| HF | w T2DM & CVD | 0.582 (0.579; 0.584) | 1.476 (0.359; 2.593) | 0.285 (0.031; 0.538) |
| AF | wo T2DM/CVD | 0.731 (0.731; 0.732) | 1.041 (0.965; 1.116) | 0.071 (0.015; 0.128) |
| AF | w T2DM | 0.700 (0.698; 0.702) | 1.472 (0.993; 1.950) | 0.133 (-0.079; 0.345) |
| AF | w T2DM & CVD | 0.553 (0.551; 0.555) | 0.863 (-0.110; 1.837) | 0.299 (0.057; 0.541) |
| Ischaemic Stroke | wo T2DM/CVD | 0.699 (0.698; 0.700) | 0.988 (0.826; 1.150) | 0.108 (-0.001; 0.217) |
| Ischaemic Stroke | w T2DM | 0.567 (0.563; 0.570) | 0.670 (-0.521; 1.860) | 0.127 (-0.243; 0.496) |
| Ischaemic Stroke | w T2DM & CVD | 0.586 (0.582; 0.589) | 0.512 (-2.076; 3.099) | -0.053 (-0.428; 0.322) |

Individuals are stratified as followed ”wo T2DM/CVD”: participants without T2DM or CVD at baseline, “w T2DM”: participants with diabetes at baseline, “w T2DM&CVD”: participants with T2DM at baseline and a history of CVD. The analysed outcomes include cardiovascular disease including heart failure (HF) and/or atrial fibrillation (AF) (CVD+), cardiovascular disease (CVD), coronary heart disease (CHD), HF, AF, and Ischaemic Stroke. Discrimination (c-statistic) and calibration (calibration-in-the-large (CIL) and calibration slope (CS)) are based on a 20% of test set of the total data used for this study. Point estimates are presented alongside 95% CI.

**Table 16:** Discriminative performance of Age & Sex & Clinical models to predict cardiovascular disease.

| Outcome | Group | C-statistic (95%CI) | CS (95%CI) | CIL (95%CI) |
|---|---|---|---|---|
| CVD+ | wo T2DM/CVD | 0.719 (0.719; 0.720) | 1.114 (1.053; 1.175) | 0.170 (0.130; 0.211) |
| CVD+ | w T2DM | 0.681 (0.680; 0.683) | 1.553 (1.170; 1.935) | 0.359 (0.209; 0.509) |
| CVD+ | w T2DM & CVD | 0.657 (0.655; 0.659) | 4.181 (2.018; 6.343) | 1.585 (1.322; 1.848) |
| CVD | wo T2DM/CVD | 0.716 (0.716; 0.717) | 1.100 (1.027; 1.172) | 0.146 (0.097; 0.194) |
| CVD | w T2DM | 0.637 (0.635; 0.638) | 1.326 (0.863; 1.789) | 0.207 (0.037; 0.377) |
| CVD | w T2DM & CVD | 0.672 (0.670; 0.673) | 4.125 (2.281; 5.969) | 1.358 (1.114; 1.602) |
| CHD | wo T2DM/CVD | 0.724 (0.723; 0.724) | 1.100 (1.021; 1.179) | 0.128 (0.073; 0.183) |
| CHD | w T2DM | 0.623 (0.621; 0.625) | 0.999 (0.564; 1.434) | 0.159 (-0.032; 0.349) |
| CHD | w T2DM & CVD | 0.681 (0.679; 0.683) | 4.034 (2.412; 5.656) | 1.179 (0.948; 1.409) |
| HF | wo T2DM/CVD | 0.746 (0.745; 0.747) | 1.027 (0.889; 1.166) | -0.056 (-0.168; 0.055) |
| HF | w T2DM | 0.674 (0.671; 0.677) | 0.829 (0.397; 1.260) | 0.047 (-0.277; 0.371) |
| HF | w T2DM & CVD | 0.636 (0.634; 0.638) | 1.253 (0.580; 1.925) | 0.291 (0.035; 0.547) |
| AF | wo T2DM/CVD | 0.737 (0.737; 0.738) | 1.043 (0.970; 1.117) | 0.074 (0.017; 0.131) |
| AF | w T2DM | 0.727 (0.725; 0.729) | 1.455 (1.047; 1.863) | 0.140 (-0.073; 0.354) |
| AF | w T2DM & CVD | 0.550 (0.548; 0.552) | 0.650 (-0.047; 1.347) | 0.319 (0.076; 0.562) |
| Ischaemic Stroke | wo T2DM/CVD | 0.709 (0.707; 0.710) | 0.974 (0.825; 1.122) | 0.112 (0.003; 0.221) |
| Ischaemic Stroke | w T2DM | 0.617 (0.614; 0.621) | 0.616 (-0.070; 1.301) | 0.192 (-0.178; 0.563) |
| Ischaemic Stroke | w T2DM & CVD | 0.561 (0.558; 0.565) | 0.493 (-0.276; 1.262) | -0.022 (-0.401; 0.356) |

Individuals are stratified as followed ”wo T2DM/CVD”: participants without T2DM or CVD at baseline, “w T2DM”: participants with diabetes at baseline, “w T2DM&CVD”: participants with T2DM at baseline and a history of CVD. The analysed outcomes include cardiovascular disease including heart failure (HF) and/or atrial fibrillation (AF) (CVD+), cardiovascular disease (CVD), coronary heart disease (CHD), HF, AF, and Ischaemic Stroke. Discrimination (c-statistic) and calibration (calibration-in-the-large (CIL) and calibration slope (CS)) are based on a 20% of test set of the total data used for this study. Models were trained on an imputed dataset. Point estimates are presented alongside 95% CI.

**Table 17:** Net reclassification table comparing the Age & Sex & Clinical and PGS-extended models, among general population (group: "wo T2DM/CVD") with and without an CHD event during the 10 years of follow-up.

| | **PGS-extended** | | | |
|---|---|---|---|---|
| **PGS-only** | Low risk [0.0, 0.1) | Intermediate risk [0.1, 0.2) | High risk [0.2, 1.0] | **Total** |
| **In participants without CVD** | | | | |
| Low risk | 24,720 | 1,014 | 15 | 25,749 |
| Intermediate risk | 654 | 824 | 89 | 1,567 |
| High risk | 0 | 14 | 11 | 25 |
| **Total** | 25,374 | 1,852 | 115 | 27,341 |
| **In participants with CVD** | | | | |
| Low risk | 995 | 120 | 6 | 1,121 |
| Intermediate risk | 80 | 145 | 26 | 251 |
| High risk | 0 | 4 | 10 | 14 |
| **Total** | 1,075 | 269 | 42 | 1,386 |
| **NRI estimates** NRI: 0.033 (0.013; 0.058)<br>Event NRI: 0.049 (0.030, 0.075)<br>Non-event NRI: -0.016 (-0.019, -0.013)) | | Pr(Up\|Event): 0.110 (0.096; 0.133)<br>Pr(Down\|Event): 0.061 (0.051; 0.075)<br>Pr(Down\|Non-event): 0.024 (0.023; 0.026)<br>Pr(Up\|Non-event): 0.041 (0.039; 0.043) | | |

n.b. Calculations are based on the test data. Various NRI estimates are provided, including the probabilities of an increased (Up) or decreased (Down) predicted risk conditional on event status. CVD: cardiovascular disease; NRI: net reclassification index, Pr: probability.

**Table 18:** Net reclassification table comparing the Age & Sex & Clinical and PGS-extended models, among individuals with type 2 diabetes and cardiovascular disease (CVD) history (group: “w T2DM&CVD”) with and without an CHD event during the 10 years of follow-up.

| | **PGS-extended** | | | |
|---|---|---|---|---|
| **PGS-only** | Low risk [0.0, 0.1) | Intermediate risk [0.1, 0.2) | High risk [0.2, 1.0] | **Total** |
| **In participants without CVD** | | | | |
| Low risk | 0 | 0 | 0 | 0 |
| Intermediate risk | 0 | 0 | 0 | 0 |
| High risk | 1 | 7 | 97 | 105 |
| **Total** | 1 | 7 | 97 | 105 |
| **In participants with CVD** | | | | |
| Low risk | 0 | 0 | 0 | 0 |
| Intermediate risk | 0 | 0 | 0 | 0 |
| High risk | 0 | 1 | 236 | 237 |
| **Total** | 0 | 1 | 236 | 237 |
| **NRI estimates** NRI: 0.072 (0.017; 0.123)<br>Event NRI: -0.004 (-0.018; 0.000)<br>Non-event NRI: 0.076 (0.021; 0.125))) | Pr(Up\|Event): 0.000 (0.000, 0.000)<br>Pr(Down\|Event): 0.004 (0.000; 0.018)<br>Pr(Down\|Non-event): 0.076 (0.021; 0.125)<br>Pr(Up\|Non-event): 0.000 (0.000; 0.000) | | | |

n.b. Calculations are based on the test data. Various NRI estimates are provided, including the probabilities of an increased (Up) or decreased (Down) predicted risk conditional on event status. CVD: cardiovascular disease; NRI: net reclassification index, Pr: probability.

**Table 19:** The difference in c-statistics of the selected polygenic scores (PGS) versus to unweighted PGS of 144 many scores that sourced GWAS partially including UK biobank data.

| PGS name | Group | Endpoint | C-statistic | | | Difference in C-statistic | P-value | R-square | MAF |
|---|---|---|---|---|---|---|---|---|---|
| | | | Selected PGS | Selected PGS Unweighted | Unweighted PGS | | | | |
| AF | wo T2DM/CVD | CVD+ | 0.58 | no | 0.58 | 0.00 | $5 \times 10^{-4}$ | 0.2 | 0.01 |
| AF | w T2DM | CVD+ | 0.58 | yes | 0.58 | 0.00 | $5 \times 10^{-4}$ | 0.2 | 0.01 |
| AF | w T2DM&CVD | CVD+ | 0.53 | yes | 0.53 | 0.00 | $5 \times 10^{-5}$ | 0.2 | 0.01 |
| AF | wo T2DM/CVD | CVD | 0.53 | no | 0.53 | 0.00 | $5 \times 10^{-4}$ | 0.01 | 0.01 |
| AF | w T2DM | CVD | 0.54 | yes | 0.54 | 0.00 | $5 \times 10^{-4}$ | 0.4 | 0.01 |
| AF | w T2DM&CVD | CVD | 0.53 | yes | 0.53 | 0.00 | $5 \times 10^{-5}$ | 0.2 | 0.01 |
| AF | wo T2DM/CVD | CHD | 0.53 | no | 0.53 | 0.00 | $5 \times 10^{-4}$ | 0.01 | 0.01 |
| AF | w T2DM | CHD | 0.54 | yes | 0.54 | 0.00 | $5 \times 10^{-4}$ | 0.4 | 0.01 |
| AF | w T2DM&CVD | CHD | 0.52 | yes | 0.52 | 0.00 | $5 \times 10^{-5}$ | 0.2 | 0.01 |
| AF | wo T2DM/CVD | HF | 0.58 | yes | 0.58 | 0.00 | $5 \times 10^{-4}$ | 0.01 | 0.01 |
| AF | w T2DM | HF | 0.57 | yes | 0.57 | 0.00 | $5 \times 10^{-4}$ | 0.01 | 0.01 |
| AF | w T2DM&CVD | HF | 0.56 | no | 0.55 | 0.01 | $5 \times 10^{-4}$ | 0.4 | 0.01 |
| AF | wo T2DM/CVD | AF | 0.66 | no | 0.66 | 0.00 | $5 \times 10^{-4}$ | 0.2 | 0.01 |
| AF | w T2DM | AF | 0.68 | yes | 0.68 | 0.00 | $5 \times 10^{-4}$ | 0.2 | 0.01 |
| AF | w T2DM&CVD | AF | 0.65 | yes | 0.65 | 0.00 | $5 \times 10^{-4}$ | 0.01 | 0.01 |
| AF | wo T2DM/CVD | Is. Stroke | 0.54 | no | 0.54 | 0.01 | $5 \times 10^{-4}$ | 0.01 | 0.01 |
| AF | w T2DM | Is. Stroke | 0.56 | yes | 0.56 | 0.00 | $5 \times 10^{-5}$ | 0.6 | 0.01 |
| AF | w T2DM&CVD | Is. Stroke | 0.58 | yes | 0.58 | 0.00 | $5 \times 10^{-4}$ | 0.01 | 0.01 |
| BMI | wo T2DM/CVD | CVD+ | 0.52 | yes | 0.52 | 0.00 | $5 \times 10^{-5}$ | 0.01 | 0.01 |
| BMI | w T2DM | CVD+ | 0.55 | yes | 0.55 | 0.00 | $5 \times 10^{-4}$ | 0.01 | 0.01 |
| BMI | w T2DM&CVD | CVD+ | 0.53 | yes | 0.53 | 0.00 | $5 \times 10^{-4}$ | 0.01 | 0.01 |
| BMI | wo T2DM/CVD | CVD | 0.52 | yes | 0.52 | 0.00 | $5 \times 10^{-4}$ | 0.01 | 0.01 |
| BMI | w T2DM | CVD | 0.55 | no | 0.55 | 0.00 | $5 \times 10^{-7}$ | 0.01 | 0.01 |
| BMI | w T2DM&CVD | CVD | 0.52 | no | 0.52 | 0.00 | $5 \times 10^{-4}$ | 0.01 | 0.01 |
| BMI | wo T2DM/CVD | CHD | 0.53 | yes | 0.53 | 0.00 | $5 \times 10^{-4}$ | 0.01 | 0.01 |
| BMI | w T2DM | CHD | 0.55 | no | 0.55 | 0.00 | $5 \times 10^{-7}$ | 0.01 | 0.01 |
| BMI | w T2DM&CVD | CHD | 0.53 | yes | 0.53 | 0.00 | $5 \times 10^{-6}$ | 0.01 | 0.01 |
| BMI | wo T2DM/CVD | HF | 0.56 | no | 0.56 | 0.00 | $5 \times 10^{-4}$ | 0.2 | 0.01 |
| BMI | w T2DM | HF | 0.58 | yes | 0.58 | 0.00 | $5 \times 10^{-8}$ | 0.2 | 0.01 |
| BMI | w T2DM&CVD | HF | 0.56 | yes | 0.56 | 0.00 | $5 \times 10^{-5}$ | 0.01 | 0.01 |
| BMI | wo T2DM/CVD | AF | 0.52 | yes | 0.52 | 0.00 | $5 \times 10^{-5}$ | 0.01 | 0.01 |
| BMI | w T2DM | AF | 0.55 | yes | 0.55 | 0.00 | $5 \times 10^{-8}$ | 0.01 | 0.01 |
| BMI | w T2DM&CVD | AF | 0.52 | yes | 0.52 | 0.00 | $5 \times 10^{-8}$ | 0.01 | 0.01 |
| BMI | wo T2DM/CVD | Is. Stroke | 0.51 | yes | 0.51 | 0.00 | $5 \times 10^{-8}$ | 0.01 | 0.01 |
| BMI | w T2DM | Is. Stroke | 0.54 | yes | 0.54 | 0.00 | $5 \times 10^{-4}$ | 0.01 | 0.01 |
| BMI | w T2DM&CVD | Is. Stroke | 0.55 | no | 0.55 | 0.01 | $5 \times 10^{-7}$ | 0.6 | 0.01 |
| CAD in diabetes | wo T2DM/CVD | CVD+ | 0.51 | no | 0.51 | 0.00 | $5 \times 10^{-7}$ | 0.8 | 0.01 |
| CAD in diabetes | w T2DM | CVD+ | 0.63 | no | 0.62 | 0.01 | $5 \times 10^{-4}$ | 0.6 | 0.01 |
| CAD in diabetes | w T2DM&CVD | CVD+ | 0.63 | no | 0.61 | 0.01 | $5 \times 10^{-4}$ | 0.01 | 0.01 |
| CAD in diabetes | wo T2DM/CVD | CVD | 0.52 | no | 0.52 | 0.00 | $5 \times 10^{-7}$ | 0.8 | 0.01 |
| CAD in diabetes | w T2DM | CVD | 0.67 | no | 0.66 | 0.01 | $5 \times 10^{-4}$ | 0.2 | 0.01 |
| CAD in diabetes | w T2DM&CVD | CVD | 0.64 | no | 0.63 | 0.02 | $5 \times 10^{-4}$ | 0.01 | 0.01 |
| CAD in diabetes | wo T2DM/CVD | CHD | 0.52 | no | 0.52 | 0.00 | $5 \times 10^{-8}$ | 0.01 | 0.01 |
| CAD in diabetes | w T2DM | CHD | 0.71 | no | 0.69 | 0.02 | $5 \times 10^{-4}$ | 0.01 | 0.01 |
| CAD in diabetes | w T2DM&CVD | CHD | 0.67 | no | 0.66 | 0.01 | $5 \times 10^{-4}$ | 0.01 | 0.01 |
| CAD in diabetes | wo T2DM/CVD | HF | 0.51 | yes | 0.51 | 0.00 | $5 \times 10^{-5}$ | 0.8 | 0.01 |
| CAD in diabetes | w T2DM | HF | 0.62 | no | 0.60 | 0.01 | $5 \times 10^{-4}$ | 0.01 | 0.01 |
| CAD in diabetes | w T2DM&CVD | HF | 0.56 | yes | 0.56 | 0.00 | $5 \times 10^{-4}$ | 0.01 | 0.01 |
| CAD in diabetes | wo T2DM/CVD | AF | 0.51 | no | 0.51 | 0.00 | $5 \times 10^{-7}$ | 0.8 | 0.01 |
| CAD in diabetes | w T2DM | AF | 0.58 | no | 0.58 | 0.01 | $5 \times 10^{-4}$ | 0.4 | 0.01 |
| CAD in diabetes | w T2DM&CVD | AF | 0.52 | no | 0.50 | 0.03 | $5 \times 10^{-7}$ | 0.8 | 0.01 |
| CAD in diabetes | wo T2DM/CVD | Is. Stroke | 0.51 | yes | 0.51 | 0.00 | $5 \times 10^{-7}$ | 0.8 | 0.01 |
| CAD in diabetes | w T2DM | Is. Stroke | 0.56 | no | 0.55 | 0.00 | $5 \times 10^{-6}$ | 0.6 | 0.01 |
| CAD in diabetes | w T2DM&CVD | Is. Stroke | 0.57 | yes | 0.57 | 0.00 | $5 \times 10^{-7}$ | 0.8 | 0.01 |

Continued on next page

**Table 19 – continued from previous page**

| PGS name | group | Endpoint | C-statistic | | | Difference in C-statistic | P-value | R-square | MAF |
|---|---|---|---|---|---|---|---|---|---|
| | | | Selected PGS | Selected PGS Unweighted | Unweighted PGS | | | | |
| CKD | wo T2DM/CVD | CVD+ | 0.51 | no | 0.51 | 0.00 | $5\times10^{-5}$ | 0.2 | 0.01 |
| CKD | w T2DM | CVD+ | 0.52 | yes | 0.52 | 0.00 | $5\times10^{-4}$ | 0.2 | 0.01 |
| CKD | w T2DM&CVD | CVD+ | 0.54 | no | 0.52 | 0.02 | $5\times10^{-4}$ | 0.4 | 0.01 |
| CKD | wo T2DM/CVD | CVD | 0.51 | no | 0.51 | 0.00 | $5\times10^{-5}$ | 0.2 | 0.01 |
| CKD | w T2DM | CVD | 0.52 | yes | 0.52 | 0.00 | $5\times10^{-7}$ | 0.2 | 0.01 |
| CKD | w T2DM&CVD | CVD | 0.54 | no | 0.52 | 0.02 | $5\times10^{-4}$ | 0.4 | 0.01 |
| CKD | wo T2DM/CVD | CHD | 0.51 | no | 0.51 | 0.00 | $5\times10^{-5}$ | 0.2 | 0.01 |
| CKD | w T2DM | CHD | 0.52 | no | 0.52 | 0.00 | $5\times10^{-4}$ | 0.8 | 0.01 |
| CKD | w T2DM&CVD | CHD | 0.54 | yes | 0.54 | 0.00 | $5\times10^{-7}$ | 0.4 | 0.01 |
| CKD | wo T2DM/CVD | HF | 0.51 | yes | 0.51 | 0.00 | $5\times10^{-4}$ | 0.8 | 0.01 |
| CKD | w T2DM | HF | 0.54 | no | 0.52 | 0.02 | $5\times10^{-6}$ | 0.01 | 0.01 |
| CKD | w T2DM&CVD | HF | 0.54 | yes | 0.54 | 0.00 | $5\times10^{-6}$ | 0.01 | 0.01 |
| CKD | wo T2DM/CVD | AF | 0.51 | no | 0.50 | 0.00 | $5\times10^{-8}$ | 0.01 | 0.01 |
| CKD | w T2DM | AF | 0.52 | yes | 0.52 | 0.00 | $5\times10^{-4}$ | 0.2 | 0.01 |
| CKD | w T2DM&CVD | AF | 0.53 | yes | 0.53 | 0.00 | $5\times10^{-6}$ | 0.01 | 0.01 |
| CKD | wo T2DM/CVD | Is. Stroke | 0.51 | yes | 0.51 | 0.00 | $5\times10^{-4}$ | 0.2 | 0.01 |
| CKD | w T2DM | Is. Stroke | 0.56 | yes | 0.56 | 0.00 | $5\times10^{-4}$ | 0.2 | 0.01 |
| CKD | w T2DM&CVD | Is. Stroke | 0.54 | yes | 0.54 | 0.00 | $5\times10^{-6}$ | 0.01 | 0.01 |
| HF | wo T2DM/CVD | CVD+ | 0.53 | yes | 0.53 | 0.00 | $5\times10^{-4}$ | 0.6 | 0.01 |
| HF | w T2DM | CVD+ | 0.54 | yes | 0.54 | 0.00 | $5\times10^{-4}$ | 0.6 | 0.01 |
| HF | w T2DM&CVD | CVD+ | 0.55 | no | 0.55 | 0.00 | $5\times10^{-7}$ | 0.01 | 0.01 |
| HF | wo T2DM/CVD | CVD | 0.53 | no | 0.52 | 0.00 | $5\times10^{-7}$ | 0.8 | 0.01 |
| HF | w T2DM | CVD | 0.54 | yes | 0.54 | 0.00 | $5\times10^{-7}$ | 0.01 | 0.01 |
| HF | w T2DM&CVD | CVD | 0.55 | no | 0.55 | 0.00 | $5\times10^{-7}$ | 0.01 | 0.01 |
| HF | wo T2DM/CVD | CHD | 0.53 | no | 0.53 | 0.00 | $5\times10^{-7}$ | 0.6 | 0.01 |
| HF | w T2DM | CHD | 0.53 | yes | 0.53 | 0.00 | $5\times10^{-6}$ | 0.01 | 0.01 |
| HF | w T2DM&CVD | CHD | 0.56 | no | 0.55 | 0.00 | $5\times10^{-8}$ | 0.2 | 0.01 |
| HF | wo T2DM/CVD | HF | 0.60 | no | 0.60 | 0.00 | $5\times10^{-4}$ | 0.6 | 0.01 |
| HF | w T2DM | HF | 0.63 | yes | 0.63 | 0.00 | $5\times10^{-4}$ | 0.2 | 0.01 |
| HF | w T2DM&CVD | HF | 0.60 | no | 0.59 | 0.01 | $5\times10^{-4}$ | 0.2 | 0.01 |
| HF | wo T2DM/CVD | AF | 0.55 | no | 0.55 | 0.01 | $5\times10^{-8}$ | 0.6 | 0.01 |
| HF | w T2DM | AF | 0.53 | no | 0.53 | 0.00 | $5\times10^{-4}$ | 0.6 | 0.01 |
| HF | w T2DM&CVD | AF | 0.59 | no | 0.58 | 0.01 | $5\times10^{-4}$ | 0.2 | 0.01 |
| HF | wo T2DM/CVD | Is. Stroke | 0.52 | no | 0.52 | 0.00 | $5\times10^{-8}$ | 0.8 | 0.01 |
| HF | w T2DM | Is. Stroke | 0.59 | yes | 0.59 | 0.00 | $5\times10^{-6}$ | 0.8 | 0.01 |
| HF | w T2DM&CVD | Is. Stroke | 0.57 | yes | 0.57 | 0.00 | $5\times10^{-6}$ | 0.8 | 0.01 |
| SBP | wo T2DM/CVD | CVD+ | 0.53 | no | 0.52 | 0.00 | $5\times10^{-4}$ | 0.01 | 0.01 |
| SBP | w T2DM | CVD+ | 0.54 | no | 0.53 | 0.01 | $5\times10^{-4}$ | 0.2 | 0.01 |
| SBP | w T2DM&CVD | CVD+ | 0.52 | no | 0.52 | 0.01 | $5\times10^{-7}$ | 0.01 | 0.01 |
| SBP | wo T2DM/CVD | CVD | 0.53 | no | 0.53 | 0.00 | $5\times10^{-6}$ | 0.01 | 0.01 |
| SBP | w T2DM | CVD | 0.54 | no | 0.53 | 0.00 | $5\times10^{-4}$ | 0.2 | 0.01 |
| SBP | w T2DM&CVD | CVD | 0.52 | no | 0.52 | 0.01 | $5\times10^{-5}$ | 0.2 | 0.01 |
| SBP | wo T2DM/CVD | CHD | 0.54 | no | 0.53 | 0.00 | $5\times10^{-6}$ | 0.01 | 0.01 |
| SBP | w T2DM | CHD | 0.53 | no | 0.53 | 0.01 | $5\times10^{-4}$ | 0.2 | 0.01 |
| SBP | w T2DM&CVD | CHD | 0.53 | no | 0.52 | 0.00 | $5\times10^{-6}$ | 0.2 | 0.01 |
| SBP | wo T2DM/CVD | HF | 0.53 | yes | 0.53 | 0.00 | $5\times10^{-4}$ | 0.01 | 0.01 |
| SBP | w T2DM | HF | 0.55 | yes | 0.55 | 0.00 | $5\times10^{-4}$ | 0.8 | 0.01 |
| SBP | w T2DM&CVD | HF | 0.53 | no | 0.53 | 0.00 | $5\times10^{-4}$ | 0.4 | 0.01 |
| SBP | wo T2DM/CVD | AF | 0.52 | no | 0.52 | 0.00 | $5\times10^{-7}$ | 0.01 | 0.01 |
| SBP | w T2DM | AF | 0.54 | yes | 0.54 | 0.00 | $5\times10^{-6}$ | 0.01 | 0.01 |
| SBP | w T2DM&CVD | AF | 0.55 | yes | 0.55 | 0.00 | $5\times10^{-4}$ | 0.6 | 0.01 |
| SBP | wo T2DM/CVD | Is. Stroke | 0.53 | no | 0.53 | 0.00 | $5\times10^{-6}$ | 0.01 | 0.01 |
| SBP | w T2DM | Is. Stroke | 0.57 | yes | 0.57 | 0.00 | $5\times10^{-4}$ | 0.01 | 0.01 |
| SBP | w T2DM&CVD | Is. Stroke | 0.58 | no | 0.58 | 0.00 | $5\times10^{-8}$ | 0.2 | 0.01 |
| T2DM | wo T2DM/CVD | CVD+ | 0.51 | yes | 0.51 | 0.00 | $5\times10^{-4}$ | 0.2 | 0.01 |
| T2DM | w T2DM | CVD+ | 0.52 | no | 0.52 | 0.00 | $5\times10^{-8}$ | 0.2 | 0.01 |

Continued on next page

**Table 19 – continued from previous page**

| PGS name | group | Endpoint | C-statistic | | | Difference in C-statistic | P-value | R-square | MAF |
|---|---|---|---|---|---|---|---|---|---|
| | | | Selected PGS | Selected PGS Unweighted | Unweighted PGS | | | | |
| T2DM | w T2DM&CVD | CVD+ | 0.54 | yes | 0.54 | 0.00 | $5 \times 10^{-7}$ | 0.6 | 0.01 |
| T2DM | wo T2DM/CVD | CVD | 0.51 | yes | 0.51 | 0.00 | $5 \times 10^{-4}$ | 0.2 | 0.01 |
| T2DM | w T2DM | CVD | 0.52 | yes | 0.52 | 0.00 | $5 \times 10^{-6}$ | 0.01 | 0.01 |
| T2DM | w T2DM&CVD | CVD | 0.53 | yes | 0.53 | 0.00 | $5 \times 10^{-7}$ | 0.4 | 0.01 |
| T2DM | wo T2DM/CVD | CHD | 0.52 | yes | 0.52 | 0.00 | $5 \times 10^{-4}$ | 0.2 | 0.01 |
| T2DM | w T2DM | CHD | 0.53 | yes | 0.53 | 0.00 | $5 \times 10^{-6}$ | 0.01 | 0.01 |
| T2DM | w T2DM&CVD | CHD | 0.52 | yes | 0.52 | 0.00 | $5 \times 10^{-7}$ | 0.4 | 0.01 |
| T2DM | wo T2DM/CVD | HF | 0.52 | yes | 0.52 | 0.00 | $5 \times 10^{-4}$ | 0.2 | 0.01 |
| T2DM | w T2DM | HF | 0.54 | yes | 0.54 | 0.00 | $5 \times 10^{-4}$ | 0.01 | 0.01 |
| T2DM | w T2DM&CVD | HF | 0.56 | yes | 0.56 | 0.00 | $5 \times 10^{-6}$ | 0.2 | 0.01 |
| T2DM | wo T2DM/CVD | AF | 0.51 | yes | 0.51 | 0.00 | $5 \times 10^{-7}$ | 0.2 | 0.01 |
| T2DM | w T2DM | AF | 0.52 | no | 0.52 | 0.00 | $5 \times 10^{-4}$ | 0.01 | 0.01 |
| T2DM | w T2DM&CVD | AF | 0.53 | no | 0.53 | 0.00 | $5 \times 10^{-6}$ | 0.4 | 0.01 |
| T2DM | wo T2DM/CVD | Is. Stroke | 0.51 | no | 0.51 | 0.00 | $5 \times 10^{-8}$ | 0.6 | 0.01 |
| T2DM | w T2DM | Is. Stroke | 0.53 | no | 0.53 | 0.01 | $5 \times 10^{-6}$ | 0.01 | 0.01 |
| T2DM | w T2DM&CVD | Is. Stroke | 0.54 | yes | 0.54 | 0.00 | $5 \times 10^{-4}$ | 0.2 | 0.01 |
| eGFR | wo T2DM/CVD | CVD+ | 0.51 | yes | 0.51 | 0.00 | $5 \times 10^{-8}$ | 0.2 | 0.01 |
| eGFR | w T2DM | CVD+ | 0.52 | yes | 0.52 | 0.00 | $5 \times 10^{-4}$ | 0.2 | 0.01 |
| eGFR | w T2DM&CVD | CVD+ | 0.57 | yes | 0.57 | 0.00 | $5 \times 10^{-4}$ | 0.01 | 0.01 |
| eGFR | wo T2DM/CVD | CVD | 0.51 | yes | 0.51 | 0.00 | $5 \times 10^{-7}$ | 0.2 | 0.01 |
| eGFR | w T2DM | CVD | 0.54 | no | 0.54 | 0.00 | $5 \times 10^{-4}$ | 0.8 | 0.01 |
| eGFR | w T2DM&CVD | CVD | 0.56 | yes | 0.56 | 0.00 | $5 \times 10^{-4}$ | 0.01 | 0.01 |
| eGFR | wo T2DM/CVD | CHD | 0.51 | yes | 0.51 | 0.00 | $5 \times 10^{-7}$ | 0.2 | 0.01 |
| eGFR | w T2DM | CHD | 0.53 | yes | 0.53 | 0.00 | $5 \times 10^{-7}$ | 0.8 | 0.01 |
| eGFR | w T2DM&CVD | CHD | 0.54 | yes | 0.54 | 0.00 | $5 \times 10^{-5}$ | 0.01 | 0.01 |
| eGFR | wo T2DM/CVD | HF | 0.52 | no | 0.52 | 0.00 | $5 \times 10^{-4}$ | 0.8 | 0.01 |
| eGFR | w T2DM | HF | 0.56 | yes | 0.56 | 0.00 | $5 \times 10^{-5}$ | 0.2 | 0.01 |
| eGFR | w T2DM&CVD | HF | 0.53 | yes | 0.53 | 0.00 | $5 \times 10^{-8}$ | 0.6 | 0.01 |
| eGFR | wo T2DM/CVD | AF | 0.51 | no | 0.50 | 0.00 | $5 \times 10^{-4}$ | 0.01 | 0.01 |
| eGFR | w T2DM | AF | 0.52 | yes | 0.52 | 0.00 | $5 \times 10^{-5}$ | 0.01 | 0.01 |
| eGFR | w T2DM&CVD | AF | 0.54 | yes | 0.54 | 0.00 | $5 \times 10^{-7}$ | 0.2 | 0.01 |
| eGFR | wo T2DM/CVD | Is. Stroke | 0.51 | yes | 0.51 | 0.00 | $5 \times 10^{-7}$ | 0.2 | 0.01 |
| eGFR | w T2DM | Is. Stroke | 0.56 | no | 0.55 | 0.01 | $5 \times 10^{-8}$ | 0.8 | 0.01 |
| eGFR | w T2DM&CVD | Is. Stroke | 0.57 | no | 0.57 | 0.00 | $5 \times 10^{-6}$ | 0.01 | 0.01 |

Individuals are stratified as followed ”wo T2DM/CVD”: participants without T2DM or CVD at baseline, “w T2DM”: participants with diabetes at baseline, “w T2DM&CVD”: participants with T2DM at baseline and a history of CVD. The PGS parameters include: association p-value theshold, R-square threshold for pairwise linkage disequilibrium (LD) and the minor allele frequency (MAF) threshold.

**Table 20:** Performance of Age & Sex & Clinical models trained on a complete case dataset to predict cardiovascular disease.

| Outcome | Group | C-statistic (95%CI) | CS (95%CI) | CIL (95%CI) |
|---|---|---|---|---|
| CVD+ | wo T2DM/CVD | 0.718 (0.718; 0.719) | 1.107 (1.038; 1.175) | 0.173 (0.128; 0.219) |
| CVD+ | w T2DM | 0.678 (0.676; 0.679) | 1.491 (1.084; 1.898) | 0.330 (0.167; 0.492) |
| CVD+ | w T2DM & CVD | 0.665 (0.663; 0.667) | 7.019 (3.663; 10.376) | 1.521 (1.241; 1.802) |
| CVD | wo T2DM/CVD | 0.713 (0.712; 0.713) | 1.067 (0.988; 1.147) | 0.171 (0.117; 0.225) |
| CVD | w T2DM | 0.639 (0.637; 0.641) | 1.305 (0.811; 1.800) | 0.206 (0.023; 0.389) |
| CVD | w T2DM & CVD | 0.683 (0.681; 0.686) | 5.843 (3.269; 8.417) | 1.312 (1.050; 1.574) |
| CHD | wo T2DM/CVD | 0.719 (0.718; 0.719) | 1.077 (0.990; 1.165) | 0.156 (0.095; 0.216) |
| CHD | w T2DM | 0.615 (0.614; 0.617) | 0.935 (0.467; 1.403) | 0.162 (-0.042; 0.366) |
| CHD | w T2DM & CVD | 0.681 (0.679; 0.683) | 5.580 (3.292; 7.868) | 1.173 (0.923; 1.423) |
| HF | wo T2DM/CVD | 0.752 (0.751; 0.754) | 1.105 (0.945; 1.266) | -0.057 (-0.181; 0.066) |
| HF | w T2DM | 0.679 (0.675; 0.682) | 0.780 (0.339; 1.220) | 0.047 (-0.300; 0.394) |
| HF | w T2DM & CVD | 0.635 (0.632; 0.637) | 1.303 (0.579; 2.027) | 0.275 (-0.005; 0.555) |
| AF | wo T2DM/CVD | 0.738 (0.737; 0.738) | 1.045 (0.962; 1.128) | 0.046 (-0.018; 0.110) |
| AF | w T2DM | 0.699 (0.697; 0.701) | 1.193 (0.786; 1.600) | 0.055 (-0.181; 0.291) |
| AF | w T2DM & CVD | 0.517 (0.514; 0.519) | 0.304 (-0.445; 1.054) | 0.365 (0.105; 0.625) |
| Ischaemic Stroke | wo T2DM/CVD | 0.708 (0.707; 0.709) | 0.970 (0.805; 1.136) | 0.119 (-0.003; 0.242) |
| Ischaemic Stroke | w T2DM | 0.593 (0.589; 0.597) | 0.638 (-0.261; 1.537) | 0.055 (-0.360; 0.470) |
| Ischaemic Stroke | w T2DM & CVD | 0.540 (0.536; 0.544) | 0.588 (-0.559; 1.735) | -0.186 (-0.623; 0.251) |

Individuals are stratified as followed ”wo T2DM/CVD”: participants without T2DM or CVD at baseline, “w T2DM”: participants with diabetes at baseline, “w T2DM&CVD”: participants with T2DM at baseline and a history of CVD. Discrimination (c-statistic) and calibration (calibration-in-the-large (CIL) and calibration slope (CS)) are based on a 20% of test set of the total data used for this study. Point estimates are presented alongside 95% CI.

**Table 21:** Performance of PGS-extended models to predict cardiovascular disease stratified by prevalent and incident T2DM.

| Outcome | Group | Prevalent/Incident T2DM | C-statistic (95%CI) |
|---|---|---|---|
| CVD+ | w T2DM | Prevalent T2DM | 0.713 (0.712; 0.715) |
| CVD+ | w T2DM | Incident T2DM | 0.747 (0.742; 0.751) |
| CVD+ | w T2DM & CVD | Prevalent T2DM | 0.624 (0.622; 0.627) |
| CVD+ | w T2DM & CVD | Incident T2DM | 0.618 (0.611; 0.625) |
| CVD | w T2DM | Prevalent T2DM | 0.711 (0.710; 0.713) |
| CVD | w T2DM | Incident T2DM | 0.738 (0.733; 0.743) |
| CVD | w T2DM & CVD | Prevalent T2DM | 0.648 (0.646; 0.651) |
| CVD | w T2DM & CVD | Incident T2DM | 0.761 (0.756; 0.767) |
| CHD | w T2DM | Prevalent T2DM | 0.739 (0.737; 0.741) |
| CHD | w T2DM | Incident T2DM | 0.832 (0.827; 0.837) |
| CHD | w T2DM & CVD | Prevalent T2DM | 0.718 (0.716; 0.720) |
| CHD | w T2DM & CVD | Incident T2DM | 0.753 (0.747; 0.759) |
| HF | w T2DM | Prevalent T2DM | 0.701 (0.698; 0.704) |
| HF | w T2DM | Incident T2DM | 0.835 (0.819; 0.850) |
| HF | w T2DM & CVD | Prevalent T2DM | 0.653 (0.651; 0.656) |
| HF | w T2DM & CVD | Incident T2DM | 0.493 (0.485; 0.501) |
| AF | w T2DM | Prevalent T2DM | 0.763 (0.761; 0.764) |
| AF | w T2DM | Incident T2DM | 0.758 (0.752; 0.764) |
| AF | w T2DM & CVD | Prevalent T2DM | 0.598 (0.596; 0.600) |
| AF | w T2DM & CVD | Incident T2DM | 0.671 (0.664; 0.679) |
| Ischaemic Stroke | w T2DM | Prevalent T2DM | 0.632 (0.628; 0.636) |
| Ischaemic Stroke | w T2DM | Incident T2DM | 0.641 (0.630; 0.652) |
| Ischaemic Stroke | w T2DM & CVD | Prevalent T2DM | 0.458 (0.454; 0.461) |
| Ischaemic Stroke | w T2DM & CVD | Incident T2DM | 0.540 (0.526; 0.554) |

Individuals are stratified as followed ”wo T2DM/CVD”: participants without T2DM or CVD at baseline, “w T2DM”: participants with diabetes at baseline, “w T2DM&CVD”: participants with T2DM at baseline and a history of CVD. Discrimination (c-statistic) are based on a 20% of test set of the total data used for this study. Models were trained on an imputed dataset. Point estimates are presented alongside 95% CI.

**Table 22:** Performance of clinical only based models (Age&Sex&Clinical) to predict cardiovascular disease stratified by prevalent and incident T2DM.

| Outcome | Group | Prevalent/Incident T2DM | C-statistic (95%CI) |
|---|---|---|---|
| CVD+ | w T2DM | Prevalent T2DM | 0.671 (0.670; 0.672) |
| CVD+ | w T2DM | Incident T2DM | 0.741 (0.737; 0.745) |
| CVD+ | w T2DM & CVD | Prevalent T2DM | 0.656 (0.653; 0.658) |
| CVD+ | w T2DM & CVD | Incident T2DM | 0.694 (0.688; 0.701) |
| CVD | w T2DM | Prevalent T2DM | 0.628 (0.627; 0.630) |
| CVD | w T2DM | Incident T2DM | 0.691 (0.686; 0.696) |
| CVD | w T2DM & CVD | Prevalent T2DM | 0.667 (0.665; 0.669) |
| CVD | w T2DM & CVD | Incident T2DM | 0.727 (0.721; 0.733) |
| CHD | w T2DM | Prevalent T2DM | 0.616 (0.615; 0.618) |
| CHD | w T2DM | Incident T2DM | 0.684 (0.678; 0.690) |
| CHD | w T2DM & CVD | Prevalent T2DM | 0.679 (0.677; 0.681) |
| CHD | w T2DM & CVD | Incident T2DM | 0.725 (0.719; 0.731) |
| HF | w T2DM | Prevalent T2DM | 0.666 (0.663; 0.669) |
| HF | w T2DM | Incident T2DM | 0.603 (0.584; 0.622) |
| HF | w T2DM & CVD | Prevalent T2DM | 0.630 (0.627; 0.632) |
| HF | w T2DM & CVD | Incident T2DM | 0.693 (0.685; 0.701) |
| AF | w T2DM | Prevalent T2DM | 0.733 (0.731; 0.735) |
| AF | w T2DM | Incident T2DM | 0.692 (0.686; 0.699) |
| AF | w T2DM & CVD | Prevalent T2DM | 0.552 (0.549; 0.554) |
| AF | w T2DM & CVD | Incident T2DM | 0.553 (0.545; 0.560) |
| Ischaemic Stroke | w T2DM | Prevalent T2DM | 0.605 (0.601; 0.608) |
| Ischaemic Stroke | w T2DM | Incident T2DM | 0.734 (0.723; 0.744) |
| Ischaemic Stroke | w T2DM & CVD | Prevalent T2DM | 0.561 (0.558; 0.565) |
| Ischaemic Stroke | w T2DM & CVD | Incident T2DM | 0.580 (0.566; 0.594) |

Individuals are stratified as followed ”wo T2DM/CVD”: participants without T2DM or CVD at baseline, “w T2DM”: participants with diabetes at baseline, “w T2DM&CVD”: participants with T2DM at baseline and a history of CVD. Discrimination (c-statistic) are based on a 20% of test set of the total data used for this study. Models were trained on an imputed dataset. Point estimates are presented alongside 95% CI.

**Table 23:** Number of CVD events during a 10-year follow-up period stratified by training and testing samples.

| Outcomes | wo T2DM/CVD | | | w T2DM | | | w T2DM&CVD | | |
|---|---|---|---|---|---|---|---|---|---|
| | Train | Test | Overall (%) | Train | Test | Overall (%) | Train | Test | Overall |
| | 114,732 | 28,727 | 143,459 | 4,181 | 1,048 | 5,229 | 1,279 | 342 | 1,621 |
| CVD+AF+HF | 10,456 (9.1) | 2,677 (9.3) | 13,133 (9.2) | 833 (19.9) | 224 (21.4) | 1,057 (20.2) | 1,023 (80.0) | 272 (79.5) | 1,295 (79.9) |
| CVD | 6,814 (5.9) | 1,796 (6.3) | 8,610 (6.0) | 628 (15.0) | 160 (15.3) | 788 (15.1) | 969 (75.8) | 255 (74.6) | 1,224 (75.5) |
| CHD | 5,255 (4.6) | 1,386 (4.8) | 6,641 (4.6) | 486 (11.6) | 122 (11.6) | 608 (11.6) | 895 (70.0) | 237 (69.3) | 1,132 (69.8) |
| Isch. Stroke | 1,192 (1.0) | 328 (1.1) | 1,520 (1.1) | 103 (2.5) | 29 (2.8) | 132 (2.5) | 127 (9.9) | 30 (8.8) | 157 (9.7) |
| HF | 1,336 (1.2) | 316 (1.1) | 1,652 (1.2) | 157 (3.8) | 39 (3.7) | 196 (3.7) | 287 (22.4) | 78 (22.8) | 365 (22.5) |
| AF | 5,110 (4.5) | 1,278 (4.4) | 6,388 (4.5) | 377 (9.0) | 96 (9.2) | 473 (9.0) | 340 (26.6) | 90 (26.3) | 430 (26.5) |

n.b.events were stratified by training (80%) and testing samples (20%) using the model derivation and evaluation. The UK biobank participants are stratified into groups: participants without a history of CVD and T2DM (wo T2DM/CVD), participants with type 2 diabetes (w T2DM), participants with a history of CVD prior to a T2DM diagnosis (w T2DM&CVD).

**Table 24:** Net reclassification comparing the predicted CHD risk distributions of Age & Sex & Clinical and PGS-extended, among individuals with type 2 diabetes (group: “w T2DM”) with and without an CHD event during the 10 years of follow-up.

| **Age & Sex & Clinical** | **PGS-extended** Low risk [0.0, 0.1) | Intermediate risk [0.1, 0.2) | High risk [0.2, 1.0] | **Total** |
|---|---|---|---|---|
| **In participants without CVD** | | | | |
| Low risk | 384 | 99 | 30 | 513 |
| Intermediate risk | 256 | 94 | 50 | 400 |
| High risk | 3 | 4 | 6 | 13 |
| **Total** | 643 | 197 | 86 | 926 |
| **In participants with CVD** | | | | |
| Low risk | 17 | 17 | 15 | 49 |
| Intermediate risk | 25 | 16 | 29 | 70 |
| High risk | 0 | 1 | 2 | 3 |
| **Total** | 42 | 34 | 46 | 122 |
| **NRI estimates** NRI: 0.378 (0.210; 0.511)<br>Event NRI: 0.287 (0.118, 0.423)<br>Non-event NRI: 0.091 (0.047, 0.146)) | Pr(Up\|Event): 0.500 (0.411; 0.589)<br>Pr(Down\|Event): 0.213 (0.149; 0.308)<br>Pr(Down\|Non-event): 0.284 (0.261; 0.311)<br>Pr(Up\|Non-event): 0.193 (0.170; 0.220) | | | |

n.b. Calculations are based on the test data. Various NRI estimates are provided, including the probabilities of an increased (Up) or decreased (Down) predicted risk conditional on event status. CVD: cardiovascular disease; NRI: net reclassification index, Pr: probability.

### Figures

**Figure 1:** Workflow of the quality control (QC) steps for the UK Biobank genotype data.

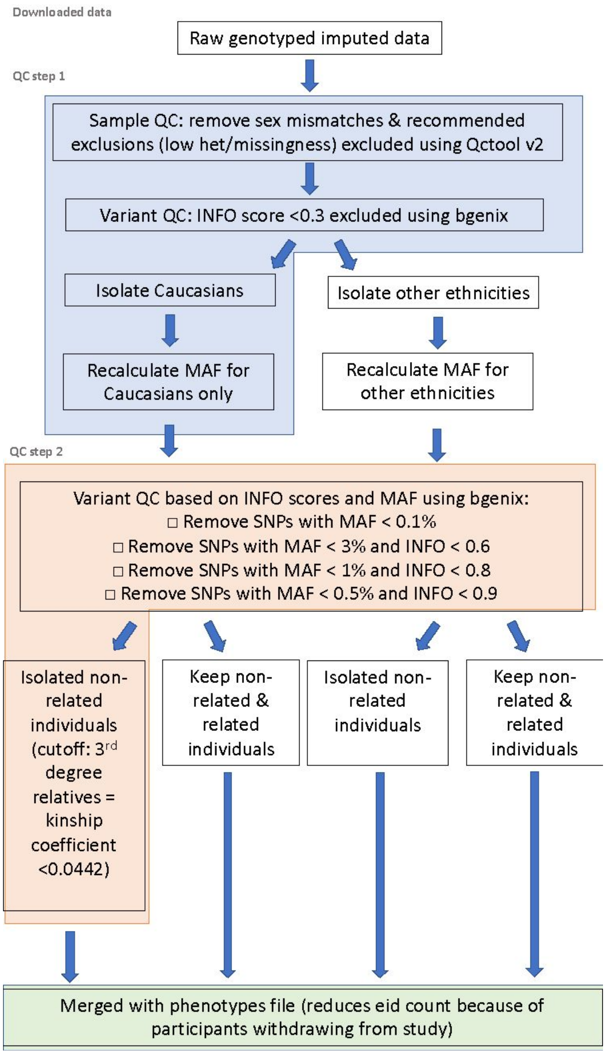

n.b. QC step 1 is highlighted in purple and QC step 2 in orange. The highlighted section in green represents the final merge between the genetic data and the sample list. The sample list is continuously updated with participant withdrawals. The non-highlighted QC steps have not been completed. Caucasians refer to individuals of European ancestry.

**Figure 2:** Workflow of data stratification into study samples.

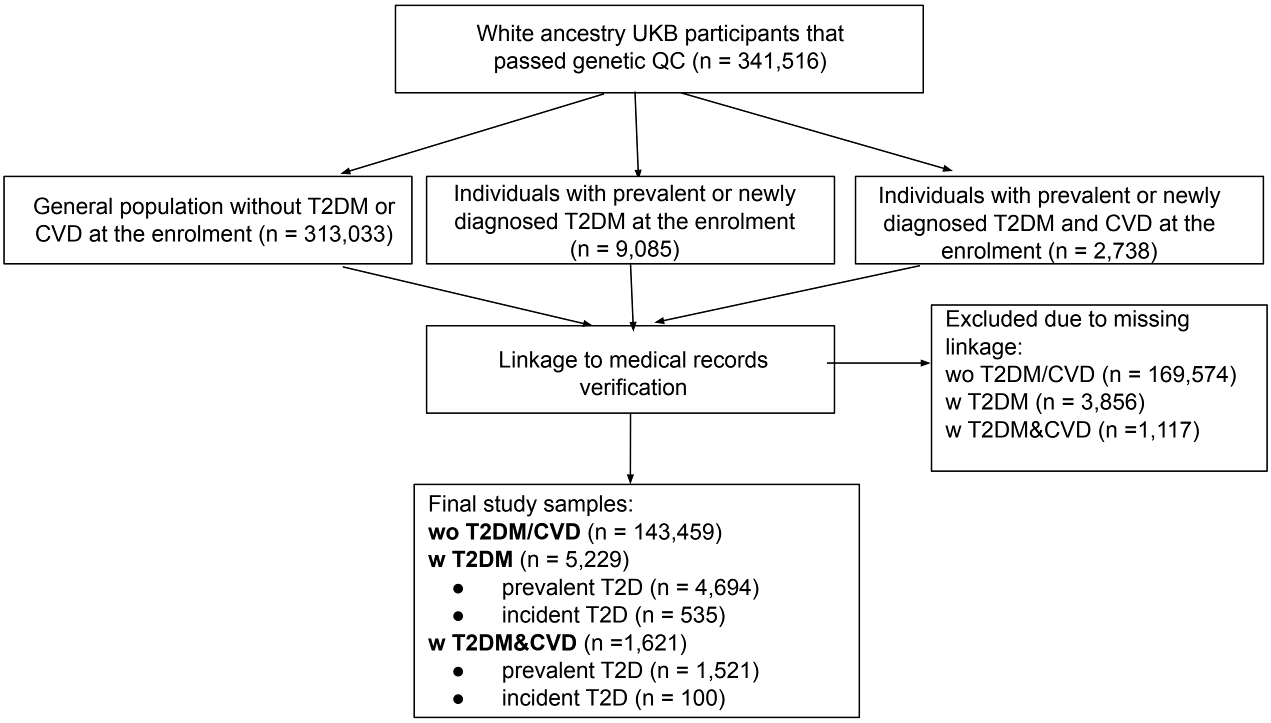

n.b. The ”wo T2DM/CVD” group indicates a general population without a history of CVD or T2DM. The ”w T2DM” group includes individuals with T2DM but not CVD history. The ”w T2DM&CVD” group includes individuals with T2DM and a history of CVD.

**Figure 3:** Flow diagram for the antihypertensive and lipid-lowering prescription extraction from the UK Biobank dataset.

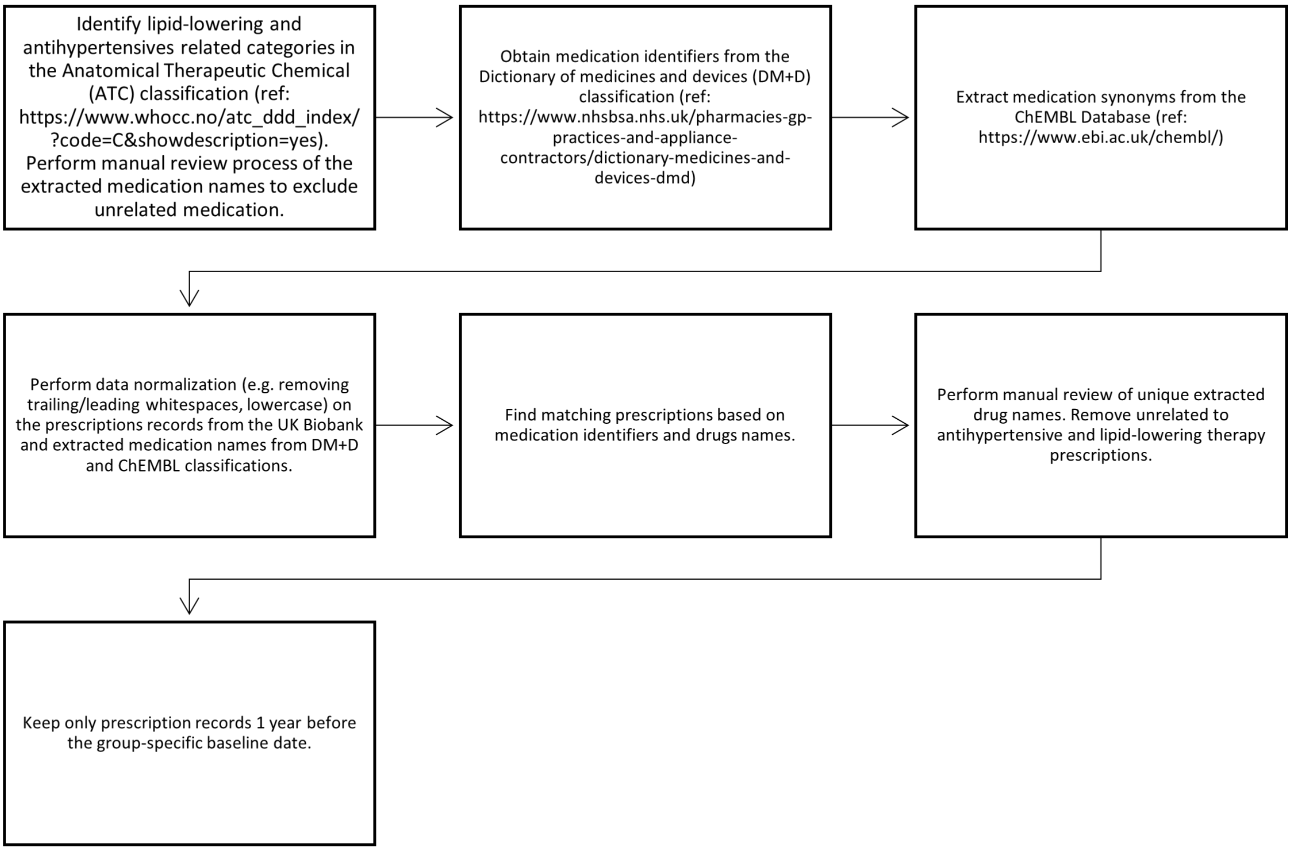

**Figure 4:** Flow diagram for type 2 diabetes identification.

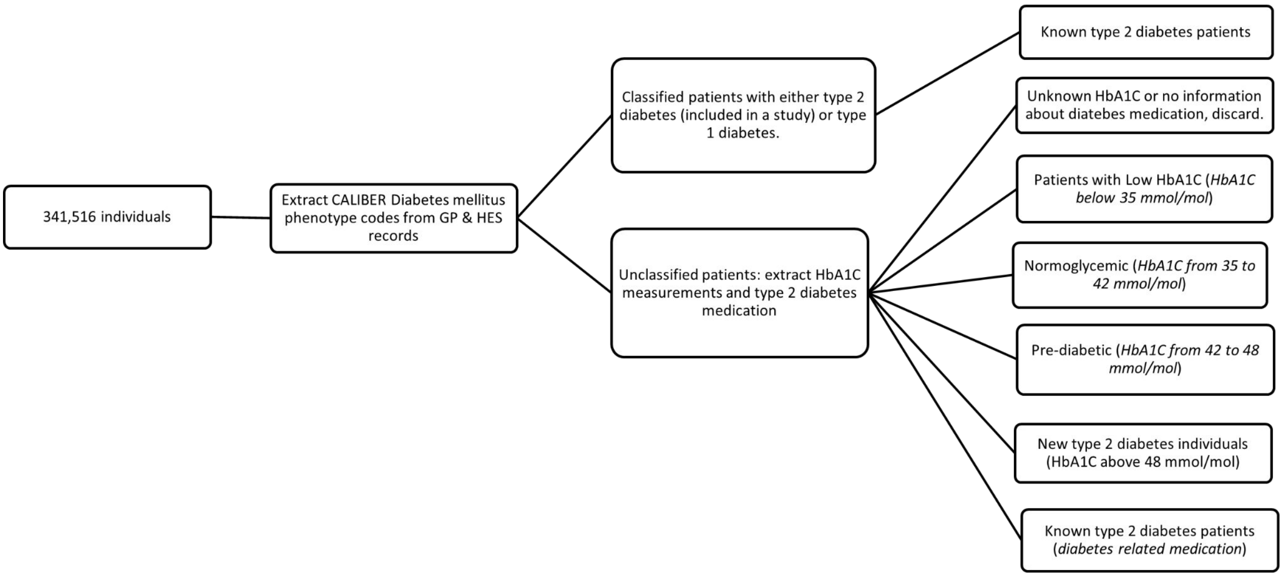

n.b. Phenotype codes were extracted from `https://www.caliberresearch.org/portal/phenotypes/diabetes`, type 2 diabetes medication is presented in Appendix Table 4. Phenotype codes are extracted from General practice (GP) and Hospital Episode Statistics (HES).

**Figure 5:** Discrimination of the best performing PGS scores.

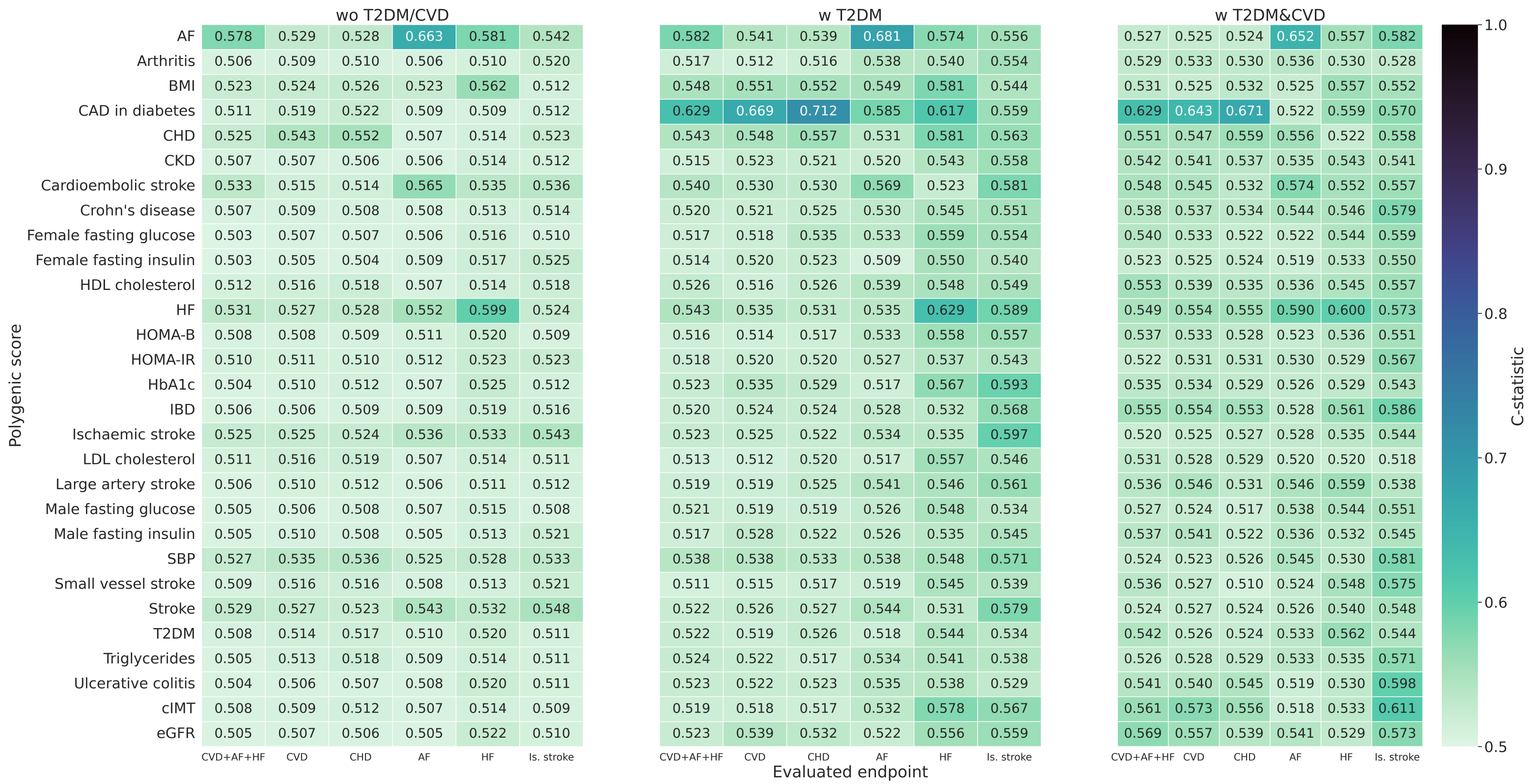

Individuals are stratified as followed ”wo T2DM/CVD”: participants without T2DM or CVD at baseline, “w T2DM”: participants with diabetes at baseline, “w T2DM&CVD”: participants with T2DM at baseline and a history of CVD.

**Figure 6:** The number of unique genetic variants used in the polygenic score trained to predict the x-axis listed cardiovascular disease.

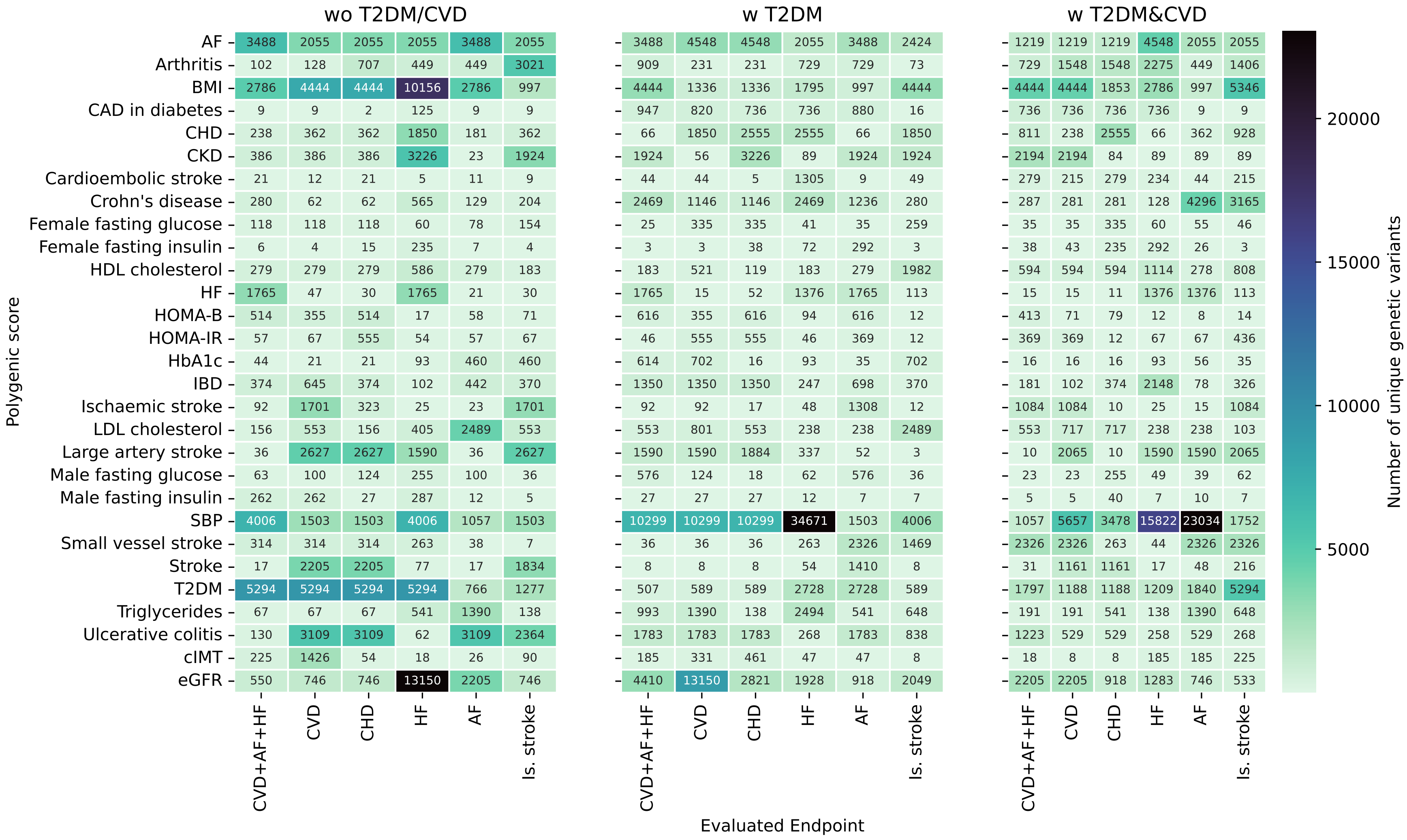

Individuals are stratified as followed ”wo T2DM/CVD”: participants without T2DM or CVD at baseline, “w T2DM”: participants with diabetes at baseline, “w T2DM&CVD”: participants with T2DM at baseline and a history of CVD.

**Figure 7:** Spearman's rank correlation for the 29 optimized univariable GS for the "wo T2DM/CVD" group for CVD endpoint.

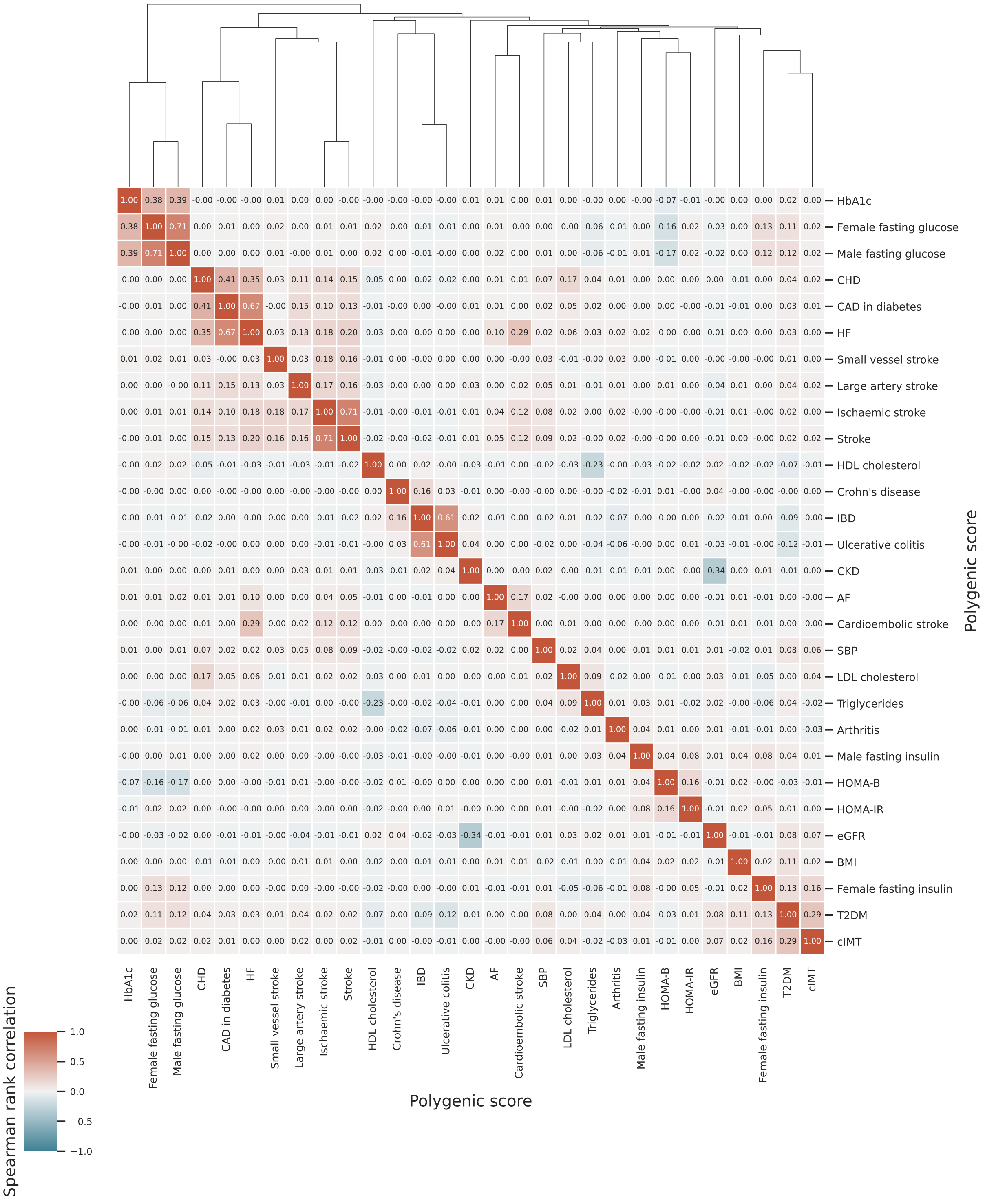

n.b. Spearman's rank correlation is a measure of linear correlation between two sets of data (here between two different genetic scores). A value of +1 indicates a perfect positive linear relationship between two GS, -1 indicates a perfect negative linear relationship, while a value of 0 indicates that the variables are linearly unrelated. The "wo T2DM/CVD" group includes participants without T2DM or CVD at baseline.

**Figure 8:** Spearman's rank correlation for the 29 optimized univariable GS for the "w T2DM" group for CVD endpoint.

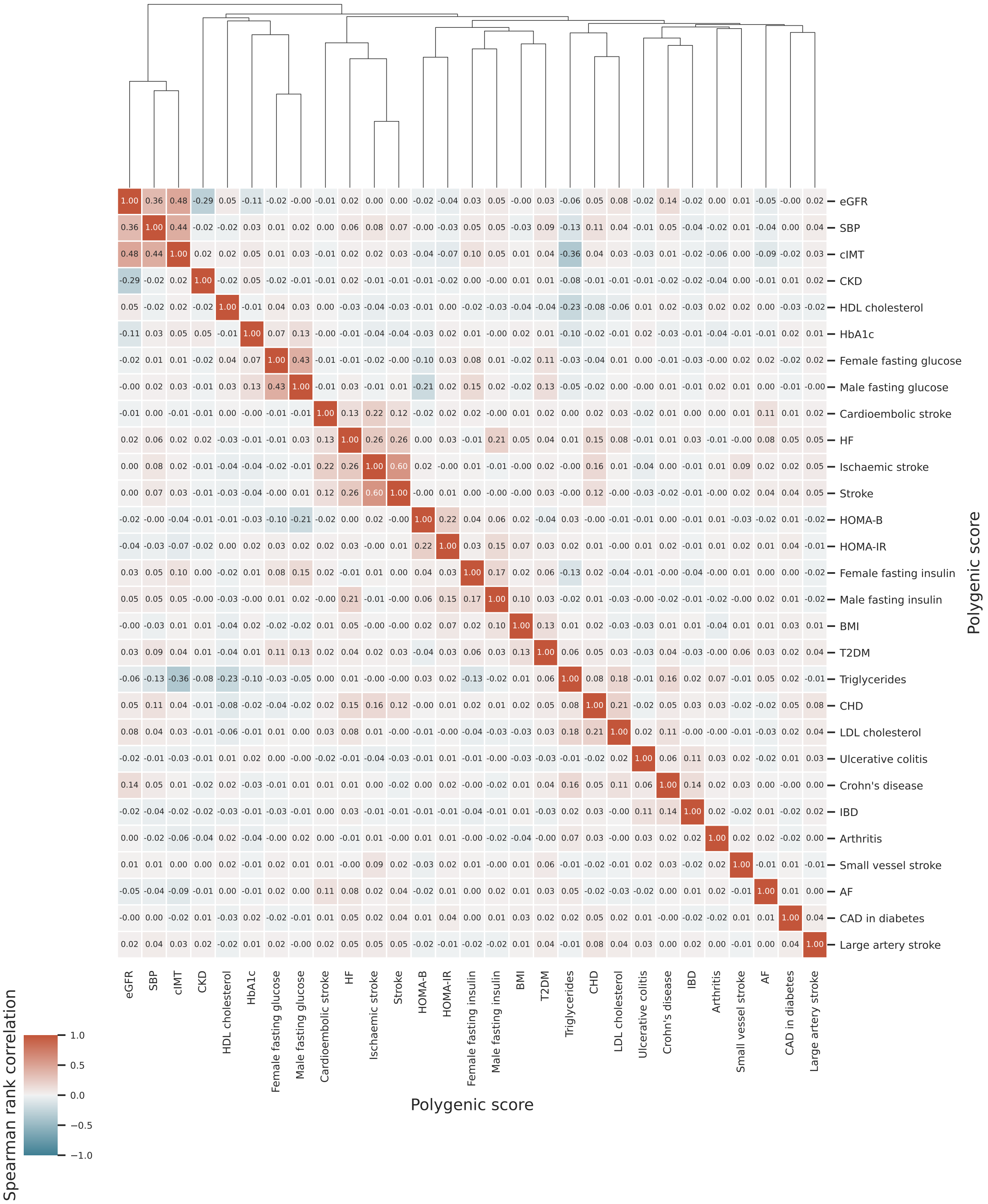

n.b. Spearman's rank correlation is a measure of linear correlation between two sets of data (here between two different genetic scores). A value of +1 indicates a perfect positive linear relationship between two GS, -1 indicates a perfect negative linear relationship, while a value of 0 indicates that the variables are linearly unrelated. The "w T2DM" includes participants with diabetes at baseline group.

**Figure 9:** Spearman's rank correlation for the 29 optimized univariable GS for the "w T2DM&CVD" group for CVD endpoint.

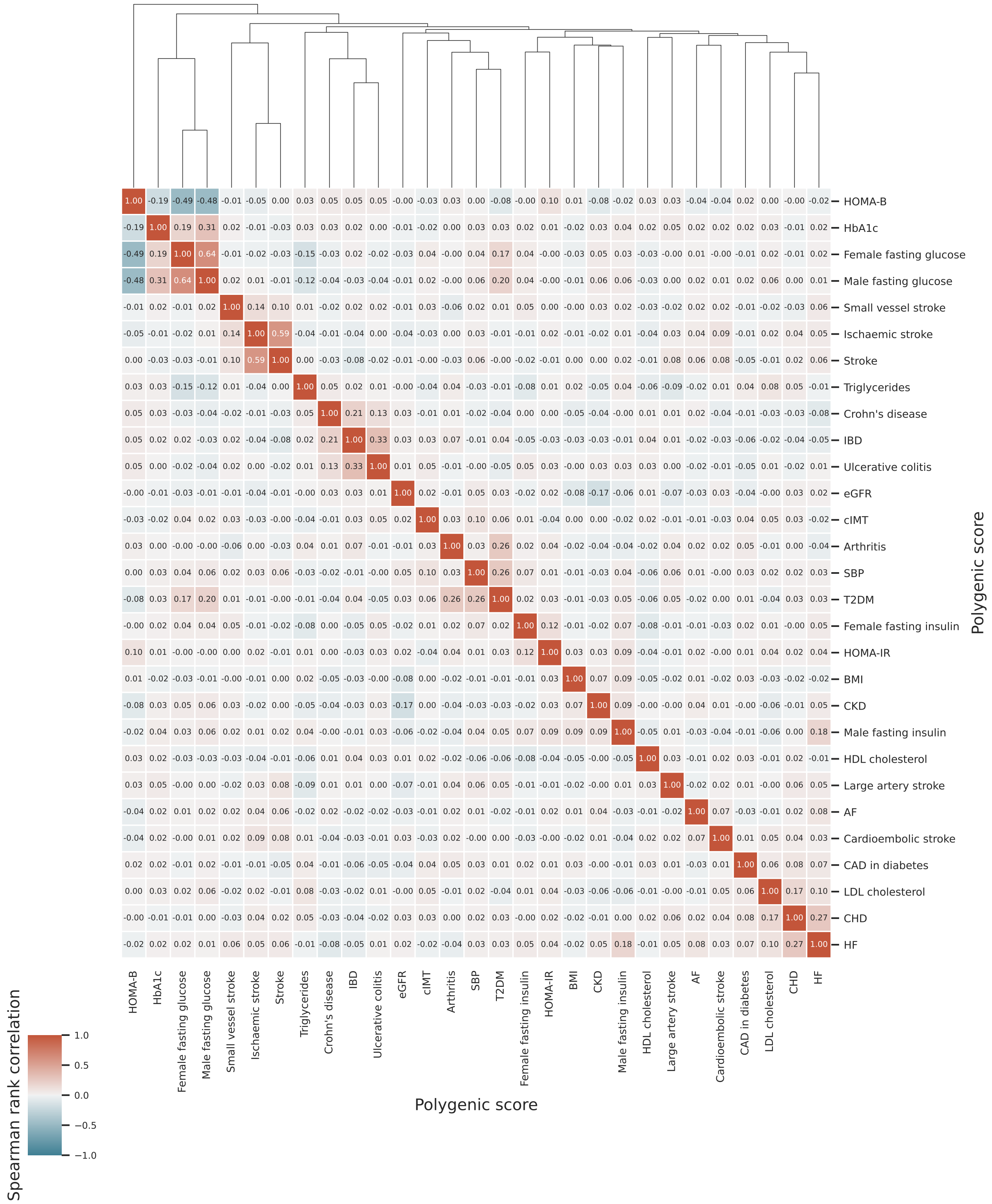

n.b. Spearman's rank correlation is a measure of linear correlation between two sets of data (here between two different genetic scores). A value of +1 indicates a perfect positive linear relationship between two GS, -1 indicates a perfect negative linear relationship, while a value of 0 indicates that the variables are linearly unrelated. The "w T2DM&CVD" group includes participants with T2DM at baseline and a history of CVD.

**Figure 10:** Training discriminative performance of four distinct machine learners stacking the univariable polygenic scores.

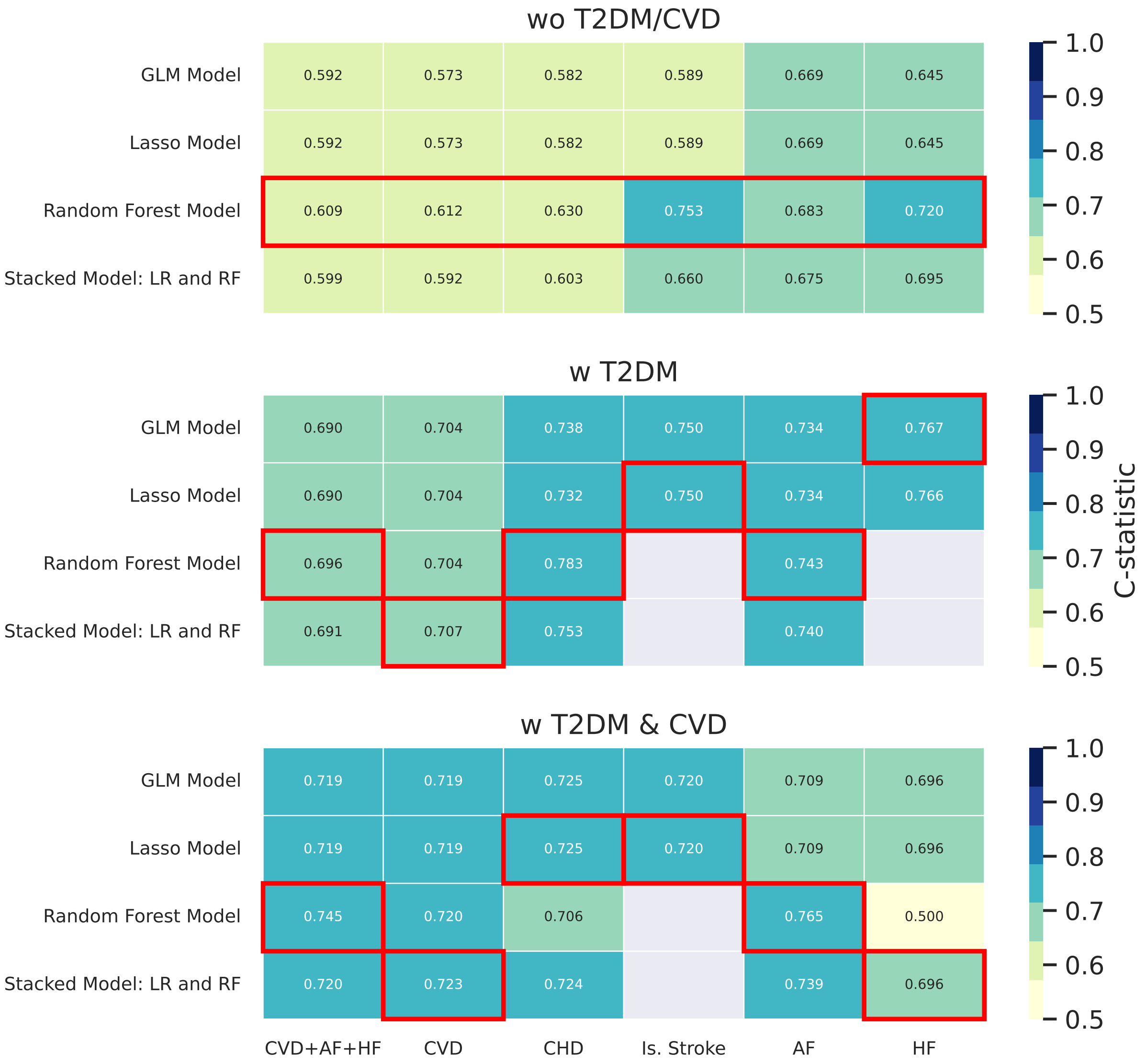

n.b. Individuals are stratified as followed ”wo T2DM/CVD”: participants without T2DM or CVD at baseline, “w T2DM”: participants with diabetes at baseline, “w T2DM&CVD”: participants with T2DM at baseline and a history of CVD. The discrimination measurement (c-statistic) was calculated using the 10-fold cross-validation of the training dataset (20% of the input data).The cross-validation is a resampling method that uses different proportions of the data to test and train a model on different iterations. GLM means Generalized linear model, RF means Random Forest, and LR indicates Logistic Regression.

**Figure 11:** An exemplar classification tree from the 200 many trees that were used by the ”PGS-only” for 10 years CHD risk in participants without T2DM or CVD at baseline.

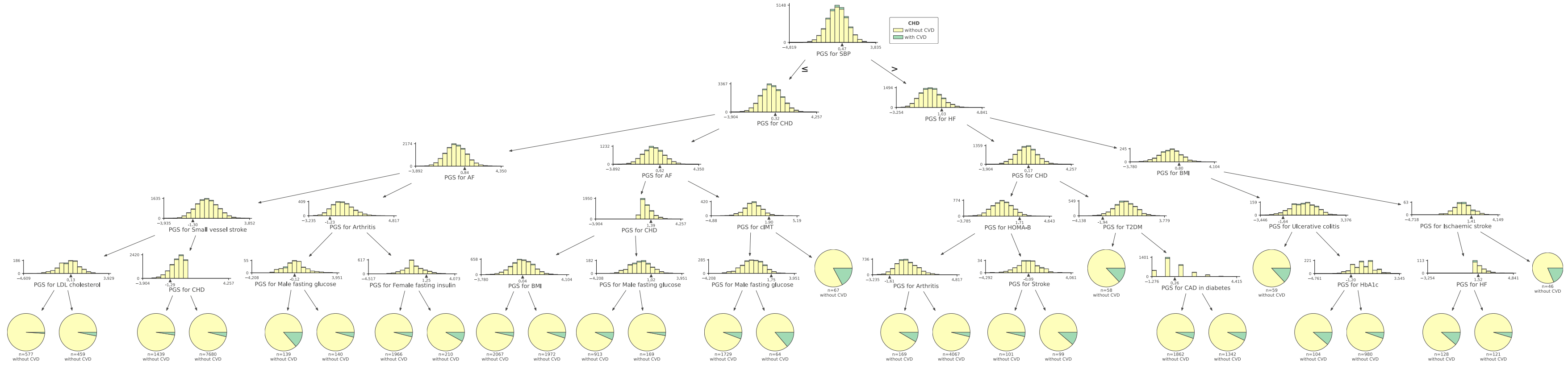

The ”PGS-only” refers to a single stacked polygenic score (PGS) prediction model.

**Figure 12:** An exemplar classification tree from the 150 many trees that were used by the ”PGS-only” model for 10 years CHD risk in participants with T2DM without history of CVD at baseline.

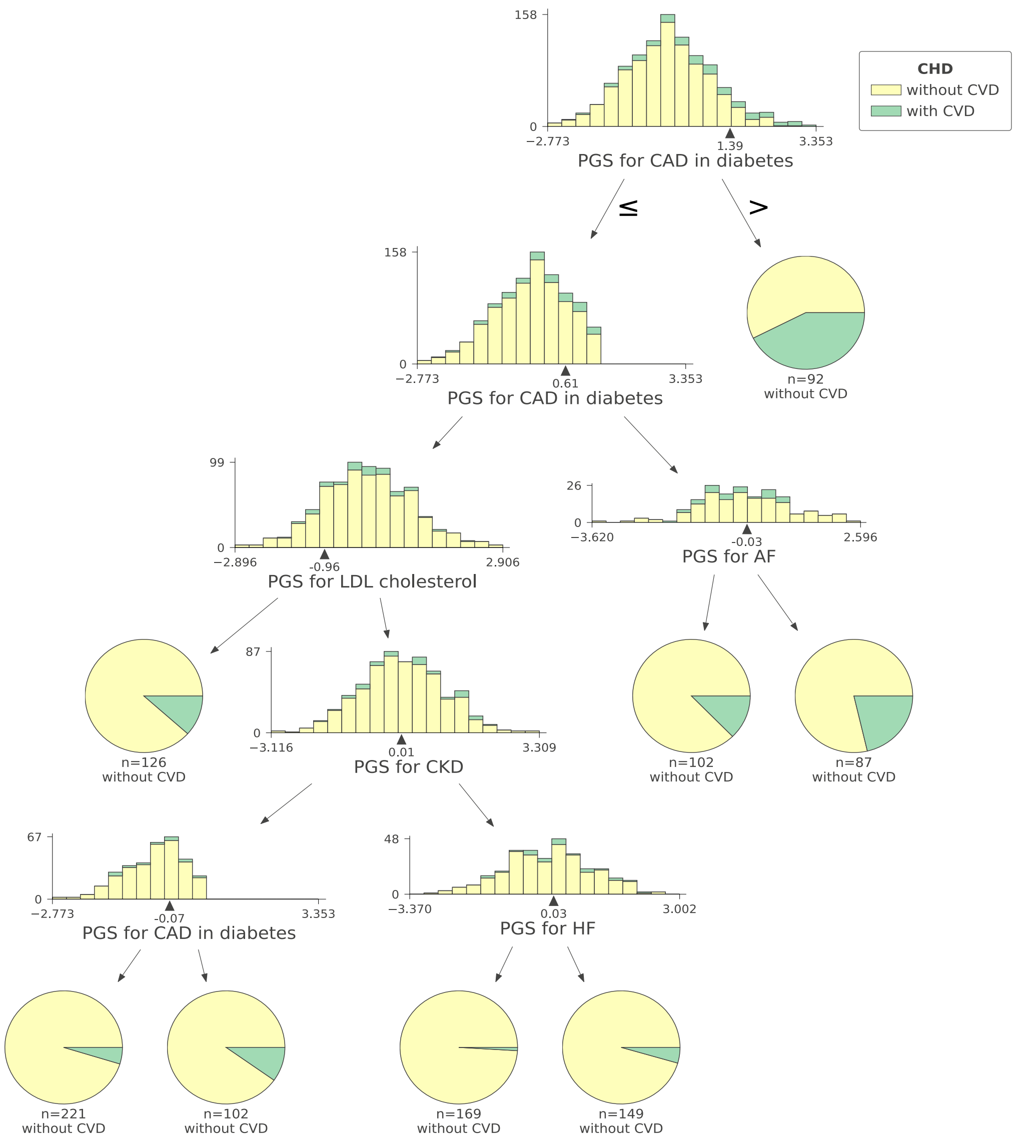

The ”PGS-only” refers to a single stacked polygenic score (PGS) prediction model.

**Figure 13:** An exemplar classification tree from the 100 many trees that were used by the ”PGS-only” model for 10 years CHD risk in participants with T2DM and a history of CVD.

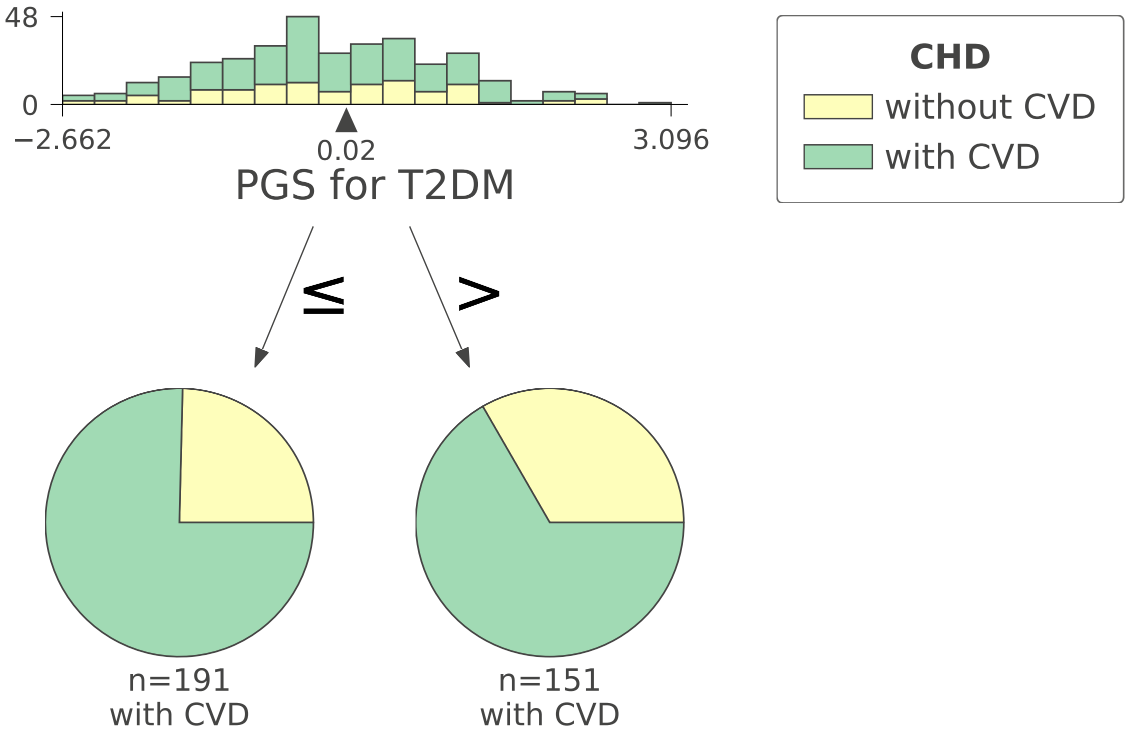

The ”PGS-only” refers to a single stacked polygenic score (PGS) prediction model.

**Figure 14:** The contribution of each of the 29 considered univariable polygenic scores to an overall stacked polygenic score (PGS-only).

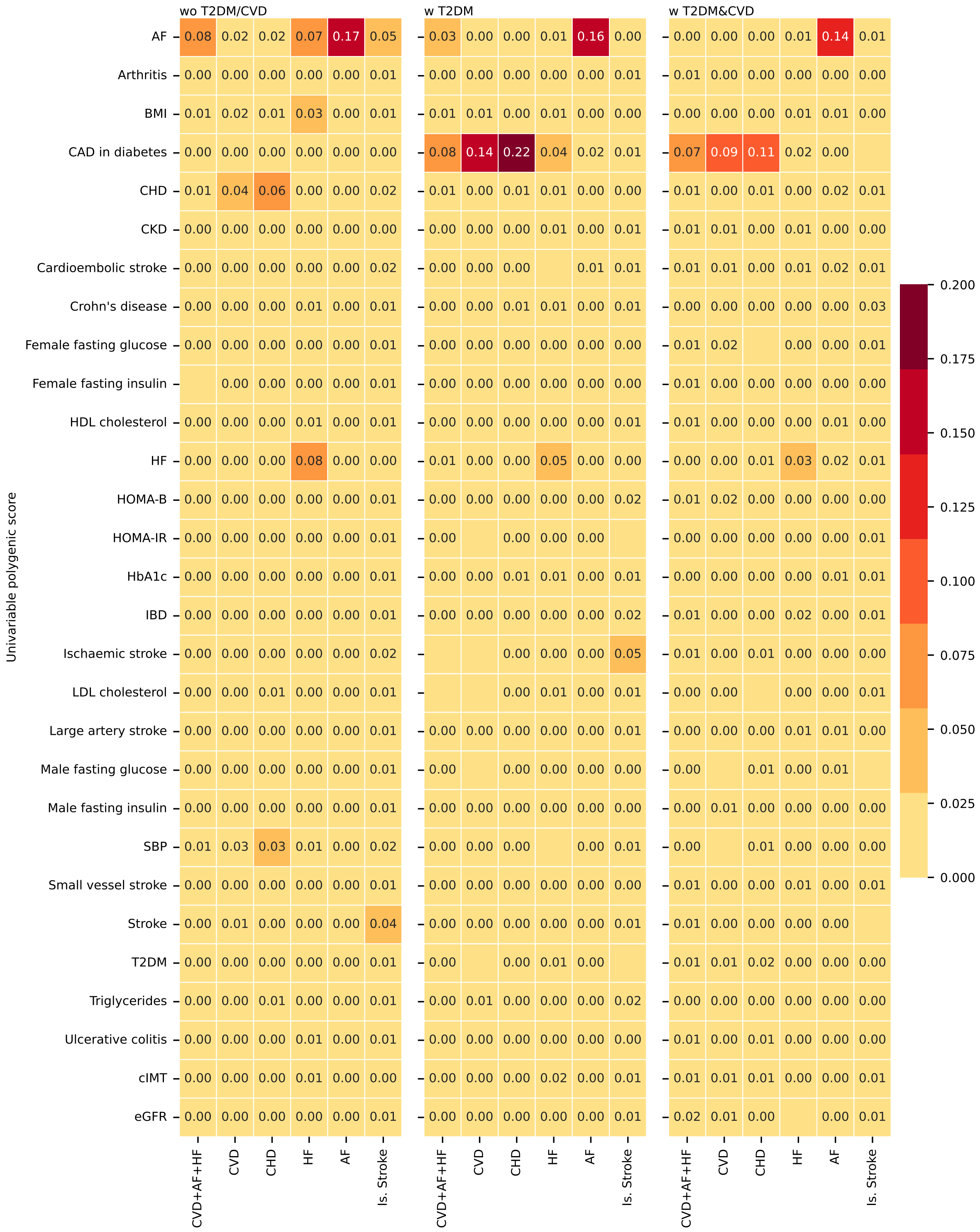

n.b. The permutation feature importance assesses the c-statistic change in the test data; iteratively the values of each GS were randomly assigned to an individual after which the c-statistic was re-estimated with these permuted data and the difference in performance used as an estimate of GS contribution to the model's predictive potential. Empty cells indicate variables with low, almost irrelevant feature importance for a specific model (feature importance is less than 0.0001). Individuals are stratified as followed "wo T2DM/CVD: participants without T2DM or CVD at baseline, "w T2DM": participants with type 2 diabetes at baseline, "w T2DM&CVD": participants with type 2 diabetes at baseline and a history of CVD.

**Figure 15:** Calibration plots for the PGS-only models stratified by outcome and participant group.

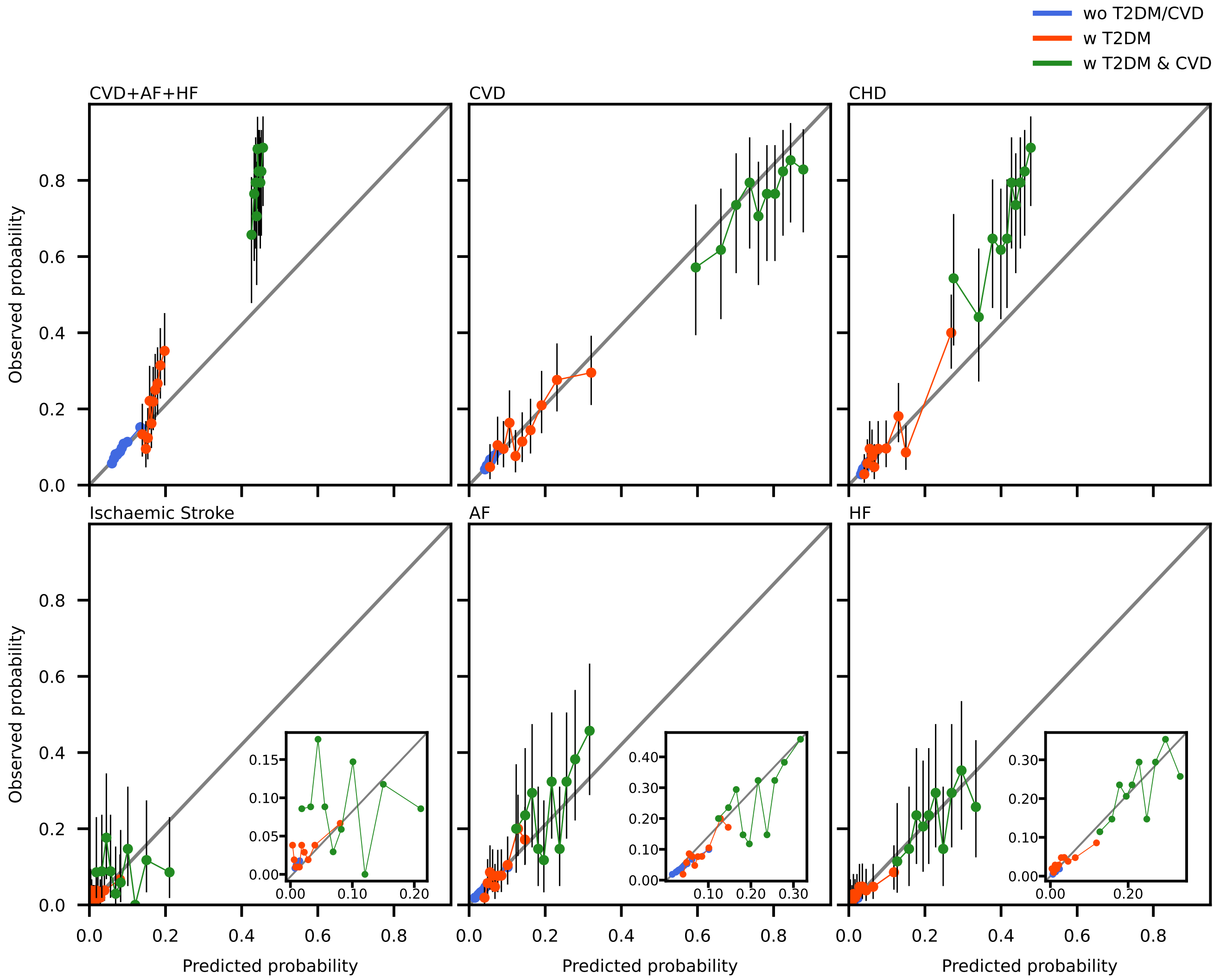

n.b The PGS-only models considered information from 29 univariable polygenic scores. Individuals are stratified as followed "wo T2DM/CVD: participants without T2DM or CVD at baseline, "w T2DM": participants with type 2 diabetes at baseline, "w T2DM&CVD": participants with type 2 diabetes at baseline and a history of CVD. The vertical line segments represent 95% confidence intervals under the assumption of a beta-distribution.

**Figure 16:** Probability distribution plots for the PGS-only models stratified by outcome, participant group, and case/control group.

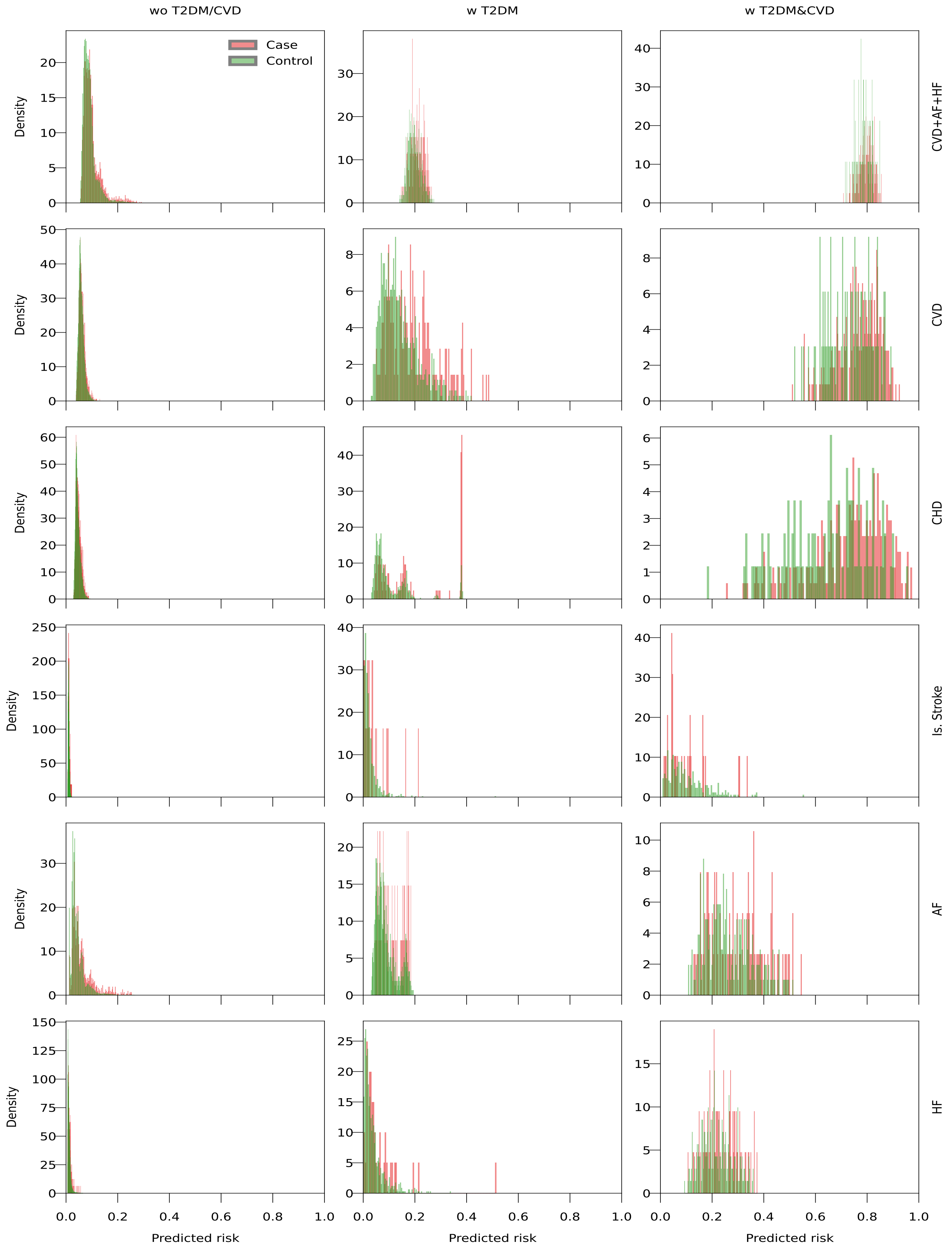

Individuals are stratified as followed “wo T2DM/CVD: participants without T2DM or CVD at baseline, “w T2DM”: participants with type 2 diabetes at baseline, “w T2DM&CVD”: participants with type 2 diabetes at baseline and a history of CVD. The PGS-only models considered information from 29 univariable polygenic scores. See Appendix 3 for the stacked PGS mean and standard deviation (SD).

**Figure 17:** Calibration plots for the PGS-plus models stratified by outcome and participant group.

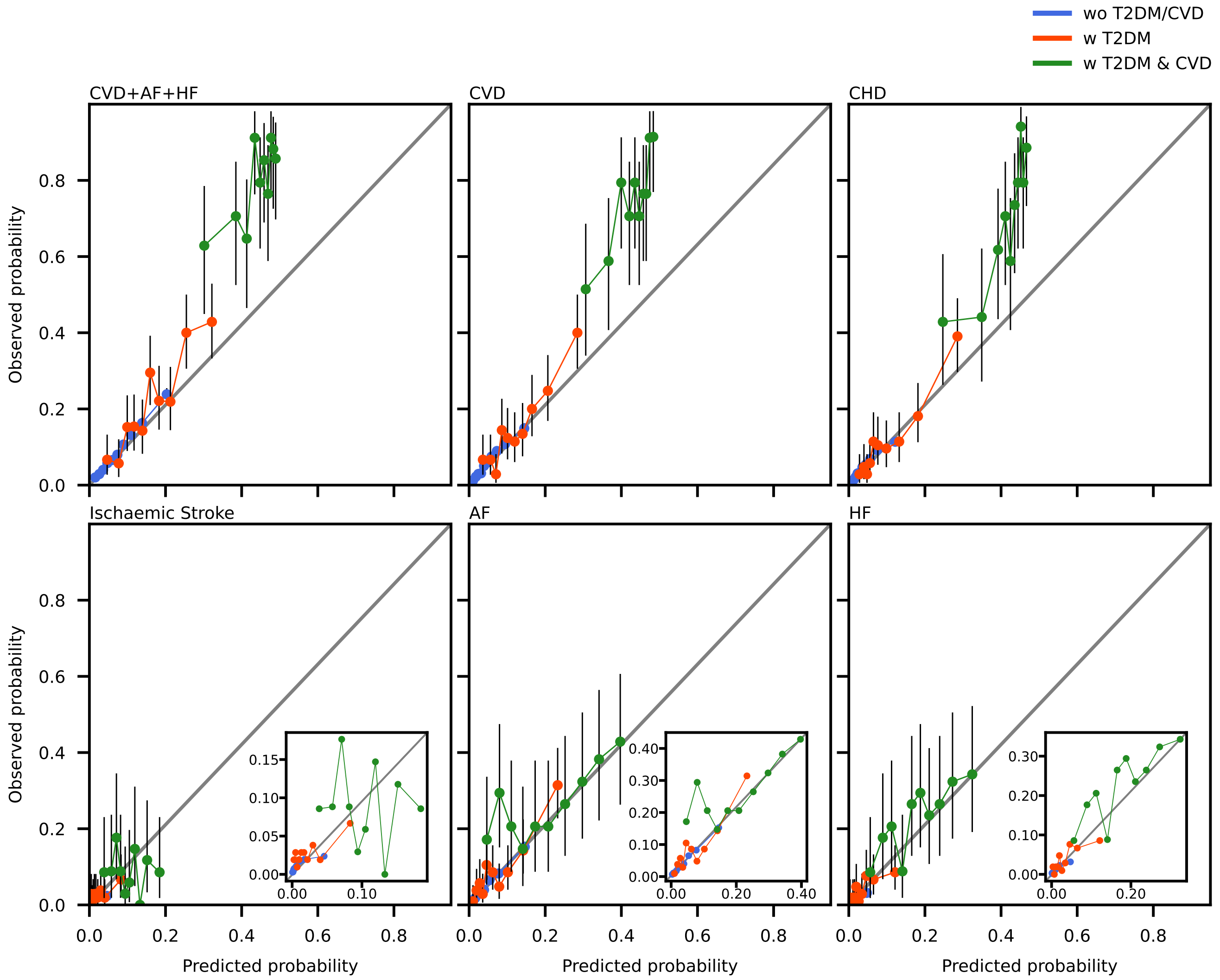

n.b The PGS-plus models considered information from a stacked PGS model, age and sex. Individuals were stratified as followed “wo T2DM/CVD: participants without T2DM or CVD at baseline, “w T2DM”: participants with type 2 diabetes at baseline, “w T2DM&CVD”: participants with type 2 diabetes at baseline and a history of CVD. The vertical line segments represent 95% confidence intervals under the assumption of a beta-distribution.

**Figure 18:** Probability distribution plots for the PGS-plus models stratified by outcome, participant group, and case/control group.

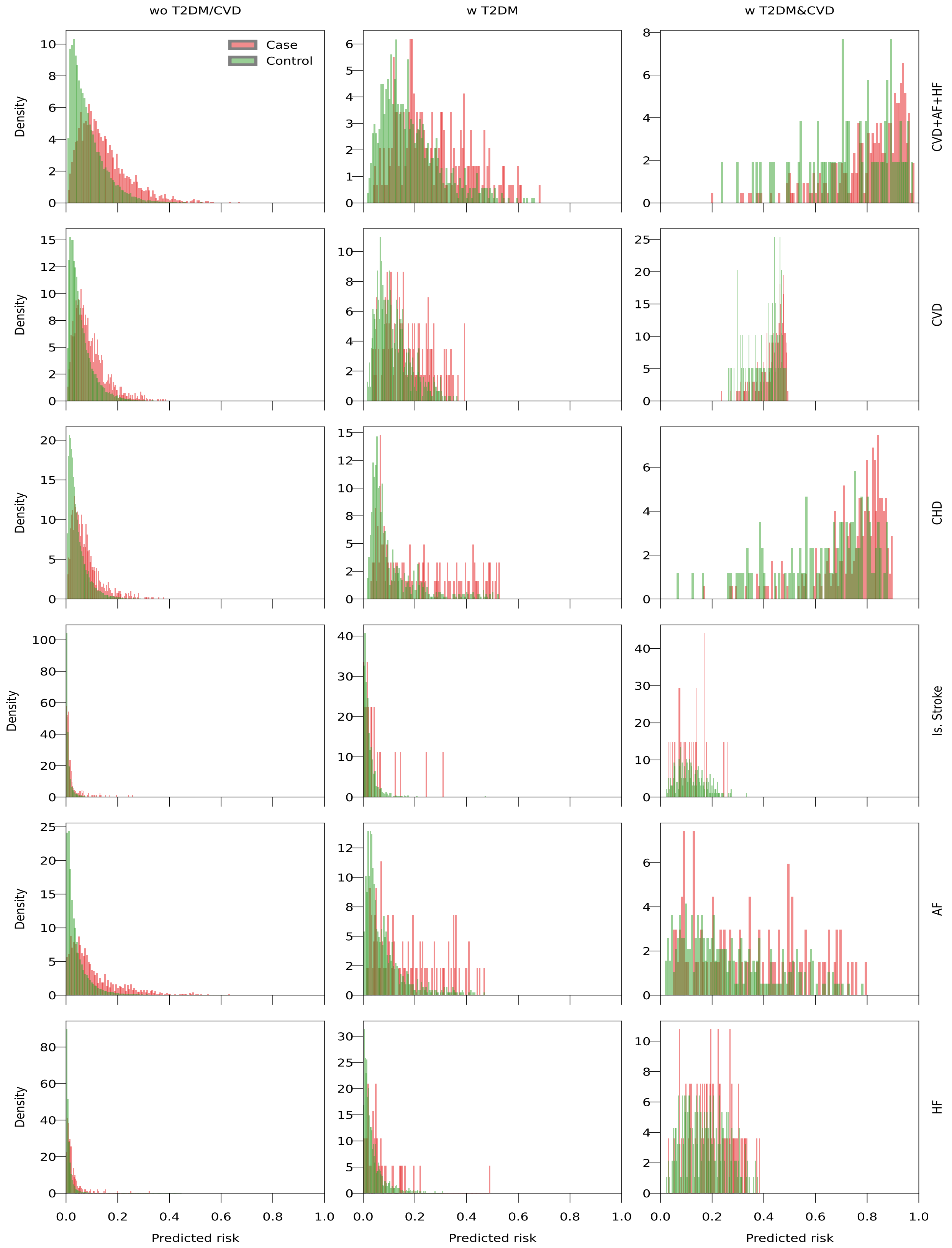

Individuals are stratified as followed “wo T2DM/CVD: participants without T2DM or CVD at baseline, “w T2DM”: participants with type 2 diabetes at baseline, “w T2DM&CVD”: participants with type 2 diabetes at baseline and a history of CVD. The PGS-plus models considered information from a stacked PGS model, age and sex.

**Figure 19:** The contribution of each clinical feature to the prediction of cardiovascular disease.

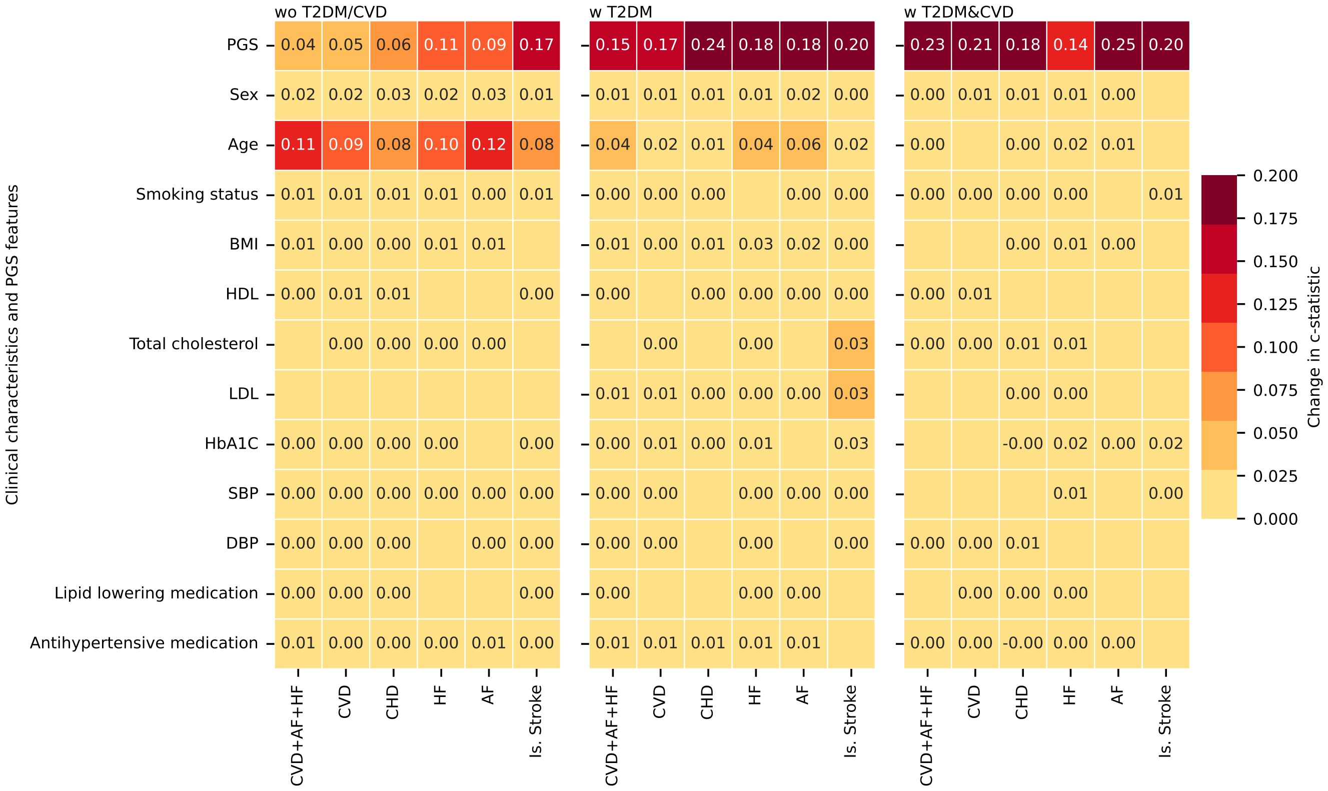

n.b. Individuals are stratified as followed "wo T2DM/CVD: participants without T2DM or CVD at baseline, "w T2DM": participants with type 2 diabetes at baseline, "w T2DM&CVD": participants with type 2 diabetes at baseline and a history of CVD. The permutation feature importance assesses the c-statistic change in the test data; iteratively the values of each variables were randomly assigned to an individual after which the c-statistic was re-estimated with these permuted data and the difference in performance used as an estimate of variable contribution to the model's predictive potential. Empty cells indicate variables with low, almost irrelevant feature importance for a specific model (feature importance is less than 0.0001).

**Figure 20:** Probability distribution plots for the PGS-extended models stratified by outcome, participant group, and case/control status.

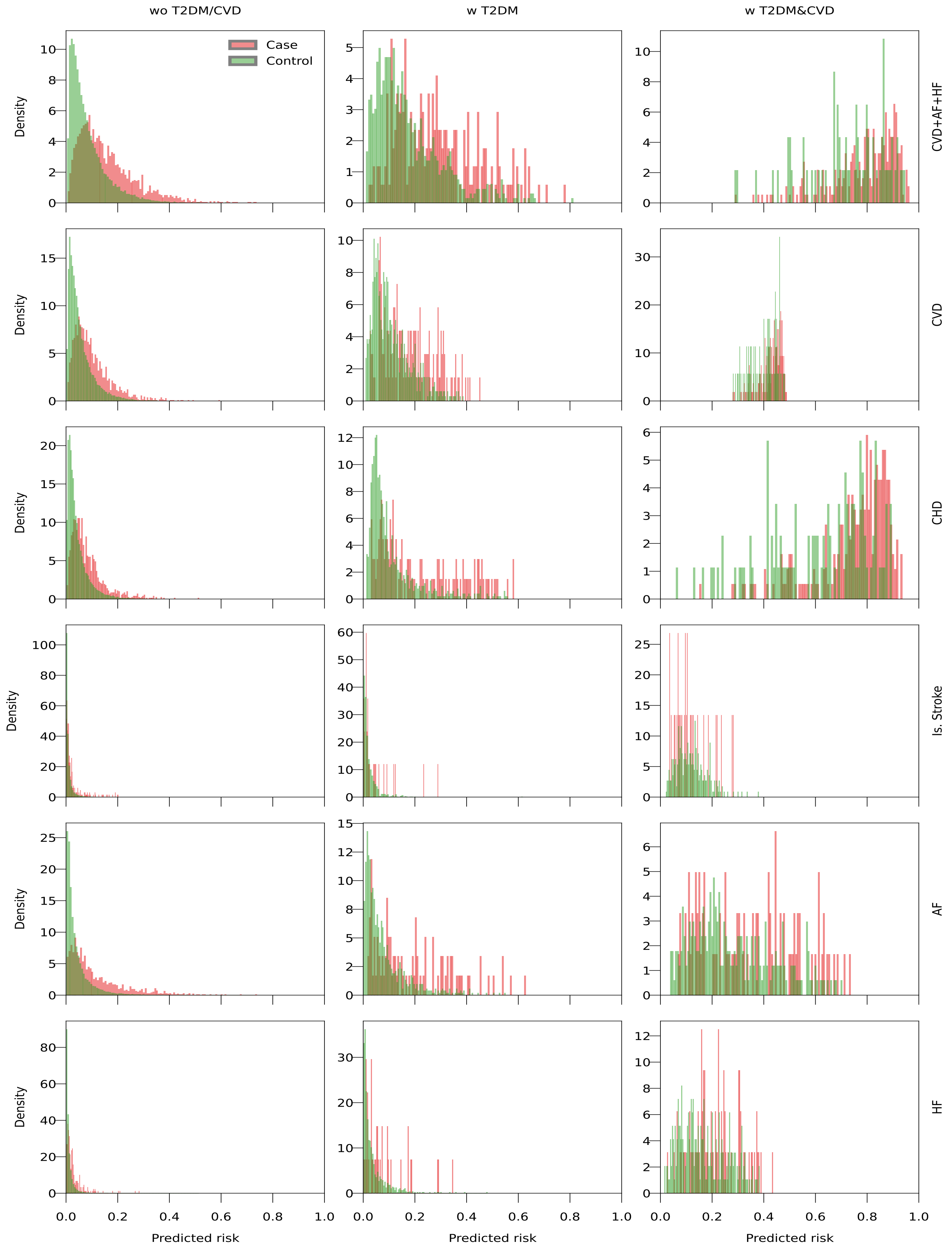

Individuals were stratified as followed "wo T2DM/CVD: participants without T2DM or CVD at baseline, "w T2DM": participants with type 2 diabetes at baseline, "w T2DM&CVD": participants with type 2 diabetes at baseline and a history of CVD. The stacked PGS combined genetic scores for at most 29 GWAS specific PGS, and was combined with non-genetic risk factors: smoking status, blood lipids, blood pressure, BMI, HbA1c, and medicines (PGS-extended).

**Figure 21:** Kaplan-Meier estimates of the 5-year cumulative incident of CVD after an enrolment date for the ”w T2DM” group, stratified by prevalent and incident type 2 diabetes.

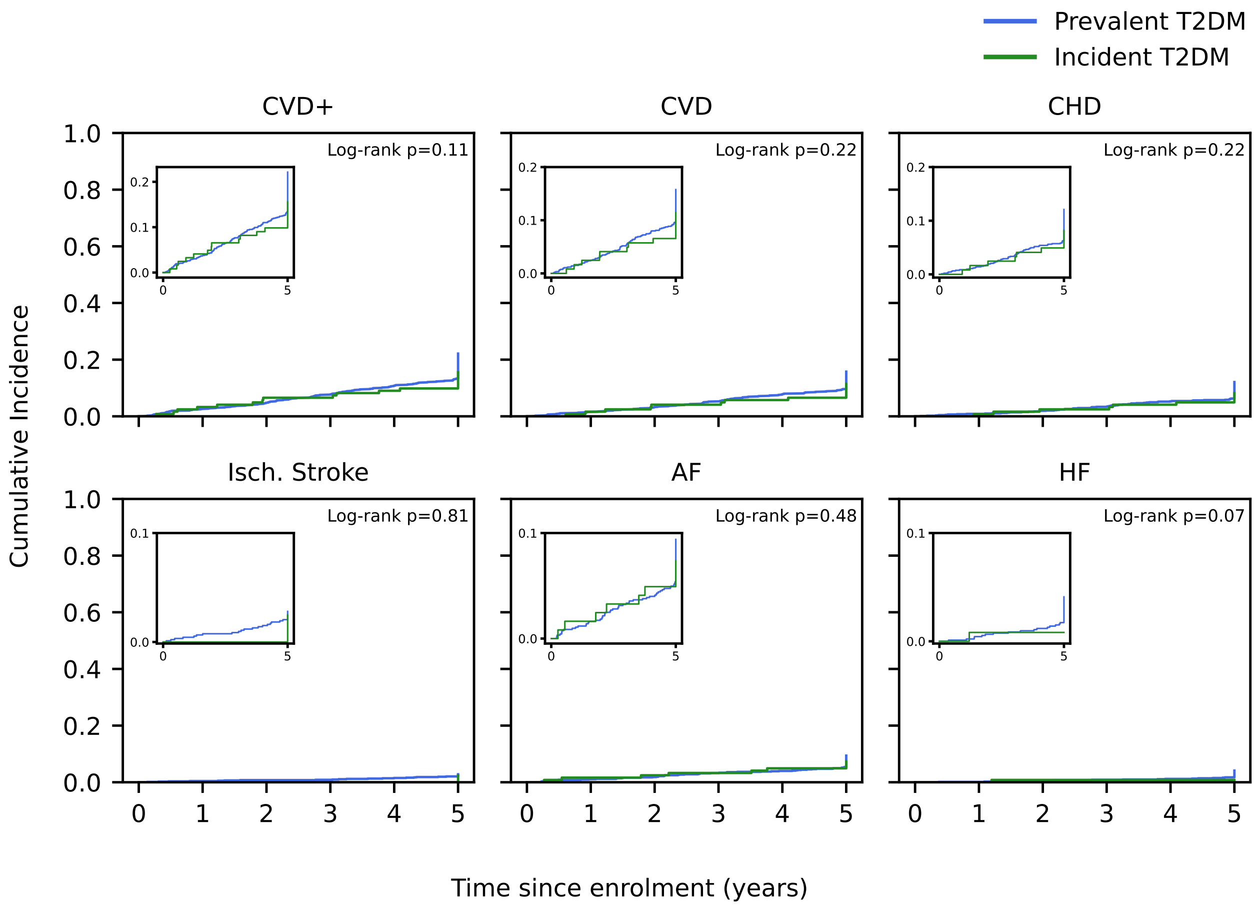

n.b. The ”w T2DM” group indicates individuals with T2DM but without CVD history.

**Figure 22:** Kaplan-Meier estimates of the 5-year cumulative incident of CVD after an enrolment date for the "w T2DM&CVD" group, stratified by prevalent and incident type 2 diabetes.

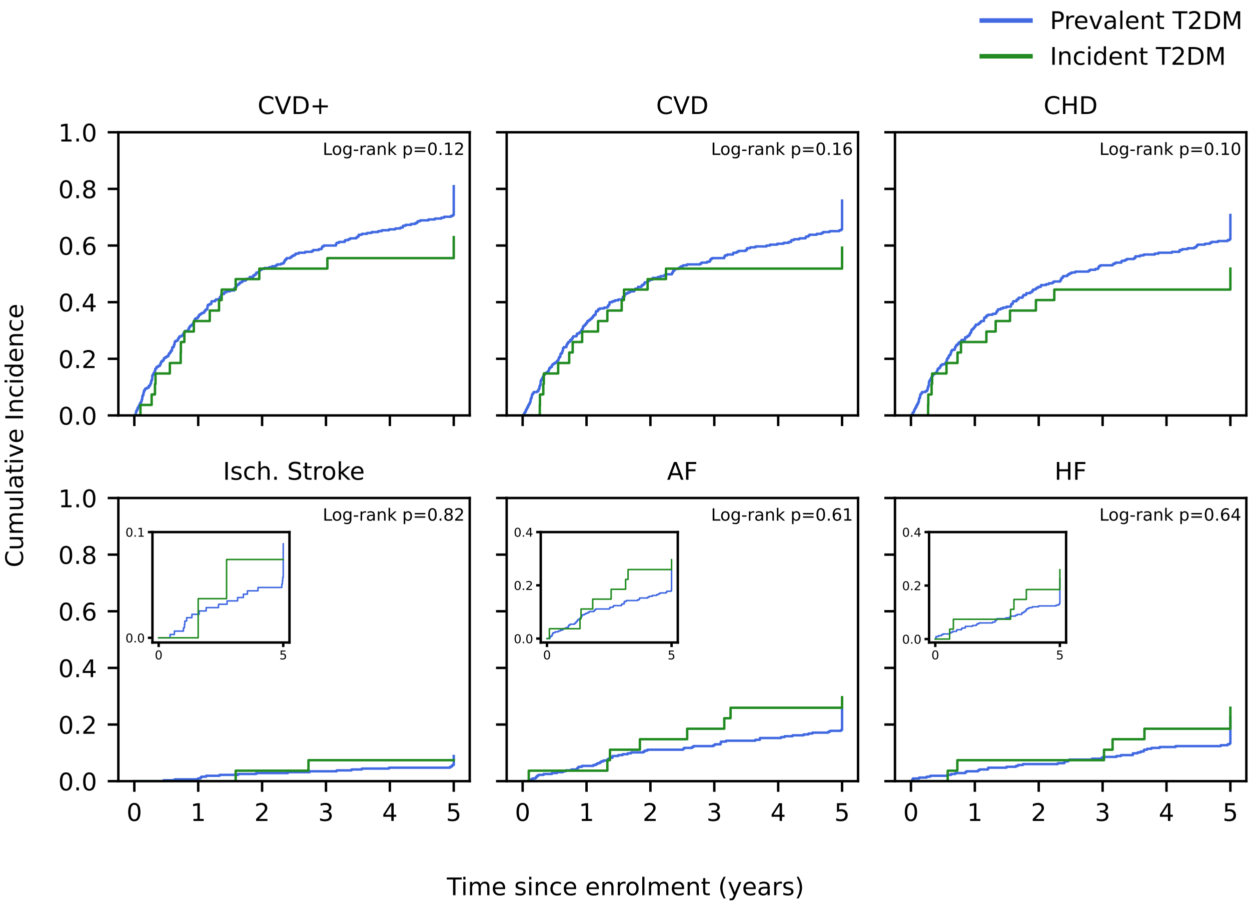

n.b. The "w T2DM&CVD" group indicates individuals with T2DM and a history of CVD.

**Figure 23:** The contribution of each feature (PRS, age, and sex) to the prediction of cardiovascular disease.

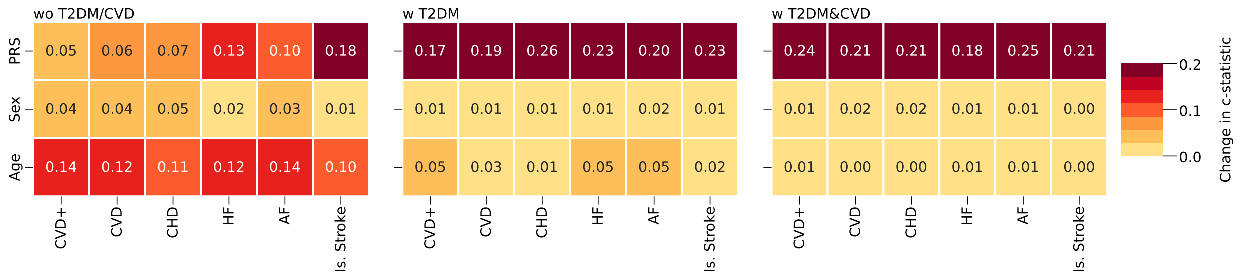

n.b. The permuted feature importance reflects the c-statistic change based on the test data; iteratively the values of each variables were randomly assigned to an individual after which the c-statistic was re-estimated with these permuted data and the difference in performance used as an estimate of variable contribution to the model's predictive potential.

**Figure 24:** Calibration plots for the PGS-extended models stratified by outcome and participant group.

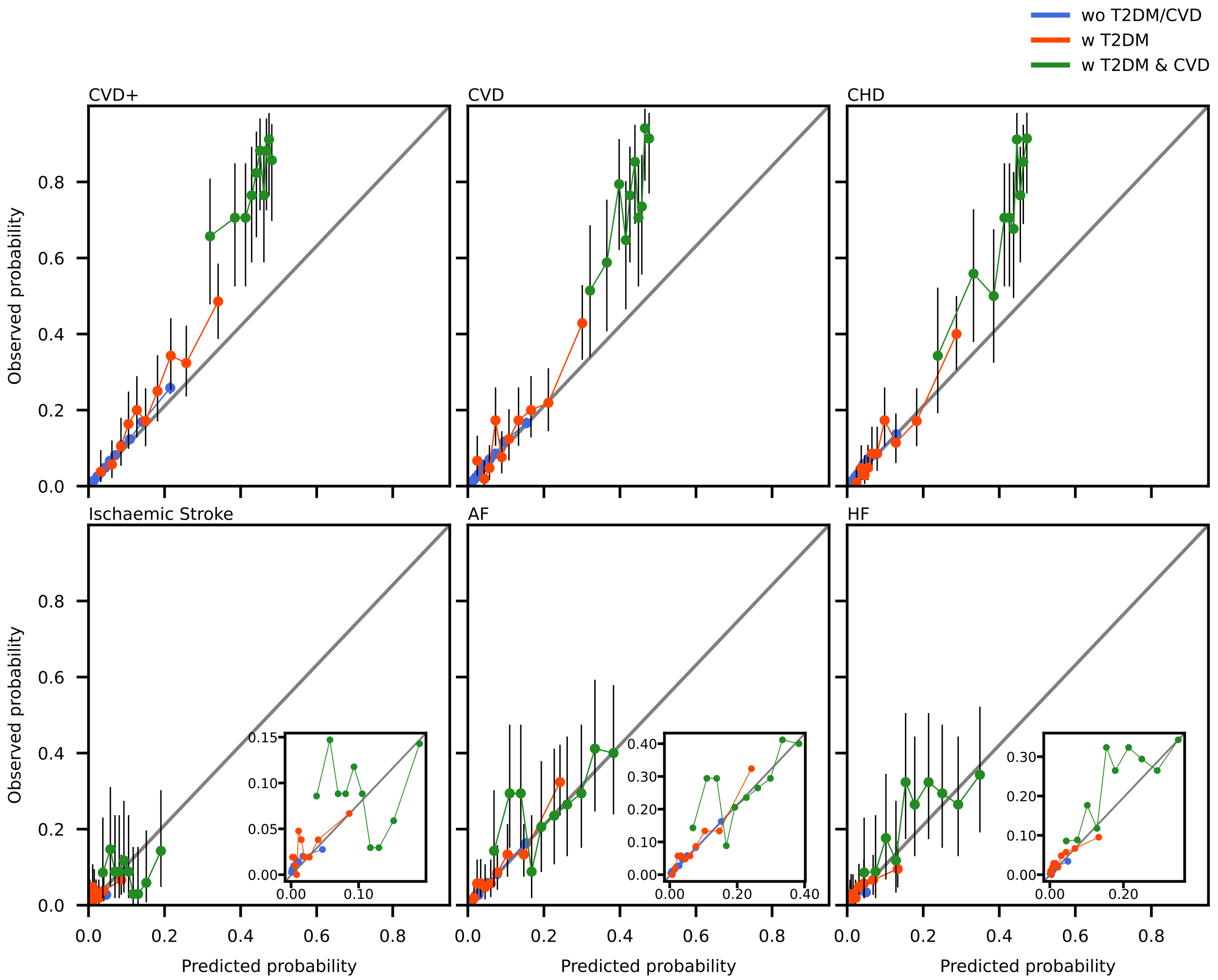

n.b. the PGS-extended models considered information from a stacked PGS model, age, sex smoking status, blood lipids, blood pressure, BMI, HbA1c, CRP, and creatinine (PGS-extended). Individuals were stratified as followed "wo T2DM/CVD: participants without T2DM or CVD at baseline, "w T2DM": participants with type 2 diabetes at baseline, "w T2DM&CVD": participants with T2DM at baseline and a history of CVD. The vertical line segments represent 95% confidence intervals calculated using the Clopper-Pearson method.
