## Appendix 3 for "Combining stacked polygenic scores with clinical risk factors improves cardiovascular risk prediction in people with type 2 diabetes"

| Group | Outcome | PGS Mean | PGS SD | Status |
|---|---|---|---|---|
| wo T2DM/CVD | CVD + AF + HF | -2.3570906990514127 | 0.28393699434913483 | Case |
| wo T2DM/CVD | CVD + AF + HF | -2.435171033205664 | 0.24706280572100672 | Control |
| w T2DM | CVD + AF + HF | -1.5703706300529936 | 0.12132636847327737 | Case |
| w T2DM | CVD + AF + HF | -1.627422419510718 | 0.12248721799588 | Control |
| w T2DM&CVD | CVD + AF + HF | -0.2289477010332008 | 0.03500419235224144 | Case |
| w T2DM&CVD | CVD + AF + HF | -0.240070705592204 | 0.03764723472353872 | Control |
| wo T2DM/CVD | CVD | -2.8001564779709587 | 0.18772582958374517 | Case |
| wo T2DM/CVD | CVD | -2.8444739964531065 | 0.1804481752981166 | Control |
| w T2DM | CVD | -1.5522870680270844 | 0.6450387033425716 | Case |
| w T2DM | CVD | -1.9239929810552177 | 0.58330440896246 | Control |
| w T2DM&CVD | CVD | 1.2507992652845834 | 0.46256423701440985 | Case |
| w T2DM&CVD | CVD | 1.055187963064099 | 0.463871141618513 | Control |
| wo T2DM/CVD | CHD | -3.055144412489871 | 0.2022573403338721 | Case |
| wo T2DM/CVD | CHD | -3.1085482047923576 | 0.19672802703610165 | Control |
| w T2DM | CHD | -1.8968952820013036 | 0.7489010884211786 | Case |
| w T2DM | CHD | -2.4316062601219732 | 0.5900360903305848 | Control |
| w T2DM&CVD | CHD | -0.3418433568384666 | 0.2374471948872424 | Case |
| w T2DM&CVD | CHD | -0.4879197436939358 | 0.29056216822827624 | Control |
| wo T2DM/CVD | Ischemic stroke | -4.542423171648233 | 0.22949185601327698 | Case |
| wo T2DM/CVD | Ischemic stroke | -4.591848152738551 | 0.21496432163241475 | Control |
| w T2DM | Ischemic stroke | -3.7946887548796404 | 1.2079725649365278 | Case |
| w T2DM | Ischemic stroke | -4.142196471391346 | 0.9045728715789572 | Control |
| w T2DM&CVD | Ischemic stroke | -2.6227361720132802 | 0.8365596089210157 | Case |
| w T2DM&CVD | Ischemic stroke | -2.5547187443216623 | 0.7794091045210637 | Control |
| wo T2DM/CVD | AF | -2.978129098711699 | 0.5990561623785567 | Case |
| wo T2DM/CVD | AF | -3.2728407812080658 | 0.5303586112442533 | Control |
| w T2DM | AF | -2.273239210716222 | 0.434340276529167 | Case |
| w T2DM | AF | -2.5316846400757647 | 0.4278461080703035 | Control |
| w T2DM&CVD | AF | -1.2600855569932892 | 0.37661618364277827 | Case |
| w T2DM&CVD | AF | -1.380843360662616 | 0.3493523353805223 | Control |
| wo T2DM/CVD | HF | -4.41044289124218 | 0.4155039564584552 | Case |
| wo T2DM/CVD | HF | -4.537776518769428 | 0.3846114842964483 | Control |
| w T2DM | HF | -3.2647273674271915 | 1.1228185872951044 | Case |
| w T2DM | HF | -3.75034165474536 | 0.9953122419609423 | Control |
| w T2DM&CVD | HF | -1.1955015372413107 | 0.3547356482816017 | Case |
| w T2DM&CVD | HF | -1.299879037759858 | 0.3674153282973063 | Control |
